# The trend of hypertension-related chronic kidney disease from 1990 to 2019 and its predictions over 25 years: An analysis of the Global Burden of Disease Study 2019

**DOI:** 10.1101/2023.03.21.23287527

**Authors:** Yi Ren, Honggang Zhang

## Abstract

**Background:** High blood pressure is a key pathogenetic factor that contributes to the deterioration of kidney function. However, the incidence trend of hypertension-related chronic kidney disease (CKD) has rarely been studied; therefore, we aimed to analyze the global, regional, and national patterns, temporal trends as well as burden of hypertension-related CKD.

**Methods:** We extracted data on hypertension-related CKD from the Global Burden of Disease (GBD) study database, including the incidence, prevalence, disability-adjusted life years (DALYs), and mortality numbers and rates (per 100,000 population) and further described according to year, location, sex, age, and socio-demographic index (SDI). The estimated annual percentage changes (EAPCs) were calculated to assess the variation in incidence, DALYs, and mortality. We used an age-period-cohort (APC) model framework to analyze the underlying trends in prevalence by age, period, and birth cohort. Nordpred APC analysis was performed to predict the future morbidity and mortality of hypertension-related CKD.

**Results:** In 2019, a total of over 1.57 million new hypertension-related CKD cases were reported worldwide, a 161.97% increase from 1990. Compared to 1990, the ASIR increased in all 21 regions in 2019. In all countries and territories except Iceland, the EAPC in ASIR and the lower boundary of its 95% confidence interval (CI) were higher than 0. ASIR, ASPR, ASDR, and ASMR were not identical among countries with different SDI regions in 2019; additionally, ASIR and ASMR were significantly different among sexes in all SDI regions in 2019. The predicted incidence and mortality counts globally continue to increase to 2044, and there is an upward trend in ASIR for both men and women.

**Conclusions:** Hypertension-related CKD cannot be ignored either as a subcategory of CKD or as a complication of hypertension. Between 1990 and 2019, the ASIR of hypertension-related CKD demonstrated an ascending trend, and according to our projections, it would remain on the rise for the next 25 years. With remarkable global population growth, aging, and an increasing number of patients with hypertension, the burden of disease caused by hypertension-related CKD continues to increase.

## Introduction

There is a substantial burden of both chronic kidney disease (CKD) and impaired kidney function at the global level, and CKD is becoming an important public health problem[1]. CKD is defined as kidney damage or GFR<60 ml/min/1.73 m^2^ for more than 3 months, and kidney damage is described as abnormal renal function (e.g., proteinuria or albuminuria, or abnormal urine sediment, e.g., dysmorphic red cells) or structural abnormalities noted on imaging studies[2]. The prevalence of hypertension is rising globally owing to the aging of the population and increases in exposure to lifestyle risk factors, including unhealthy diets (i.e., high sodium and low potassium intake) and lack of physical activity[3]. Hypertension has become the second or third leading cause of renal replacement therapy (RRT) in Europe, alongside glomerulonephritis, in recent years[4]. Between 1975 and 2015, the highest worldwide blood pressure levels have shifted from high-income countries to low-income countries in south Asia and sub-Saharan Africa, while blood pressure has been persistently high in central and eastern Europe[5]. The number of patients with raised blood pressure has risen worldwide, with the increase occurring mainly in low-income and middle-income countries, which implies that hypertension affects the health of the global population and increases the burden of disease on people in low-income countries in particular.

Although hypertension can be well managed with the use of appropriate medication, the harm of hypertensive complications, including acute cerebral hemorrhage and acute myocardial infarction, has been recognized. However, hypertension-related CKD may be inadequately addressed, and limited quantitative assessments of the burden of this disease have been performed. In our study, we analyzed recent trends in the incidence, prevalence, DALYs, and deaths of hypertension-related CKD from 1990 to 2019 at the global, regional, and national levels using data from the Global Burden of Disease (GBD) study 2019. Gender, age, and socio-demographic index (SDI) disparities in the burden of this disease were revealed, providing some enlightening guidance for medical agencies, policymakers, and the general public.

## Materials and methods

### Study Data

Data regarding hypertension-related CKD, including global, regional, and national age-standardized incidence rates (ASIR), age-standardized prevalence rates (ASPR), age-standardized disability-adjusted life years (DALYs) rates (ASDR), age-standardized mortality rates (ASMR) and absolute numbers of incidence, prevalence, DALYs, and mortality, were acquired from the GBD 2019 study (http://ghdx.healthdata.org/gbd-results-tool), which provides the largest and most recent comparative assessment of the burdens of diseases and injuries in global and 204 countries and territories from 1990 to 2019. DALYs were the total healthy life years lost from onset to death, including both years of life lost due to premature death and years lived with disability (DALY= YLLs+ YLDs). Ethics approval and informed consent were exempted from this study because of public accessibility to the data.

The SDI is a composite indicator estimated from the lagged distribution of per capita income, total fertility rate for females under age 25, and average years of schooling for adults aged 15 and older. Data on SDI were available for 204 countries in 2019 (https://ghdx.healthdata.org/record/ihme-data/gbd-2019-socio-demographic-index-sdi-1950-2019) and were classified into five groups, including high (n = 39), high-middle (n = 48), middle (n = 41), low-middle (n = 43), and low (n = 33) SDI countries and territories.

### Statistical Analysis

ASIR, ASPR, ASDR, and ASMR were expressed as the number per 100,000 population with a 95% uncertainty interval (UI). The natural logarithm of the rates was calculated by using the following regression model, y = α+βx+ε, where y denotes ln (ASR), x denotes the calendar year, and ε denotes the error term; estimated annual percentage change (EAPC) = 100×(exp(β)-1); moreover, its 95% confidence interval (CI) can also be obtained from the above method. The ASR is deemed to be in an increasing trend if the EAPC estimation and the lower boundary of its 95% CI are both >0. In contrast, the ASR is in a decreasing trend if the EAPC estimation and the upper boundary of its 95% CI are both <0. Otherwise, the ASR is deemed to be stable over time. The Wilcoxon signed-rank test was used to compare sex differences in ASIR, ASPR, ASDR, and ASMR among the five SDI regions. Pearson correlation was performed to evaluate the relationship between the burden of hypertension-related CKD (ASIR, ASPR, ASDR, and ASMR) and SDI in 2019. We used decomposition analyses to quantify the superimposed contribution of the effect of the differences in factors in the population to their overall variation in value. Decomposition of morbidity and mortality of hypertension-related CKD by age structure, population growth, and epidemiological change allows the quantification of the contribution of each of these factors to the overall effect.

To analyze and estimate the effects of age, period, and cohort on the global prevalence of hypertension-related CKD, we applied Stata SE 16 software (StataCorp, College Station, TX, United States) using the age-period-cohort (APC) model and IE method. Generally, the APC model fits a log-linear Poisson model and can be expressed as log(Y_i_)=μ+ α×age_i_+β×period_i_+γ×cohort_i_+ε, where Y_i_ is the CKD prevalence or mortality rate, α, β, and γ are the coefficients of age, period and cohort, respectively, μ is the intercept and ε is the residual of the model. The intrinsic estimator (IE) method integrated into the age-period-cohort model was used to obtain the net effects for three dimensions[6]. The relative risk (RR) values for each age, period, and cohort represent the independent risks in comparison with the reference group. This approach has been adopted in descriptive epidemiology for certain chronic diseases[7,8]. Moreover, we predicted the number of new cases and deaths of hypertension-related CKD from 2019 to 2044 by running a Nordpred APC analysis using the Nordpred package in R, taking into account the changing rates and changing population structure, which has been well demonstrated and recognized in previous studies[9].

Statistical analyses were performed using R Statistical Software (version 4.2.1; R Foundation for Statistical Computing, Vienna, Austria). A *P* value less than 0.05 was considered statistically significant.

## Results

### Global burden of hypertension-related CKD

Globally, the incidence cases of hypertension-related CKD grew by 161.97% from 602,690 (95% UI: 548,661 to 659,099) in 1990 to 1,578,842 (95% UI: 1,447,111 to 1,715,250) in 2019, and ASIR continued to rise over these three decades (Figure 1). Despite an increase in prevalence number as well, the ASPR of hypertension-related CKD peaked in 2018 (432.74 per 100,000 population, 95% UI: 401.42 to 432.74), and it was no longer higher in 2019 (Figure 1). From 1990 to 2019, there was a moderate trend in the worldwide ASIR and ASPR of CKD caused by hypertension, with EAPC values of 0.69 (95% CI: 0.68 to 0.70) and 0.68 (95% CI: 0.65 to 0.7), respectively (Supplementary Table S1 and Table S2).

**Figure 1:**
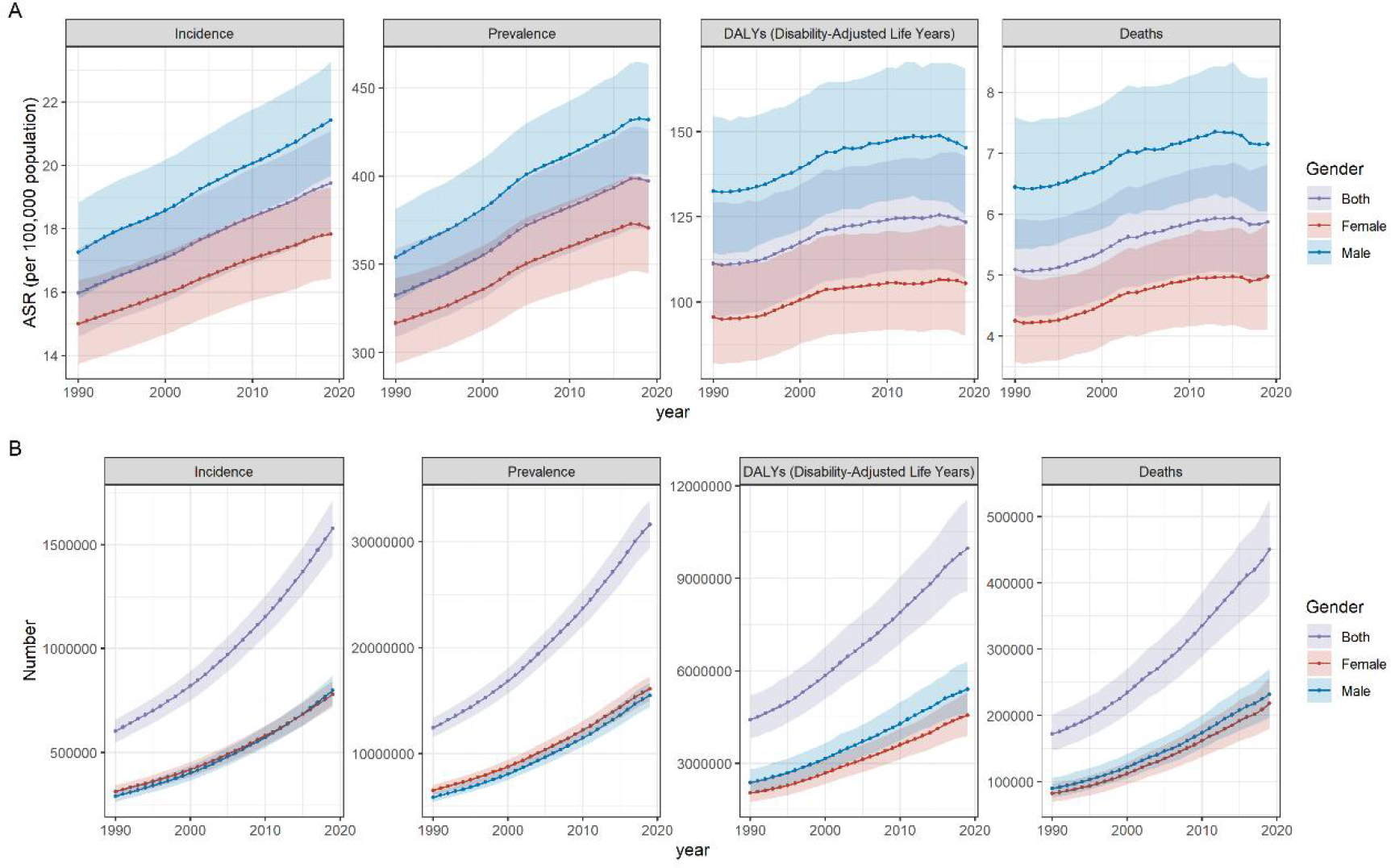
Trends in global of hypertension-related CKD: age-standardized rates (ASR) (A) and numbers (B) of incidence, prevalence, DALYs, and deaths from 1990 to 2019. Sbade areas represent 95% uncertainty intervals.

The ASDR of hypertension-related CKD rose from 111.27 (95% UI: 96.36 to 129.18) per 100,000 population in 1990 to 123.41 (95% UI: 106.86 to 142.66) per 100,000 population in 2019. The ASDR peaked in 2016 (125.59 per 100,000 people; 95% UI: 109.65 to 143.71), after which it began to decline (Figure 1). The global mortality count of hypertension-related CKD increased by 161.90% from 171,878 (95% UI: 146,475 to 201,264) in 1990 to 450,147 (95% UI: 381,995 to 525,405) in 2019. The ASMR peaked in 2015 (5.94 per 100,000 people; 95% UI: 5.06 to 6.85), after which it began to decrease; however, it recovered in 2018 (Figure 1). From 1990 to 2019, the EAPC of ASDR and ASMR were 0.49 (95% CI: 0.43 to 0.55) and 0.64 (95% CI: 0.57 to 0.72), respectively, indicating an increase in the burden of CKD brought on by hypertension (Supplementary Table S3 and Table S4).

### Regional burden of hypertension-related CKD

All countries and territories were divided into 21 regions according to epidemiological similarities and geographical proximity. At the regional level, the number of incident cases in East Asia was 283,695 (95% UI: 255,672 to 312,985), which ranked first among the 21 geographical regions in 2019. However, the highest ASIR and ASPR were recorded in North Africa and Middle East (36.55 per 100,000 population; 95% UI: 33.59 to 39.58) and Central Latin America (705.04 per 100,000 population; 95% UI: 653.68 to 760.29) in 2019, respectively. All 21 regions experienced an increase in ASIR, with the most significant ASIR increase detected in Andean Latin America (EAPC=2.26; 95% CI: 2.15 to 2.37), followed by North Africa and Middle East (EAPC=2.00; 95% CI: 1.9 to 2.1). The highest EAPC of ASPR was similarly found in Andean Latin America (EAPC=1.99; 95% CI: 1.9 to 2.08) (Supplementary Table S1 and Table S2).

Between 1990 and 2019, the highest ASDR and ASMR were both observed in Southern Sub-Saharan Africa, 317.60 per 100,000 population (95% UI: 275.47-366.77) and 15.12 per 100,000 population (95% UI: 13.06-17.38), respectively, in 2003. In 2019, Central Latin America ranked first in terms of ASDR (279.03 per 100,000 population; 95% UI: 228.09 to 338.68). The most pronounced ASDR increase occurred in Central Latin America (EAPC=2.47; 95% CI: 2.14 to 2.80), whereas the most pronounced decrease was detected in High-income Asia Pacific (EAPC=-1.37; 95% CI: -1.65 to -1.09). The highest deaths cases and ASMR in 2019 were recorded in East Asia (75,016; 95% UI: 60,877 to 89,893) and Central Latin America (12.51 per 100,000 persons; 95% UI: 9.94 to 12.51), respectively. The most pronounced ASMR increase was observed in Central Latin America (EAPC=2.33; 95% CI: 2.01 to 2.64), followed by High-income North America and Andean Latin America, while the most pronounced decrease was detected in High-income Asia Pacific (EAPC=-1.61; 95% CI:-1.97 to -1.24) (Supplementary Table S3 and Table S4). Additionally, ASIR, ASPR, ASDR, and ASMR were correlated with SDI, with Pearson correlation coefficients of 0.549, 0.438, -0.601, and -0.554, respectively (p < 0.001) (Figure 2).

**Figure 2:**
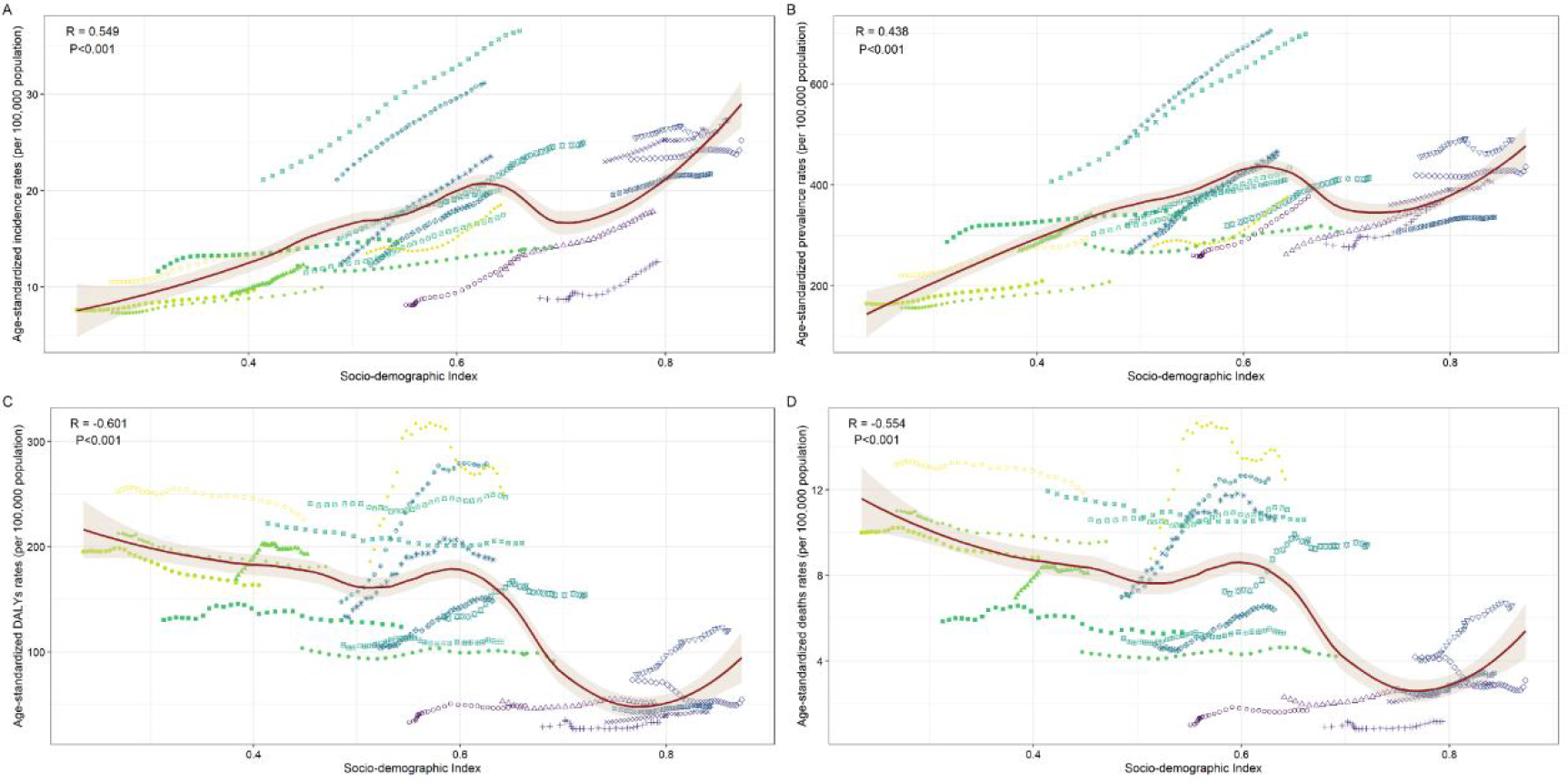
The trend in age-standardized incidence rates (ASIR) (A), age-standardized prevalence rates (ASPR) (B), age-standardized disability-adjusted life year rates (ASDR) (C), and age-standardized mortality rates (ASMR) (D) of hypertension-related CKD across 21 geographical regions and global by socio-demographic index (SDI) for both sexes combined, 1990-2019. For each region, points from left to right depict estimates from each year from 1990 to 2019. Sbade areas represent 95% uncertainty intervals.

### National burden of hypertension-related CKD

At the national level, the highest absolute number of newly diagnosed hypertension-related CKD cases was recorded in China in 2019 (268,862; 95% UI: 241,803 to 297,432), followed by India (166,500; 95% UI: 149,302 to 183,791) and the United States of America (157,609; 95% UI: 144,239 to 172,082) (Figure 3A). The highest ASIR of hypertension-related CKD in 2019 occurred in Saudi Arabia (45.66 per 100,000 population; 95% UI: 41.39 to 50.22) (Figure 3B), and Saudi Arabia also had the highest ASPR (926.25 per 100,000 people; 95% UI: 841.29 to 1,008.65). Between 1990 and 2019, the most pronounced incidence cases increase was observed in Qatar (1551.90%; 95% CI: 1445.91% to 1663.39%). From 1990 to 2019, the highest EAPC of ASIR and ASPR were found in Morocco, 2.67 (95% CI: 2.6 to 2.74) (Figure 3C) and 2.69 (95% CI: 2.64 to 2.74), respectively, followed by Turkey (EAPC of ASIR: 2.45; 95% CI: 2.27 to 2.64; EAPC of ASPR: 2.44; 95% CI: 2.22 to 2.65) and Ecuador (EAPC of ASIR: 2.40; 95% CI: 2.26 to 2.53; EAPC of ASPR: 2.25; 95% CI: 2.13 to 2.37), whereas Ireland was the only country in which the EAPC and the upper boundary of its 95% CI were both less than 0 in ASIR and ASPR among all countries (Supplementary Table S5 and Table S6).

**Figure 3:**
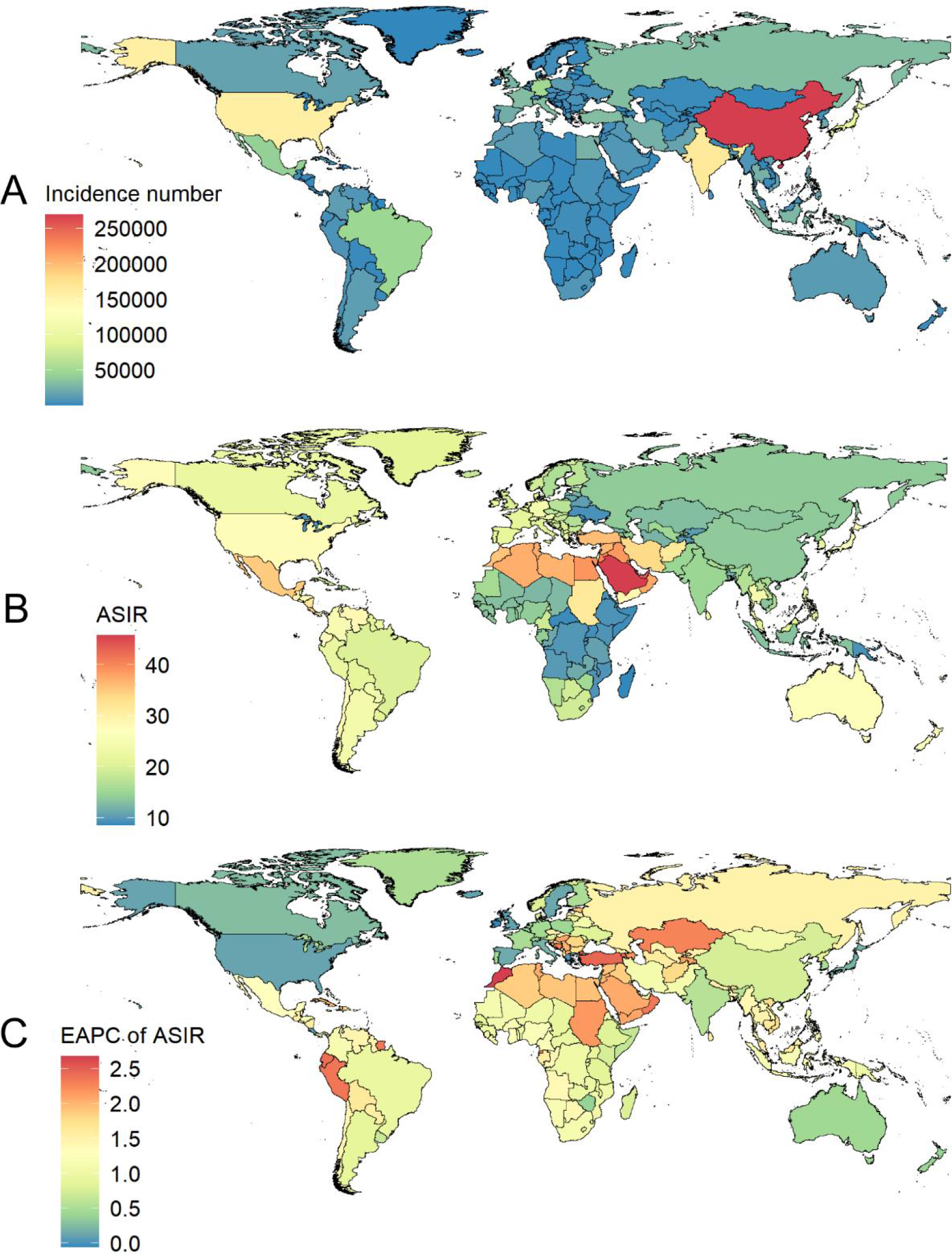
The incidence of hypertension-related CKD for both genders in 204 countries and territories. (A) Absolute number of incidence in 2019 of hypertension-related CKD. (B) Age-standardized incidence rates (ASIR) in 2019 of hypertension-related CKD. (C) Estimated annual percentage change (EAPC) of hypertension-related CKD ASIR from 1990 to 2019.

From 1990 to 2019, the most significant DALYs count increase was observed in the United Arab Emirates (727.93%; 95% CI: 1445.91% to 1663.39%). The highest ASDR of hypertension-related CKD occurred in Mauritius (635.53 per 100,000 population; 95% UI: 496.72 to 794.12) in 2019. Between 1990 and 2019, the highest EAPC of ASDR was found in Estonia (4.29; 95% CI: 3.92 to 4.66), followed by El Salvador (4.11; 95% CI: 3.41 to 4.82), whereas Mongolia (−2.68; 95% CI: -3.05 to -2.31) showed the lowest EAPC of ASDR (Supplementary Table S7). The highest absolute number of deaths from hypertension-related CKD was recorded in China in 2019 (70,260; 95% UI: 56,866 to 84,481), followed by India (50,909; 95% UI: 39,096 to 64,888) and the United States of America (43,329; 95% UI: 35,082 to 50,786). Absolute death numbers of hypertension-related CKD increased in all countries except Mongolia, with the greatest increase observed in Estonia (669.28%; 95% CI: 880.86% to 438.5%). The most significant increase and decrease in ASMR were detected in Estonia (EAPC =6.23; 95% CI: 5.75 to 6.70) and Mongolia (EAPC = -3.25; 95% CI: -3.7 to -2.8), respectively (Supplementary Table S8).

### Burden of hypertension-related CKD by SDI

The SDI ranged from 0 to 1, while a greater SDI indicated better socioeconomic development and was classified as high SDI, high-moderate SDI, moderate SDI, low-moderate SDI, and low SDI. Between 1990 and 2019, the most significant increases in ASIR and ASPR were both detected in the middle SDI, with EAPC values of 1.22 (95% CI: 1.19 to 1.25) and 1.12 (95% CI: 1.07 to 1.18), respectively (Supplementary Table S1 and Table S2). Only low-SDI regions showed a decreasing trend in ASDR and ASMR, with EAPC values of -0.41 (95% CI:-0.47 to -0.34) and -0.45 (95% CI:-0.52 to -0.37), respectively. The most significant increases in ASDR and ASMR were noted in high-SDI regions, with EAPC values of 0.98 (95% CI: 0.93 to 1.04) and 1.26 (95% CI: 1.15 to 1.37), respectively (Supplementary Table S3 and Table S4).

The Kruskal-Wallis test revealed that ASIR (χ^2^ = 59.975; df = 4; p < 0.0001) and ASPR (χ^2^ = 60.389; df = 4; p < 0.0001) were not identical among countries with different SDI regions (at least 4 groups significantly different) in 2019. At the same time, there were also differences in ASDR (χ^2^ = 58.969; df = 4; p < 0.0001) and ASMR (χ^2^ = 49.431; df = 4; p < 0.0001) for different SDI regions (at least 4 groups significantly different). Generally, high-middle and high SDI regions had higher ASIR in 2019; low-middle SDI and middle SDI regions had higher ASMR in 2019 (Supplementary Figure S1).

The Wilcoxon signed-rank test revealed that ASIR was significantly different among genders in all SDI regions (p < 0.05), and it showed gender inequality in middle (W = 569; p < 0.05), high-middle (W = 809; p < 0.05) and high (W = 469; p < 0.01) SDI regions in terms of ASPR in 2019. We also found gender inequality in terms of ASDR in low (W = 218; p < 0.0001), low-middle (W = 649; p < 0.05), middle (W = 597; p < 0.05), and high (W = 446; p < 0.01) SDI regions, and ASMR was significantly different among genders in all SDI regions (p < 0.05) in 2019 (Figure 4).

**Figure 4:**
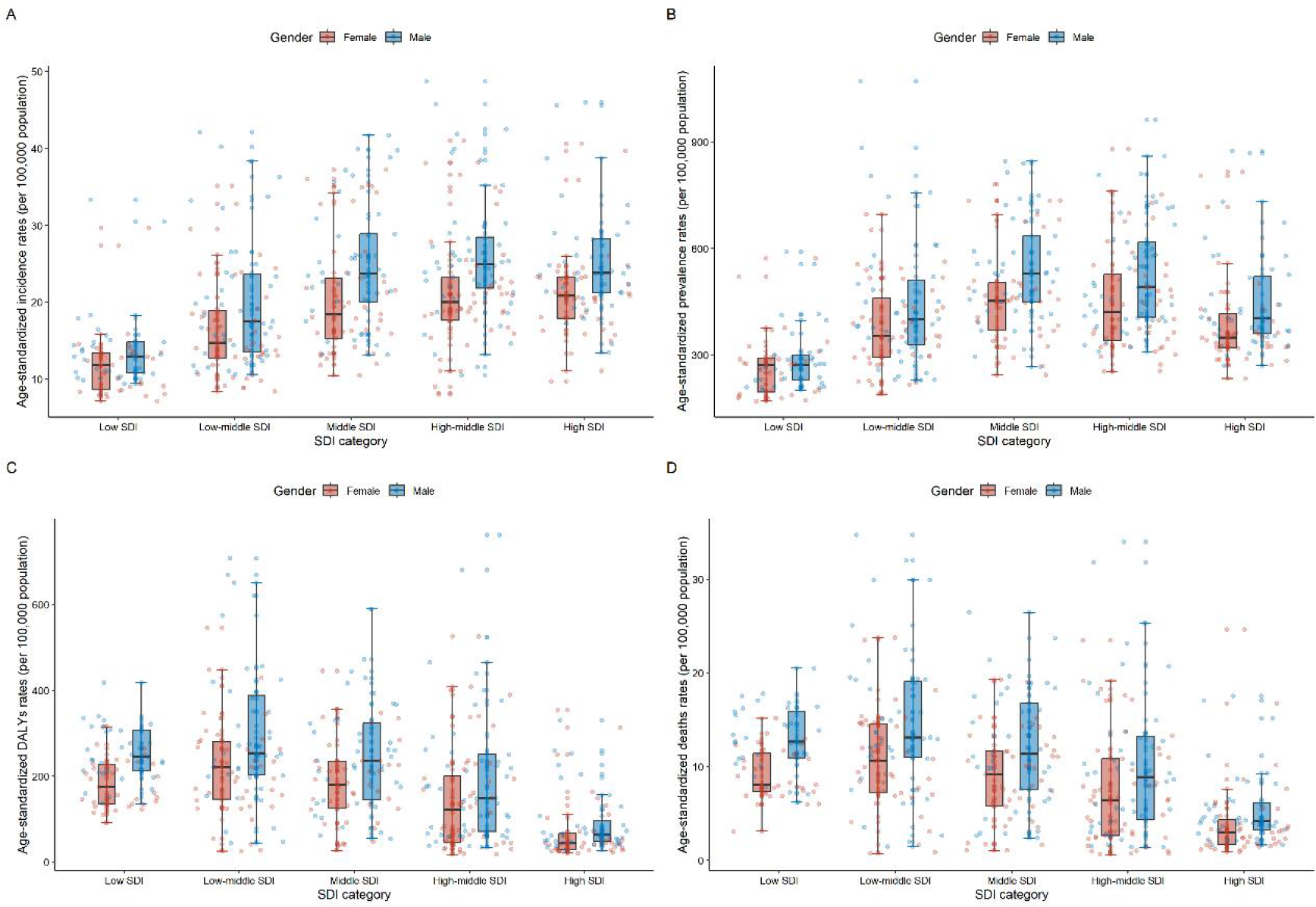
Gender-specific burden in terms of age-standardized incidence rates (ASIR) (A), age-standardized prevalence rates (ASPR) (B), age-standardized disability-adjusted life years rates (ASDR) (C), and age-standardized mortality rates (ASMR) (D) at 204 countries in 2019.

At the global and SDI levels, decomposition analyses of morbidity and mortality by population growth, aging, and epidemiological change were performed (Supplementary Table S9 and Table S10). Between 1990 and 2019, there was a significant increase in the incidence and deaths of hypertension-related CKD globally and in each SDI quintile, most notably in the middle SDI quintile, which experienced the largest increases in incidence and deaths (Figure 5A and 5B). The epidemiological change that captures the underlying change in age and population-adjusted morbidity of hypertension-related CKD over the past 30 years increased globally and was evident in the middle and high-middle SDI quintiles (Figure 5A), while the contribution of epidemiological change to overall mortality was obvious in the high SDI quintile (Figure 5B). Decomposition analysis in different SDI regions revealed substantial heterogeneity in demographic and epidemiologic trends. Population growth and aging were main drivers of change in morbidity and mortality of hypertension-related CKD in global.

**Figure 5:**
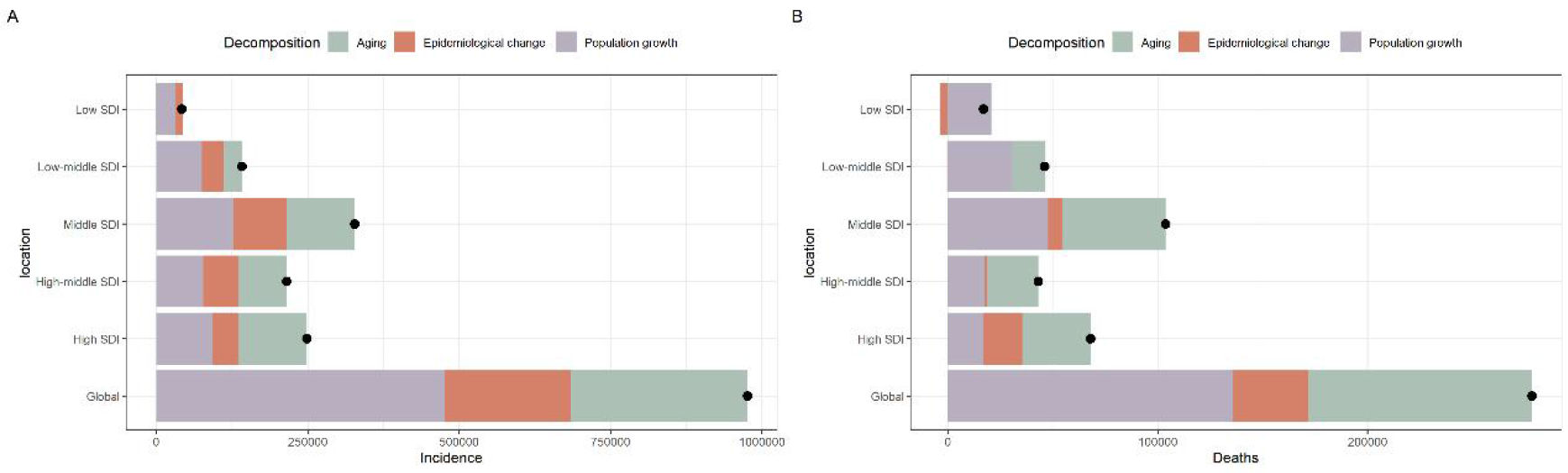
Changes in hypertension-related CKD incidence (A) and deaths (B) according to population-level determinants of population growth, aging, and epidemiological change from 1990 to 2019 at the global level and by socio-demographic index (SDI) regions. Black dots represent the overall value of change contributed by all three components.

### Burden of hypertension-related CKD by age group

In 2019, the changes in global incidence rates by age were similar in both sexes, remaining steady growth and reaching two peaks in the age range of 80-84 years (Figure 6A and 6B). Incidence rates increased by more than 10 (per 100,000 population) in the 60-64 years, 65-69 years, 70-74 years, 75-79 years, 80-84 years, and 85-89 years age groups and increased by more than 5 (per 100,000 population) in the 55-59 years age group from 1990 to 2019. Global DALY rates tended to increase with age in 2019, and the disease burden of CKD increased in the older population compared to 1990 (Figure 6C and 6D).

**Figure 6:**
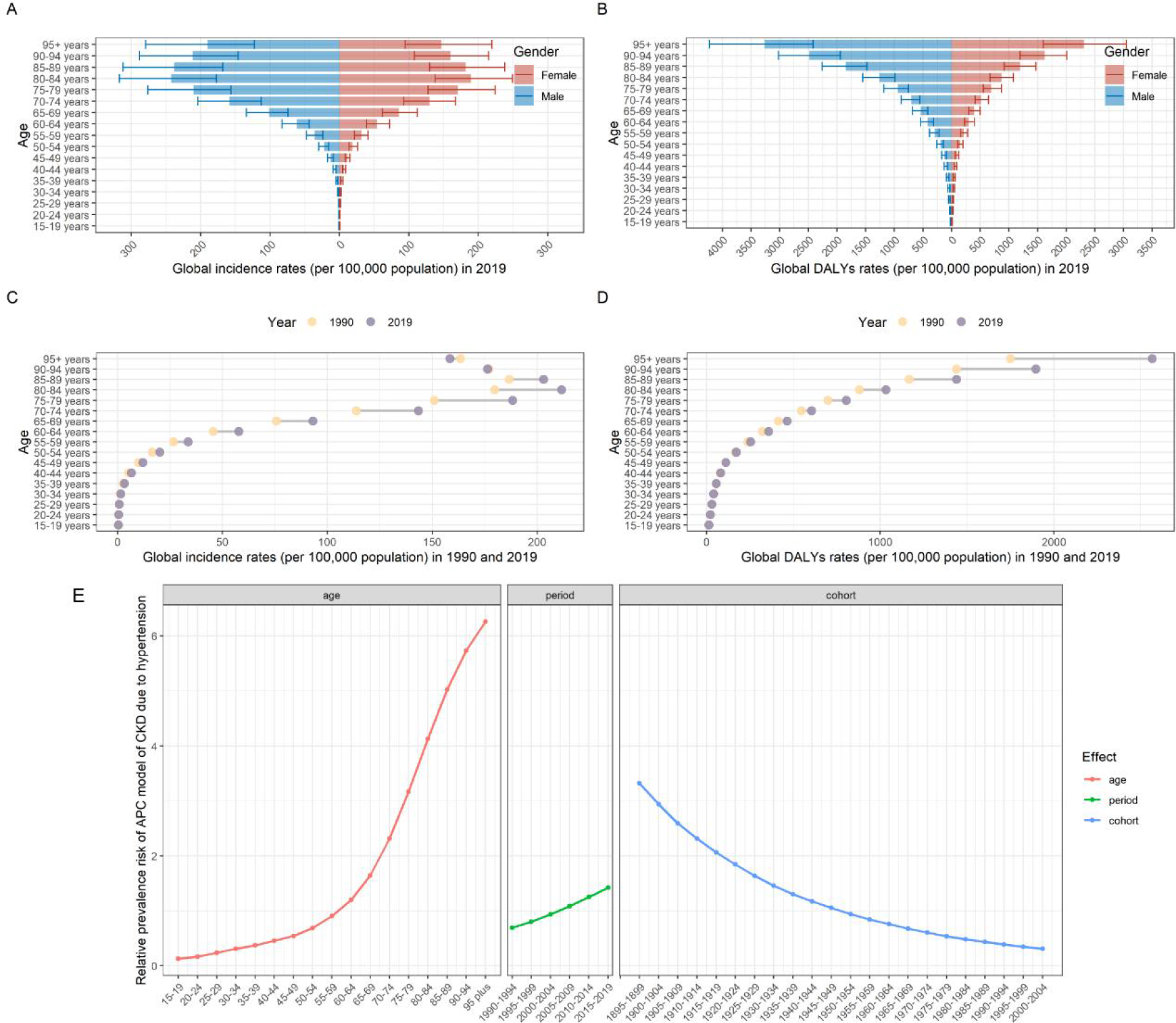
Global incidence rates (A) and disability-adjusted life years (DALYs) rates (B) for male and female by age in 2019. Global incidence rates (C) and disability-adjusted life years (DALYs) rates (D) for both sex by age in 1990 and 2019. Age, period and cohort effect relative risk of liver cancer prevalence in global, 1990-2019 (E).

**Figure 7:**
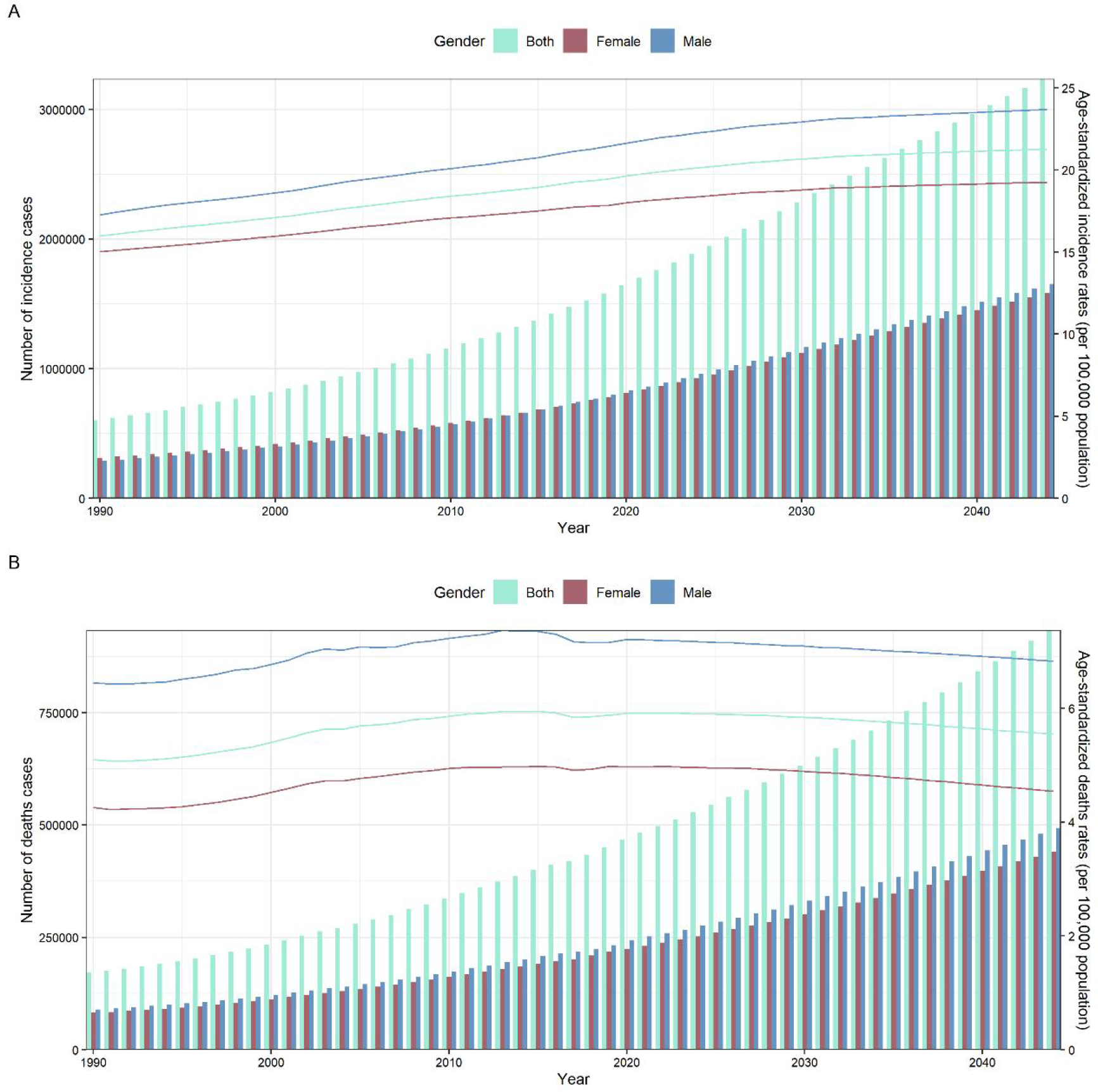
Incidence (A) and deaths (B) trends of hypertension-related CKD from 1990 to 2019 and projections from 2020 to 2044.

The APC-IE method presented estimated coefficients for the age, period, and cohort effects, and then these coefficients were calculated to the exponential value (exp(coef.) = e^coef.^) which denotes the prevalence relative risk (RR) of a particular age, period, or birth cohort relative to each average level (Supplementary Table S11 and Figure S2)[10,11]. Figure 6E was also plotted to reflect the age, period, and cohort effect trends of RR. After controlling for period and cohort effects, the age effect on hypertension-related CKD showed that the prevalence risk significantly increased with advancing age. From the age groups 15-19 to 80-84 years, the prevalence RR increased by 31.76 times. After controlling for cohort and age effects, the period effect on prevalence presented moderately increasing trends from 1990 to 2019, and the prevalence RR increased from 0.69 in 1990-1995 to 1.42 in 2015-2019. The cohort effect showed that the prevalence risk continuously decreased from the earlier birth cohort to the later birth cohort after correcting for age and period effects. From the 1895-1899 to 2000-2004 birth cohorts, the RR of hypertension-related CKD incidence significantly decreased from 3.32 to 0.31.

### Predictions of hypertension-related CKD

The Nordpred APC model has previously been shown to provide excellent estimates of future chronic disease burden, which was chosen to predict morbidity and mortality of hypertension-related CKD. We converted the Norpred model forecasts for each 5-year period into forecasts for each year, as described in previous studies and in general based on a linear relationship. With the Nordpred APC model and hypertension-related CKD data derived from the GBD website, the morbidity and mortality of hypertension-related CKD in the next 25 years were projected.

The ASIR for both genders would increase from 19.45 (per 100,000 population) in 2019 to 21.28 (per 100,000 population) in 2044. The number of new cases would increase from 798,860 (ASIR: 21.43 per 100,000 population) in 2019 to 1,653,734 (ASIR: 23.69 per 100,000 population) in 2044 among males, and the number of new cases would increase from 779,982 (ASIR: 17.84 per 100,000 population) to 1,582,026 (ASIR: 19.26 per 100,000 population) among females during the same period. Between 2019 and 2022, the ASMR for both sexes would increase from 5.88 (per 100,000 population) to 5.92 (per 100,000 population), and then the ASMR would decrease to 5.55 (per 100,000 population) in 2044. However, in the next 25 years, the number of deaths due to hypertensive nephropathy will increase from 232,212 (ASIR: 7.16 per 100,000 population) to 493,059 (ASIR: 6.83 per 100,000 population) in males and from 217,936 (ASIR: 4.98 per 100,000 population) to 440,435 (ASIR: 4.54 per 100,000 population) in females.

## Discussion

In this study, we showed that the incidence, prevalence, DALYs, and deaths from hypertension-related CKD increased from 1990 to 2019. The prevalence of CKD is higher in women than in men[12]. For hypertension-related CKD, which was the focus of our study, although the prevalence was slightly higher in women than in men, more new cases were reported in males than in females since 2014 (Figure 1). Previous studies reported that hypertension was the second greatest risk factor for CKD after diabetes [13], contributing to 26.6% of CKD ASDR globally, and accounted for the largest proportion of CKD burden in East Asia, Eastern Europe, Tropical Latin America, and Western sub-Saharan Africa[1]. Our investigation identified regions with a large burden of hypertension-related CKD as well as regions where this disease was increasing significantly. We found that East Asia had the largest count of DALYs, and Central Latin America ranked first in ASDR (279.03 per 100,000 population; 95% UI: 228.09 to 338.68). Additionally, the most pronounced ASDR increase occurred in Central Latin America (EAPC=2.47; 95% CI: 2.14 to 2.80).

The Wilcoxon signed-rank test revealed that the ASIR and ASMR of hypertension-related CKD were significantly different among sexes in all SDI regions (p < 0.05) (Figure 4A and 4D), which was consistent with prior findings that the prevalence of hypertension was slightly higher in men than in women[3] and that men were more likely to suffer kidney failure[14]. Based on the decomposition analysis and the age effect in the APC-IE model, we found that the increase in the incidence of hypertension-related CKD was strongly associated with advancing age. This finding was also reported by Chen et al.[15] that males and elderly people tended to have a higher disease burden of hypertension-related CKD. However, this did not mean that the burden of CKD in the young and middle-aged population could be ignored. The global incidence rate increased by more than 5 (per 100,000 population) in the 55-59 years age group from 1990 to 2019. The age group with the highest incidence of CKD was 85-89 years in 1990, while in 2019, it changed to 80-84 years (Figure 6C), which indicated a slightly earlier peak in incidence.

Because of its extremely high prevalence in older adults, hypertension is not only a leading cause of preventable morbidity and mortality but, perhaps more importantly, is underrecognized as a major contributor to premature disability and institutionalization[16]. According to GBD 2019, high systolic blood pressure increased the risk of 18 diseases, which we grouped by different regions and ranked from highest to lowest of these diseases burden increased by hypertension, using all age DALY rates as the reference standard (Supplementary Figure S3). The burden of hypertension-related CKD was ranked fifth in global and all SDI regions (Supplementary Figure S4) and second in Andean Latin America and Central Latin America. We have not seen the same degree of progress in preventing mortality from hypertension-related CKD as we have in hypertensive cardiac damage and brain damage. From 1990 to 2019, the global ASMR from hypertensive heart disease, ischemic heart disease, and stroke attributed to high systolic blood pressure decreased by 21.49% (95% UI: 10.13% to 35.18%), 32.64% (95% UI: 28.74% to 36.81%) and 34.89% (95% UI: 28.69% to 40.87%), respectively, but a similar decrease was not observed for hypertension-related CKD, which actually increased by 17.56% (95% UI: 7.59% to 24.87%).

According to the predictions of our study, the number of new cases and deaths of hypertension-related CKD would be expected to continue to increase globally during the next 25 years due to population growth, aging, and the increase in the incidence rate. In addition to the chronic course of kidney damage caused by hypertension, which leads to a lack of awareness of hypertension-related CKD, the following reasons may contribute to the increased burden of this disease. Current studies suggest that intensive antihypertensive therapy may benefit patients by reducing their major adverse cardiovascular events[17,18], while few studies have focused on protecting renal function in hypertensive patients. According to a meta-analysis, blood pressure lowering treatment significantly reduced the risk of cardiovascular disease and death in various populations of patients; however, there was a lack of overall benefit of blood pressure lowering for renal failure events[19]. Although cardiovascular disease is the leading cause of death in CKD[20], should more emphasis be placed on the protection of renal function in the early stages of hypertension-related CKD? In which populations? How can kidney function be better protected? These are questions that cannot be avoided to achieve precision medicine care for hypertension-related CKD. The association between cardiovascular disease and CKD also needs more research. The ASMR of hypertension-related CKD was associated with regional development status and medical level, which was related to the management and control of hypertension, in addition to the fact that dialysis can prolong the life of patients[21]; the ASIR of low and low-middle SDI regions were not significant; however, they had higher ASDR than other regions.

There are several limitations in the present study. In some cases of concomitant hypertension and CKD, the sequence of events (i.e., which came first, CKD or hypertension) cannot be established[22], and most of the data sources were cross-sectional studies that only measured glomerular filtration rate once. These heterogeneous databases inevitably increase the uncertainty of the analyzed data. In addition, stratified analysis by CKD stage (i.e., analysis of disease severity) would be meaningful, but relevant data are not currently available.

## Conclusions

This study estimated the temporal trends of hypertension-related CKD morbidity and mortality globally from 1990 to 2019. We found that the burden of hypertension-related CKD had increased significantly over the past 30 years, and the upward trend of incidence is expected to continue in the next 25 years. Consequently, effective strategies and increased awareness are essential to improve the current status of hypertension-related CKD, thereby improving the quality of life in patients and reducing avoidable deaths.

## Data Availability

The data used for these analyses are all publicly available at the online GBD repository (http://ghdx.healthdata.org/gbd-resultstool).

## Author contributions

HZ and YR conceived the study. YR accessed and acquired the raw data, performed the data analysis, prepared tables and figures, and wrote the manuscript. HZ reviewed the manuscript.

## Acknowledgments

We appreciate the work of the Global Burden of Disease study 2019 collaborators.

## Funding

This work was supported by the CAMS Initiative for Innovative Medicine (CAMS-I2M) [grant number 2016-I2M-3-006].

## Conflict of interest

The authors declare that the research was conducted in the absence of any commercial or financial relationships that could be construed as a potential conflict of interest.

## Notes

### Competing Interest Statement

The authors have declared no competing interest.

