## Supplementary for "The trend of hypertension-related chronic kidney disease from 1990 to 2019 and its predictions over 25 years: An analysis of the Global Burden of Disease Study 2019"

Table S1. The incident count and ASIR of CKD due to hypertension in 1990 and 2019 for both sexes and all locations, and its temporal trends from 1990 to 2019.

| Incidence | 1990 | | 2019 | | 1990-2019 |
|---|---|---|---|---|---|
| location | Incidence count (95% UI) | ASIR per 100,000 (95% UI) | Incidence count (95% UI) | ASIR per 100,000 (95% UI) | EAPC in ASIR (95% CI) |
| Global | 602690 (548661 to 659099) | 15.97 (14.6 to 17.4) | 1578842 (1447111 to 1715250) | 19.45 (17.85 to 21.09) | 0.69 (0.68 to 0.7) |
| Low SDI | 22089 (20011 to 24354) | 9.73 (8.9 to 10.62) | 64070 (58170 to 70185) | 12.82 (11.64 to 14.03) | 0.98 (0.95 to 1.02) |
| Low-middle SDI | 68675 (61978 to 75451) | 11.69 (10.62 to 12.85) | 210408 (190971 to 231092) | 15.69 (14.28 to 17.17) | 0.95 (0.91 to 0.99) |
| Middle SDI | 134769 (121248 to 149134) | 13.56 (12.3 to 14.95) | 462464 (422782 to 504623) | 18.85 (17.26 to 20.52) | 1.22 (1.19 to 1.25) |
| High-middle SDI | 140355 (127379 to 154411) | 13.65 (12.43 to 14.94) | 355810 (324230 to 387249) | 17.5 (15.97 to 19.03) | 0.94 (0.92 to 0.97) |
| High SDI | 236512 (215706 to 259152) | 22.08 (20.29 to 24.01) | 485205 (447563 to 524053) | 24.96 (23.02 to 27.05) | 0.33 (0.29 to 0.37) |
| Central Asia | 3935 (3493 to 4465) | 8.15 (7.26 to 9.14) | 10442 (9247 to 11830) | 13.38 (11.99 to 14.99) | 1.87 (1.68 to 2.05) |
| Central Europe | 16480 (14750 to 18426) | 11.28 (10.16 to 12.49) | 38276 (34276 to 42213) | 17.8 (16.13 to 19.58) | 1.35 (1.25 to 1.45) |
| Eastern Europe | 23667 (21161 to 26528) | 8.84 (7.99 to 9.84) | 41688 (37491 to 46592) | 12.66 (11.43 to 14.07) | 1.36 (1.17 to 1.56) |
| Australasia | 5584 (5153 to 6042) | 23 (21.35 to 24.68) | 13518 (12299 to 14738) | 26.23 (23.91 to 28.56) | 0.43 (0.39 to 0.48) |
| High-income Asia Pacific | 46095 (42163 to 50510) | 23.25 (21.38 to 25.29) | 119281 (109155 to 129952) | 25.23 (23.13 to 27.42) | 0.18 (0.13 to 0.22) |

| | | | | | |
|---|---|---|---|---|---|
| High-income North America | 92506 (83857 to 101842) | 25.46 (23.25 to 27.86) | 173525 (158854 to 189169) | 27.26 (25 to 29.56) | 0.12 (0.06 to 0.19) |
| Western Europe | 119480 (107931 to 131484) | 19.57 (17.83 to 21.33) | 211707 (193537 to 230024) | 21.73 (19.9 to 23.55) | 0.33 (0.28 to 0.39) |
| Andean Latin America | 2463 (2227 to 2741) | 12.41 (11.24 to 13.73) | 13021 (11778 to 14313) | 23.54 (21.28 to 25.83) | 2.26 (2.15 to 2.37) |
| Caribbean | 3219 (2897 to 3569) | 12.33 (11.15 to 13.66) | 10244 (9353 to 11180) | 19.78 (18.06 to 21.6) | 1.63 (1.53 to 1.73) |
| Central Latin America | 18008 (16263 to 19978) | 21.15 (19.24 to 23.43) | 74471 (69268 to 80303) | 31.14 (28.98 to 33.58) | 1.28 (1.21 to 1.35) |
| Southern Latin America | 8371 (7420 to 9287) | 18.55 (16.59 to 20.46) | 21209 (19295 to 23155) | 24.97 (22.76 to 27.2) | 1.05 (0.97 to 1.13) |
| Tropical Latin America | 13574 (12279 to 14934) | 14.97 (13.57 to 16.39) | 48423 (44506 to 52900) | 20.09 (18.42 to 22.03) | 0.97 (0.9 to 1.04) |
| North Africa and Middle East | 34923 (31561 to 38611) | 21.13 (19.25 to 23.14) | 155753 (142428 to 169332) | 36.55 (33.59 to 39.58) | 2 (1.9 to 2.1) |
| Southeast Asia | 28971 (26250 to 31934) | 11.51 (10.49 to 12.65) | 105544 (96169 to 115268) | 17.5 (15.98 to 19.09) | 1.47 (1.43 to 1.5) |
| South Asia | 65965 (59154 to 72938) | 11.64 (10.53 to 12.8) | 201541 (180807 to 223001) | 14.59 (13.18 to 16.01) | 0.66 (0.59 to 0.74) |
| East Asia | 99677 (88818 to 111563) | 12.19 (10.94 to 13.56) | 283695 (255672 to 312985) | 14.11 (12.82 to 15.54) | 0.74 (0.65 to 0.82) |
| Oceania | 276 (246 to 313) | 9.28 (8.4 to 10.36) | 854 (755 to 968) | 12.29 (11.05 to 13.7) | 0.88 (0.83 to 0.94) |
| Central Sub-Saharan Africa | 1543 (1364 to 1737) | 7.36 (6.68 to 8.09) | 4908 (4343 to 5485) | 9.97 (8.98 to 11.02) | 1.12 (1.02 to 1.22) |

| | | | | | |
|---|---|---|---|---|---|
| Eastern Sub-Saharan Africa | 5320<br>(4797 to 5853) | 7.62<br>(6.94 to 8.29) | 14716<br>(13293 to 16205) | 9.69<br>(8.78 to 10.62) | 0.86<br>(0.79 to 0.93) |
| Southern Sub-Saharan Africa | 3647<br>(3272 to 4016) | 13.5<br>(12.22 to 14.89) | 10247<br>(9311 to 11254) | 18.54<br>(16.85 to 20.33) | 1.05<br>(0.86 to 1.24) |
| Western Sub-Saharan Africa | 8983<br>(8188 to 9841) | 10.52<br>(9.65 to 11.5) | 25778<br>(23525 to 28219) | 14.11<br>(12.87 to 15.43) | 1.16<br>(1.06 to 1.27) |

Table S2. The prevalence count and ASPR of CKD due to hypertension in 1990 and 2019 for both sexes and all locations, and its temporal trends from 1990 to 2019

| Prevalence | 1990 | | 2019 | | 1990-2019 |
|---|---|---|---|---|---|
| location | Prevalence count (95% UI) | ASPR per 100,000 (95% UI) | Prevalence count (95% UI) | ASPR per 100,000 (95% UI) | EAPC in ASPR (95% CI) |
| Global | 12423987 (11534930 to 13371365) | 332.52 (308.9 to 358.71) | 31601781 (29348813 to 33945841) | 397.32 (369.52 to 426.45) | 0.68 (0.65 to 0.7) |
| Low SDI | 518257 (476887 to 560140) | 219.91 (203.43 to 237.83) | 1461943 (1350267 to 1572973) | 280.2 (259.47 to 303.25) | 0.91 (0.88 to 0.94) |
| Low-middle SDI | 1664143 (1534322 to 1792105) | 280.79 (260.08 to 303.04) | 4657869 (4323164 to 5000905) | 352.9 (327.45 to 379.37) | 0.75 (0.71 to 0.8) |
| Middle SDI | 3192727 (2950528 to 3447792) | 316.79 (292.79 to 342.46) | 9794478 (9052719 to 10543624) | 416.75 (385.88 to 448.85) | 1.12 (1.07 to 1.18) |
| High-middle SDI | 3052059 (2833584 to 3299050) | 302.11 (280.52 to 325.39) | 7195598 (6684084 to 7746626) | 365.02 (339.43 to 392.31) | 0.83 (0.78 to 0.88) |
| High SDI | 3990531 (3674402 to 4341758) | 382 (352.87 to 414.23) | 8473251 (7853689 to 9159927) | 428.7 (397.94 to 461.2) | 0.32 (0.27 to 0.36) |
| Central Asia | 126457 (114879 to 139369) | 260.25 (237.65 to 284.45) | 280488 (255456 to 307689) | 376.93 (345.25 to 410.66) | 1.42 (1.27 to 1.57) |
| Central Europe | 366533 (336052 to 399779) | 262.03 (240.6 to 285.97) | 768862 (706725 to 839307) | 364.9 (336 to 396.94) | 1.06 (1.01 to 1.12) |
| Eastern Europe | 745854 (686384 to 808438) | 283.36 (261.08 to 306.79) | 1205330 (1105073 to 1312526) | 370.64 (340.32 to 402.12) | 1.08 (0.96 to 1.2) |
| Australasia | 83247 (77778 to 88981) | 360.4 (337.08 to 385.11) | 212544 (195414 to 231697) | 406.43 (373.81 to 443.46) | 0.4 (0.36 to 0.44) |
| High-income Asia Pacific | 796918 (737733 to 860900) | 419.1 (389.25 to 452.53) | 2140770 (1975069 to 2328203) | 436.08 (405.09 to 471.98) | 0.12 (0.09 to 0.15) |

| | | | | | |
|---|---|---|---|---|---|
| High-income North America | 1647491 (1502324 to 1818427) | 455.54 (417.53 to 500.86) | 3115516 (2865186 to 3393252) | 488.93 (449.58 to 531.23) | 0.12 (0.03 to 0.22) |
| Western Europe | 1801925 (1643119 to 1976881) | 307.14 (281.79 to 335.48) | 3365585 (3093337 to 3650520) | 336.6 (311.37 to 363.06) | 0.28 (0.24 to 0.33) |
| Andean Latin America | 55208 (49435 to 62040) | 265.87 (240.06 to 295.52) | 257942 (235153 to 282627) | 465.43 (424.46 to 508.86) | 1.99 (1.9 to 2.08) |
| Caribbean | 79280 (72534 to 86531) | 301.48 (275.95 to 328.71) | 235671 (216235 to 255972) | 457.6 (419.97 to 496.94) | 1.46 (1.37 to 1.55) |
| Central Latin America | 409583 (375043 to 448208) | 483.46 (442.23 to 531.41) | 1653256 (1534993 to 1781947) | 705.04 (653.68 to 760.29) | 1.25 (1.18 to 1.32) |
| Southern Latin America | 139607 (126315 to 153200) | 319.22 (289.62 to 350.13) | 348608 (318448 to 379025) | 413.43 (378.62 to 449.12) | 0.96 (0.87 to 1.04) |
| Tropical Latin America | 287222 (263062 to 311532) | 321.16 (294.31 to 348.02) | 969173 (894763 to 1050517) | 409.5 (378.35 to 443.76) | 0.78 (0.71 to 0.84) |
| North Africa and Middle East | 650430 (598583 to 707930) | 406.71 (374.55 to 443.73) | 2782706 (2571747 to 3015285) | 698.77 (643.82 to 756.88) | 1.98 (1.88 to 2.08) |
| Southeast Asia | 805639 (743317 to 871366) | 303.62 (281.32 to 327.35) | 2542556 (2355929 to 2734205) | 434.02 (402.64 to 466.81) | 1.28 (1.24 to 1.31) |
| South Asia | 1624825 (1497599 to 1759068) | 286.8 (265.6 to 310.46) | 4531918 (4210218 to 4882296) | 333.08 (309.11 to 358.99) | 0.47 (0.39 to 0.55) |
| East Asia | 2355590 (2174274 to 2551777) | 282.76 (260.65 to 305.94) | 5914318 (5489853 to 6385094) | 307.77 (285.54 to 332.4) | 0.69 (0.57 to 0.81) |
| Oceania | 8698 (7702 to 9872) | 269.98 (244.74 to 299.92) | 25422 (22470 to 28928) | 342.62 (310.6 to 377.81) | 0.76 (0.72 to 0.79) |
| Central Sub-Saharan Africa | 36641 (32032 to 42682) | 157.07 (142.31 to 174.95) | 114287 (100653 to 131884) | 208.45 (188.72 to 232.08) | 1.02 (0.93 to 1.12) |

| | | | | | |
|---|---|---|---|---|---|
| Eastern Sub-Saharan Africa | 129194 (118045 to 140336) | 164.5 (151.57 to 177.87) | 362386 (330945 to 394212) | 209.65 (193.05 to 227.07) | 0.89 (0.82 to 0.96) |
| Southern Sub-Saharan Africa | 77933 (71508 to 84722) | 278.49 (256.48 to 302.82) | 206118 (189960 to 223779) | 375.96 (347.29 to 406.62) | 0.99 (0.8 to 1.18) |
| Western Sub-Saharan Africa | 195712 (180274 to 210594) | 220.84 (203.68 to 238.81) | 568323 (523342 to 610616) | 295.01 (272.29 to 319.58) | 1.12 (1.04 to 1.21) |

Table S3.The DALYs count and ASDR of CKD due to hypertension in 1990 and 2019 for both sexes and all locations, and its temporal trends from 1990 to 2019.

| DALYs | 1990 | | 2019 | | 1990-2019 |
|---|---|---|---|---|---|
| location | DALYs count (95% UI) | ASDR (95% UI) | DALYs count (95% UI) | ASDR (95% UI) | EAPC in ASDR (95% CI) |
| Global | 4424386 (3817916 to 5211065) | 111.27 (96.36 to 129.18) | 9962410 (8582331 to 11544122) | 123.41 (106.86 to 142.66) | 0.49 (0.43 to 0.55) |
| Low SDI | 444426 (370274 to 537075) | 187 (156.48 to 222.31) | 892050 (742774 to 1068895) | 166.36 (141.42 to 194.38) | -0.41 (-0.47 to -0.34) |
| Low-middle SDI | 902748 (753866 to 1084681) | 145.11 (122.17 to 171.77) | 2025514 (1698389 to 2428570) | 146.64 (124.1 to 172.77) | 0.08 (-0.02 to 0.18) |
| Middle SDI | 1584836 (1352882 to 1886236) | 149.45 (128.64 to 174.94) | 3880511 (3312419 to 4538069) | 159.82 (137.32 to 185.98) | 0.41 (0.35 to 0.47) |
| High-middle SDI | 824558 (707656 to 956778) | 79.72 (68.57 to 91.85) | 1569896 (1347109 to 1814174) | 79.78 (68.43 to 92.04) | 0.15 (0.05 to 0.26) |
| High SDI | 665072 (571560 to 764597) | 64.23 (55.48 to 73.54) | 1587608 (1343430 to 1830593) | 81.57 (69.66 to 93.81) | 0.98 (0.93 to 1.04) |
| Central Asia | 16977 (13673 to 21200) | 33.22 (26.79 to 41.06) | 37385 (29660 to 46519) | 48.82 (39.17 to 60.3) | 1.15 (0.88 to 1.41) |
| Central Europe | 75668 (63969 to 89214) | 52.98 (45.02 to 61.75) | 109060 (88669 to 131707) | 52.2 (42.7 to 62.85) | 0.41 (0.22 to 0.61) |
| Eastern Europe | 75293 (62786 to 90072) | 28.9 (24.24 to 34.48) | 105598 (86489 to 126472) | 33.45 (27.52 to 39.93) | 0.11 (-0.28 to 0.5) |
| Australasia | 7478 (6472 to 8639) | 33.47 (28.7 to 38.96) | 23087 (18113 to 28848) | 42.98 (34.12 to 53.32) | 0.9 (0.87 to 1.02) |

| | | | | | |
|---|---|---|---|---|---|
| High-income Asia Pacific | 138646 (116898 to 164289) | 73.48 (61.86 to 86.4) | 274538 (222050 to 325496) | 54.81 (45.17 to 64.2) | -1.37 (-1.65 to -1.09) |
| High-income North America | 282204 (242247 to 321798) | 79.05 (67.92 to 89.96) | 762709 (649818 to 878231) | 120.6 (102.68 to 138.93) | 1.72 (1.57 to 1.87) |
| Western Europe | 270282 (223715 to 318910) | 46.44 (38.85 to 54.52) | 514454 (419715 to 616051) | 48.9 (40.22 to 58.16) | 0.46 (0.35 to 0.57) |
| Andean Latin America | 27203 (22453 to 32962) | 133.82 (110.76 to 161.79) | 103597 (82360 to 129266) | 187.72 (148.51 to 235.05) | 1.39 (1.09 to 1.69) |
| Caribbean | 28031 (23070 to 33644) | 106.45 (88.04 to 127.54) | 76529 (62122 to 93596) | 148.44 (120.36 to 182.43) | 1.57 (1.44 to 1.69) |
| Central Latin America | 125992 (106367 to 148563) | 147.65 (125.8 to 173.29) | 660896 (539825 to 800499) | 279.03 (228.09 to 338.68) | 2.47 (2.14 to 2.8) |
| Southern Latin America | 59975 (50348 to 71196) | 133.72 (113.12 to 157.76) | 130026 (109456 to 153964) | 154.37 (130.1 to 182.74) | 0.52 (0.28 to 0.76) |
| Tropical Latin America | 98091 (81261 to 116395) | 106.54 (89.66 to 124.98) | 262804 (222572 to 309161) | 109.81 (93.04 to 129.38) | 0.11 (0.02 to 0.2) |
| North Africa and Middle East | 360774 (295600 to 451184) | 222.39 (183.37 to 282.61) | 834868 (674719 to 1006266) | 203.28 (165.77 to 243.04) | -0.31 (-0.38 to -0.25) |
| Southeast Asia | 682481 (574665 to 821797) | 241.26 (206.14 to 286.87) | 1525410 (1288327 to 1809533) | 247.03 (210.3 to 289.49) | 0.14 (0.08 to 0.2) |
| South Asia | 759161 (609946 to 949310) | 130.49 (105.12 to 161.56) | 1772342 (1438401 to 2149967) | 123.65 (100.99 to 149.93) | -0.25 (-0.42 to -0.08) |
| East Asia | 936900 (783805 to 1114965) | 103.7 (87.31 to 121.25) | 1804537 (1492474 to 2140785) | 91.21 (75.93 to 107.27) | 0 (-0.13 to 0.14) |
| Oceania | 5663 (4607 to 6997) | 169.19 (139.69 to 204.6) | 14948 (11672 to 19030) | 192.97 (153.13 to 238.79) | 0.33 (0.16 to 0.5) |

| | | | | | |
|---|---|---|---|---|---|
| Central Sub-Saharan Africa | 47508 (37640 to 59322) | 212.59 (172.94 to 259.99) | 96855 (72849 to 125612) | 181.49 (135.44 to 236.53) | -0.64 (-0.67 to -0.61) |
| Eastern Sub-Saharan Africa | 147284 (119022 to 180757) | 195.37 (159.57 to 235.05) | 265561 (220767 to 318323) | 163.46 (137.32 to 193.14) | -0.76 (-0.85 to -0.67) |
| Southern Sub-Saharan Africa | 54740 (45690 to 66637) | 186.15 (155.52 to 225.69) | 143163 (121473 to 167952) | 250.55 (215.25 to 291.5) | 0.48 (-0.07 to 1.03) |
| Western Sub-Saharan Africa | 224036 (181076 to 274386) | 253.02 (208.97 to 305.25) | 444042 (355795 to 542376) | 225.89 (185.66 to 268.58) | -0.3 (-0.37 to -0.23) |

Table S4. The deaths count and ASMR of CKD due to hypertension in 1990 and 2019 for both sexes and all locations, and its temporal trends from 1990 to 2019.

| Deaths | 1990 | | 2019 | | 1990-2019 |
|---|---|---|---|---|---|
| location | Deaths count (95% UI) | ASMR per 100,000 (95% UI) | Deaths count (95% UI) | ASMR per 100,000 (95% UI) | EAPC in ASMR (95% CI) |
| Global | 171878 (146475 to 201264) | 5.1 (4.35 to 5.92) | 450148 (381995 to 525405) | 5.88 (4.95 to 6.82) | 0.64 (0.57 to 0.72) |
| Low SDI | 16712 (13855 to 20027) | 9.25 (7.65 to 11.22) | 33686 (28186 to 39859) | 8.16 (6.85 to 9.66) | -0.45 (-0.52 to -0.37) |
| Low-middle SDI | 31217 (25926 to 37734) | 6.49 (5.41 to 7.84) | 77408 (64238 to 92260) | 6.6 (5.46 to 7.8) | 0.05 (-0.06 to 0.17) |
| Middle SDI | 56141 (47663 to 66208) | 6.97 (5.91 to 8.19) | 159948 (134649 to 186187) | 7.58 (6.33 to 8.79) | 0.44 (0.38 to 0.5) |
| High-middle SDI | 32934 (27751 to 38719) | 3.73 (3.16 to 4.37) | 76005 (63417 to 88727) | 3.93 (3.27 to 4.57) | 0.32 (0.18 to 0.45) |

| | | | | | |
|---|---|---|---|---|---|
| High SDI | 34768<br>(28884 to 40688) | 3.37<br>(2.81 to 3.92) | 102807<br>(81322 to 121590) | 4.48<br>(3.63 to 5.27) | 1.26<br>(1.15 to 1.37) |
| Central Asia | 432<br>(330 to 571) | 1.01<br>(0.76 to 1.36) | 968<br>(731 to 1276) | 1.69<br>(1.26 to 2.27) | 1.57<br>(1.19 to 1.94) |
| Central Europe | 3079<br>(2554 to 3664) | 2.3<br>(1.92 to 2.73) | 5515<br>(4309 to 6971) | 2.44<br>(1.92 to 3.07) | 0.81<br>(0.53 to 1.08) |
| Eastern Europe | 2200<br>(1805 to 2661) | 0.91<br>(0.75 to 1.1) | 3994<br>(3169 to 4929) | 1.17<br>(0.93 to 1.42) | 0.47<br>(-0.06 to 1) |
| Australasia | 433<br>(355 to 524) | 2.11<br>(1.69 to 2.6) | 1768<br>(1268 to 2379) | 2.96<br>(2.13 to 3.94) | 1.34<br>(1.24 to 1.45) |
| High-income Asia Pacific | 7039<br>(5665 to 8421) | 4.19<br>(3.37 to 5) | 19454<br>(14256 to 24116) | 3.09<br>(2.33 to 3.77) | -1.61<br>(-1.97 to -1.24) |
| High-income North America | 15087<br>(12634 to 17460) | 4.06<br>(3.39 to 4.68) | 46890<br>(37979 to 54924) | 6.6<br>(5.39 to 7.69) | 2.08<br>(1.9 to 2.25) |
| Western Europe | 15831<br>(12592 to 19393) | 2.7<br>(2.16 to 3.29) | 42100<br>(31945 to 52137) | 3.44<br>(2.64 to 4.24) | 1.37<br>(1.18 to 1.56) |
| Andean Latin America | 1247<br>(1019 to 1523) | 7.11<br>(5.8 to 8.67) | 5614<br>(4310 to 7135) | 10.6<br>(8.13 to 13.49) | 1.64<br>(1.3 to 1.98) |
| Caribbean | 1123<br>(910 to 1360) | 4.76<br>(3.87 to 5.73) | 3333<br>(2647 to 4172) | 6.41<br>(5.08 to 8.03) | 1.57<br>(1.41 to 1.73) |
| Central Latin America | 4793<br>(4032 to 5672) | 6.98<br>(5.84 to 8.22) | 28288<br>(22372 to 34718) | 12.51<br>(9.94 to 15.32) | 2.33<br>(2.01 to 2.64) |
| Southern Latin America | 3000<br>(2463 to 3586) | 7.24<br>(5.99 to 8.54) | 8145<br>(6633 to 9692) | 9.43<br>(7.7 to 11.22) | 0.97<br>(0.67 to 1.26) |
| Tropical Latin America | 3671<br>(3052 to 4303) | 5.05<br>(4.2 to 5.91) | 12156<br>(10003 to 14530) | 5.32<br>(4.36 to 6.35) | 0.38<br>(0.31 to 0.46) |

| | | | | | |
|---|---|---|---|---|---|
| North Africa and Middle East | 15619 (12632 to 20398) | 11.94 (9.6 to 15.89) | 36295 (29062 to 44169) | 10.6 (8.53 to 12.82) | -0.39 (-0.48 to -0.31) |
| Southeast Asia | 23342 (19695 to 27704) | 10.61 (9.04 to 12.5) | 58097 (48565 to 68582) | 11.07 (9.29 to 13.06) | 0.18 (0.12 to 0.25) |
| South Asia | 25377 (19845 to 32336) | 5.85 (4.51 to 7.43) | 64240 (50515 to 79707) | 5.34 (4.23 to 6.58) | -0.58 (-0.8 to -0.36) |
| East Asia | 30613 (25123 to 36574) | 4.41 (3.68 to 5.21) | 75016 (60877 to 89893) | 4.22 (3.45 to 5.02) | 0.19 (0.06 to 0.32) |
| Oceania | 168 (135 to 210) | 6.94 (5.7 to 8.46) | 455 (347 to 578) | 8.12 (6.42 to 9.97) | 0.39 (0.23 to 0.56) |
| Central Sub-Saharan Africa | 1775 (1412 to 2220) | 11.02 (8.93 to 13.39) | 3698 (2721 to 4858) | 9.59 (7.04 to 12.34) | -0.59 (-0.62 to -0.55) |
| Eastern Sub-Saharan Africa | 5740 (4609 to 7010) | 9.99 (8.05 to 12.14) | 10714 (8898 to 12802) | 8.84 (7.33 to 10.5) | -0.56 (-0.65 to -0.47) |
| Southern Sub-Saharan Africa | 2100 (1755 to 2568) | 8.79 (7.3 to 10.78) | 5876 (5008 to 6895) | 12.5 (10.57 to 14.7) | 0.76 (0.22 to 1.3) |
| Western Sub-Saharan Africa | 9208 (7610 to 11234) | 13.18 (10.93 to 16.04) | 17533 (14370 to 20924) | 11.86 (9.78 to 13.93) | -0.27 (-0.34 to -0.2) |

Table S5. The incident count and ASIR of CKD due to hypertension in 1990 and 2019 for both sexes in 204 countries, and its temporal trends from 1990 to 2019.

| location | incident count_1990 (95% UI) | ASIR_1990 (95% UI) | incident count_2019 (95% UI) | ASIR_2019 (95% UI) | incident count_change (*100%) (95% CI) | EAPC (95% CI) |
|---|---|---|---|---|---|---|
| Afghanistan | 1336 (1160 to 1540) | 19.44 (17.12 to 21.94) | 3678 (3171 to 4262) | 31.37 (27.72 to 35.4) | 1.75 (1.55 to 1.96) | 1.79 (1.71 to 1.88) |
| Albania | 209 (180 to 243) | 10.14 (8.84 to 11.59) | 706 (619 to 803) | 16.36 (14.33 to 18.57) | 2.38 (2.05 to 2.71) | 1.72 (1.67 to 1.76) |
| Algeria | 2637 (2287 to 3017) | 21.9 (19.51 to 24.63) | 12397 (11030 to 13952) | 37.09 (33.28 to 41.16) | 3.7 (3.45 to 3.98) | 1.86 (1.76 to 1.97) |
| American Samoa | 4 (3 to 5) | 16.72 (14.74 to 19.05) | 11 (10 to 13) | 23.89 (21.15 to 27.12) | 1.95 (1.77 to 2.15) | 1.19 (1.09 to 1.29) |
| Andorra | 11 (9 to 12) | 20.89 (18.64 to 23.63) | 30 (27 to 34) | 21.71 (19.37 to 24.32) | 1.78 (1.6 to 1.98) | 0.13 (0.03 to 0.24) |
| Angola | 248 (212 to 288) | 6.94 (6.15 to 7.76) | 1036 (885 to 1203) | 10.03 (8.83 to 11.37) | 3.18 (3 to 3.37) | 1.39 (1.26 to 1.51) |
| Antigua and Barbuda | 8 (7 to 9) | 15.99 (14.05 to 17.9) | 25 (23 to 29) | 24.7 (22.03 to 27.63) | 2.05 (1.87 to 2.25) | 1.52 (1.4 to 1.65) |
| Argentina | 5910 (5199 to 6635) | 18.68 (16.54 to 20.81) | 13310 (11989 to 14740) | 24.12 (21.72 to 26.73) | 1.25 (1.09 to 1.43) | 0.88 (0.79 to 0.97) |
| Armenia | 193 (164 to 233) | 7.21 (6.18 to 8.49) | 553 (481 to 637) | 13.46 (11.72 to 15.5) | 1.86 (1.58 to 2.15) | 2.37 (2.23 to 2.51) |
| Australia | 4637 (4280 to 5017) | 22.93 (21.23 to 24.68) | 11443 (10364 to 12541) | 26.32 (23.85 to 28.84) | 1.47 (1.32 to 1.63) | 0.44 (0.39 to 0.49) |
| Austria | 2456 (2153 to 2778) | 19.43 (17.26 to 21.71) | 4639 (4139 to 5167) | 24.61 (21.81 to 27.29) | 0.89 (0.76 to 1.05) | 0.77 (0.72 to 0.81) |
| Azerbaijan | 407 (339 to 487) | 7.89 (6.73 to 9.19) | 1406 (1184 to 1646) | 14.29 (12.32 to 16.34) | 2.46 (2.18 to 2.78) | 2.18 (1.95 to 2.41) |

| | | | | | | |
|---|---|---|---|---|---|---|
| Bahamas | 23<br>(20 to 26) | 14.4<br>(12.64 to 16.25) | 84<br>(73 to 95) | 21.09<br>(18.56 to 23.73) | 2.67<br>(2.45 to 2.91) | 1.28<br>(1.13 to 1.42) |
| Bahrain | 43<br>(38 to 50) | 25.47<br>(22.9 to 28.43) | 453<br>(385 to 526) | 42.39<br>(38.27 to 46.81) | 9.43<br>(8.72 to 10.17) | 1.85<br>(1.71 to 1.98) |
| Bangladesh | 3423<br>(2966 to 3944) | 7.63<br>(6.67 to 8.62) | 14883<br>(13001 to 17069) | 11.59<br>(10.2 to 13.22) | 3.35<br>(3.07 to 3.61) | 1.45<br>(1.28 to 1.62) |
| Barbados | 40<br>(35 to 46) | 14.18<br>(12.41 to 16.07) | 106<br>(94 to 119) | 21.79<br>(19.18 to 24.53) | 1.64<br>(1.48 to 1.83) | 1.46<br>(1.33 to 1.58) |
| Belarus | 981<br>(849 to 1141) | 7.81<br>(6.72 to 9.01) | 1596<br>(1389 to 1815) | 10.43<br>(9.02 to 11.93) | 0.63<br>(0.51 to 0.78) | 1.08<br>(0.76 to 1.4) |
| Belgium | 3420<br>(3035 to 3836) | 21.03<br>(18.85 to 23.33) | 5503<br>(5003 to 6068) | 22.84<br>(20.7 to 25.28) | 0.61<br>(0.47 to 0.75) | 0.35<br>(0.28 to 0.41) |
| Belize | 13<br>(11 to 15) | 14.07<br>(12.33 to 16.11) | 63<br>(55 to 72) | 21.95<br>(19.49 to 24.73) | 3.73<br>(3.35 to 4.1) | 1.53<br>(1.46 to 1.6) |
| Benin | 211<br>(186 to 240) | 10.61<br>(9.51 to 11.93) | 678<br>(589 to 777) | 14.12<br>(12.47 to 15.94) | 2.21<br>(2.07 to 2.36) | 1.05<br>(0.97 to 1.12) |
| Bermuda | 9<br>(8 to 10) | 14.23<br>(12.43 to 16.02) | 29<br>(26 to 33) | 23.07<br>(20.36 to 25.88) | 2.28<br>(2.08 to 2.51) | 1.68<br>(1.49 to 1.87) |
| Bhutan | 24<br>(21 to 28) | 9.65<br>(8.52 to 10.95) | 87<br>(76 to 100) | 15.39<br>(13.55 to 17.55) | 2.62<br>(2.4 to 2.87) | 1.8<br>(1.73 to 1.87) |
| Bolivia (Plurinational State of) | 419<br>(369 to 474) | 13.53<br>(12.1 to 15.19) | 1840<br>(1630 to 2046) | 21.4<br>(19.1 to 23.68) | 3.39<br>(3.17 to 3.66) | 1.61<br>(1.57 to 1.66) |
| Bosnia and Herzegovina | 402<br>(352 to 461) | 10.01<br>(8.84 to 11.33) | 1073<br>(942 to 1212) | 17.92<br>(15.86 to 20.09) | 1.67<br>(1.45 to 1.92) | 2.29<br>(2.16 to 2.41) |
| Botswana | 70<br>(60 to 81) | 12.45<br>(10.93 to 14.12) | 257<br>(222 to 301) | 19.11<br>(16.84 to 21.66) | 2.69<br>(2.53 to 2.87) | 1.41<br>(1.25 to 1.58) |

| | | | | | | |
|---|---|---|---|---|---|---|
| Brazil | 13246<br>(11983 to 14582) | 14.98<br>(13.57 to 16.4) | 47102<br>(43268 to 51511) | 20<br>(18.34 to 21.9) | 2.56<br>(2.4 to 2.73) | 0.95<br>(0.89 to 1.02) |
| Brunei Darussalam | 23<br>(20 to 26) | 26.64<br>(23.67 to 29.52) | 76<br>(66 to 88) | 28.8<br>(25.49 to 32.18) | 2.35<br>(2.17 to 2.52) | 0.27<br>(0.15 to 0.38) |
| Bulgaria | 1375<br>(1182 to 1610) | 11.2<br>(9.77 to 12.83) | 2598<br>(2277 to 2966) | 18.34<br>(16.09 to 20.74) | 0.89<br>(0.69 to 1.11) | 1.75<br>(1.69 to 1.8) |
| Burkina Faso | 401<br>(353 to 455) | 9.47<br>(8.51 to 10.56) | 1141<br>(997 to 1306) | 12.91<br>(11.45 to 14.5) | 1.85<br>(1.71 to 2.02) | 1.19<br>(1.09 to 1.29) |
| Burundi | 166<br>(147 to 188) | 7.4<br>(6.64 to 8.21) | 382<br>(333 to 434) | 9.17<br>(8.18 to 10.28) | 1.3<br>(1.17 to 1.43) | 0.88<br>(0.79 to 0.96) |
| Cabo Verde | 21<br>(18 to 24) | 8.83<br>(7.82 to 10) | 58<br>(51 to 67) | 14.2<br>(12.5 to 16.22) | 1.78<br>(1.62 to 1.97) | 1.88<br>(1.75 to 2.01) |
| Cambodia | 379<br>(324 to 438) | 8.57<br>(7.5 to 9.71) | 1574<br>(1363 to 1809) | 13.46<br>(11.91 to 15.35) | 3.16<br>(2.9 to 3.44) | 1.61<br>(1.51 to 1.72) |
| Cameroon | 599<br>(525 to 683) | 13.67<br>(12.24 to 15.24) | 2270<br>(1960 to 2620) | 18.82<br>(16.66 to 21.16) | 2.79<br>(2.61 to 2.97) | 1.22<br>(1.14 to 1.31) |
| Canada | 7245<br>(6418 to 8046) | 21.92<br>(19.48 to 24.17) | 15900<br>(14311 to 17609) | 22.32<br>(20.07 to 24.83) | 1.19<br>(0.99 to 1.39) | 0.26<br>(0.17 to 0.36) |
| Central African Republic | 75<br>(65 to 88) | 6.91<br>(6.2 to 7.74) | 177<br>(152 to 206) | 8.82<br>(7.81 to 9.87) | 1.34<br>(1.24 to 1.44) | 0.94<br>(0.84 to 1.04) |
| Chad | 278<br>(244 to 316) | 9.85<br>(8.79 to 11.01) | 690<br>(598 to 801) | 12.57<br>(11.12 to 14.23) | 1.48<br>(1.35 to 1.61) | 0.91<br>(0.78 to 1.04) |
| Chile | 1775<br>(1568 to 1990) | 18.7<br>(16.68 to 20.9) | 6701<br>(6010 to 7378) | 27.73<br>(24.86 to 30.55) | 2.78<br>(2.51 to 3.08) | 1.43<br>(1.33 to 1.52) |
| China | 94504<br>(84066 to 106106) | 12.01<br>(10.77 to 13.38) | 268862<br>(241803 to 297432) | 13.89<br>(12.59 to 15.31) | 1.84<br>(1.73 to 1.96) | 0.75<br>(0.66 to 0.84) |

| | | | | | | |
|---|---|---|---|---|---|---|
| Colombia | 3065 (2655 to 3518) | 17.41 (15.26 to 19.76) | 13075 (11641 to 14700) | 24.78 (22.05 to 27.93) | 3.27 (2.93 to 3.63) | 1.14 (1.07 to 1.21) |
| Comoros | 17 (15 to 19) | 7.9 (7.1 to 8.78) | 48 (42 to 54) | 10.17 (9.03 to 11.37) | 1.81 (1.67 to 1.97) | 0.99 (0.91 to 1.07) |
| Congo | 81 (69 to 93) | 7.91 (7.06 to 8.83) | 276 (237 to 318) | 11.18 (9.91 to 12.5) | 2.43 (2.28 to 2.58) | 1.4 (1.27 to 1.52) |
| Cook Islands | 2 (2 to 2) | 14.42 (12.76 to 16.2) | 5 (5 to 6) | 21.92 (19.31 to 24.72) | 1.97 (1.8 to 2.16) | 1.35 (1.27 to 1.43) |
| Costa Rica | 594 (541 to 652) | 33.78 (31.06 to 36.7) | 1814 (1662 to 1981) | 34.69 (31.87 to 37.79) | 2.05 (1.99 to 2.12) | 0.1 (0.1 to 0.1) |
| Côte d'Ivoire | 466 (400 to 540) | 12.02 (10.8 to 13.25) | 1565 (1354 to 1812) | 15.15 (13.53 to 16.83) | 2.36 (2.19 to 2.51) | 0.9 (0.81 to 0.99) |
| Croatia | 840 (722 to 956) | 13.19 (11.46 to 14.88) | 1840 (1619 to 2062) | 21.08 (18.62 to 23.68) | 1.19 (0.97 to 1.45) | 1.79 (1.72 to 1.86) |
| Cuba | 1105 (958 to 1265) | 10.8 (9.4 to 12.37) | 3553 (3154 to 3984) | 19.05 (16.89 to 21.48) | 2.22 (1.99 to 2.46) | 1.98 (1.89 to 2.06) |
| Cyprus | 210 (182 to 240) | 24.99 (22.38 to 27.77) | 540 (476 to 610) | 26.42 (23.56 to 29.48) | 1.57 (1.42 to 1.73) | 0.22 (0.12 to 0.32) |
| Czechia | 1559 (1370 to 1777) | 11.39 (10.01 to 12.92) | 3758 (3261 to 4271) | 17.71 (15.59 to 19.99) | 1.41 (1.25 to 1.59) | 1.48 (1.39 to 1.58) |
| Democratic People's Republic of Korea | 1747 (1512 to 1999) | 11.73 (10.27 to 13.24) | 4634 (4047 to 5236) | 14.79 (12.97 to 16.66) | 1.65 (1.49 to 1.86) | 0.87 (0.69 to 1.06) |
| Democratic Republic of the Congo | 1076 (938 to 1229) | 7.37 (6.63 to 8.18) | 3220 (2795 to 3671) | 9.76 (8.64 to 10.88) | 1.99 (1.83 to 2.16) | 1.01 (0.91 to 1.11) |
| Denmark | 1535 (1349 to 1735) | 17.87 (15.86 to 20.07) | 2688 (2397 to 3008) | 22.01 (19.78 to 24.48) | 0.75 (0.61 to 0.91) | 0.83 (0.79 to 0.87) |

| | | | | | | |
|---|---|---|---|---|---|---|
| Djibouti | 10<br>(9 to 12) | 7.96<br>(7.13 to 8.87) | 60<br>(52 to 69) | 10.87<br>(9.79 to 12.17) | 4.86<br>(4.54 to 5.16) | 1.24<br>(1.11 to 1.37) |
| Dominica | 12<br>(11 to 14) | 17.2<br>(15.22 to 19.43) | 21<br>(19 to 24) | 23.6<br>(20.96 to 26.58) | 0.75<br>(0.65 to 0.85) | 1.08<br>(0.93 to 1.23) |
| Dominican Republic | 370<br>(319 to 431) | 9.86<br>(8.62 to 11.27) | 1643<br>(1440 to 1861) | 17.61<br>(15.49 to 19.84) | 3.44<br>(3.17 to 3.74) | 1.8<br>(1.68 to 1.93) |
| Ecuador | 719<br>(628 to 826) | 13.8<br>(12.15 to 15.65) | 4169<br>(3722 to 4654) | 27.66<br>(24.81 to 30.64) | 4.8<br>(4.38 to 5.23) | 2.4<br>(2.26 to 2.53) |
| Egypt | 6205<br>(5348 to 7035) | 22.11<br>(19.48 to 24.64) | 24681<br>(21312 to 27950) | 38.83<br>(34.52 to 43.1) | 2.98<br>(2.81 to 3.15) | 1.94<br>(1.83 to 2.06) |
| El Salvador | 525<br>(455 to 605) | 17.46<br>(15.24 to 20.11) | 1707<br>(1510 to 1942) | 28.99<br>(25.65 to 32.91) | 2.25<br>(2.03 to 2.48) | 1.8<br>(1.73 to 1.87) |
| Equatorial Guinea | 13<br>(11 to 14) | 6.91<br>(6.19 to 7.62) | 57<br>(49 to 66) | 12.44<br>(11.08 to 13.84) | 3.57<br>(3.34 to 3.87) | 2.36<br>(2.22 to 2.51) |
| Eritrea | 61<br>(52 to 72) | 6.76<br>(5.99 to 7.54) | 233<br>(199 to 271) | 9.36<br>(8.28 to 10.52) | 2.81<br>(2.61 to 3.04) | 1.12<br>(1.03 to 1.2) |
| Estonia | 187<br>(162 to 219) | 9.38<br>(8.11 to 10.98) | 379<br>(329 to 435) | 15.51<br>(13.49 to 17.91) | 1.02<br>(0.87 to 1.16) | 1.94<br>(1.75 to 2.12) |
| Eswatini | 42<br>(36 to 49) | 14.37<br>(12.7 to 16.34) | 110<br>(94 to 127) | 18.67<br>(16.48 to 20.99) | 1.62<br>(1.51 to 1.75) | 0.82<br>(0.62 to 1.02) |
| Ethiopia | 1372<br>(1214 to 1530) | 7.49<br>(6.73 to 8.21) | 3706<br>(3310 to 4109) | 9.48<br>(8.46 to 10.49) | 1.7<br>(1.59 to 1.82) | 0.82<br>(0.73 to 0.92) |
| Fiji | 56<br>(48 to 66) | 15.48<br>(13.64 to 17.52) | 150<br>(131 to 173) | 20.26<br>(18.02 to 22.75) | 1.67<br>(1.47 to 1.87) | 0.85<br>(0.81 to 0.88) |
| Finland | 1087<br>(939 to 1250) | 14.7<br>(12.78 to 16.76) | 2331<br>(2079 to 2605) | 17.55<br>(15.7 to 19.63) | 1.14<br>(0.94 to 1.38) | 0.5<br>(0.4 to 0.6) |

| | | | | | | |
|---|---|---|---|---|---|---|
| France | 16361 (14486 to 18419) | 18.29 (16.38 to 20.41) | 30877 (27821 to 34173) | 21.26 (19.05 to 23.6) | 0.89 (0.75 to 1.02) | 0.49 (0.43 to 0.55) |
| Gabon | 50 (44 to 57) | 9.34 (8.39 to 10.41) | 142 (123 to 162) | 14.21 (12.61 to 15.76) | 1.81 (1.67 to 1.94) | 1.55 (1.47 to 1.63) |
| Gambia | 36 (32 to 42) | 10.4 (9.3 to 11.7) | 132 (116 to 153) | 13.86 (12.32 to 15.7) | 2.66 (2.5 to 2.82) | 1.03 (0.96 to 1.09) |
| Georgia | 474 (401 to 557) | 7.84 (6.75 to 9.15) | 710 (623 to 806) | 12.48 (10.78 to 14.28) | 0.5 (0.35 to 0.65) | 1.51 (1.39 to 1.63) |
| Germany | 27662 (24594 to 30900) | 20.86 (18.63 to 23.24) | 48756 (44224 to 53587) | 24.11 (22.04 to 26.29) | 0.76 (0.63 to 0.9) | 0.46 (0.43 to 0.49) |
| Ghana | 560 (481 to 656) | 9.25 (8.17 to 10.44) | 2118 (1813 to 2460) | 13.48 (11.93 to 15.18) | 2.78 (2.6 to 2.96) | 1.32 (1.25 to 1.38) |
| Greece | 3876 (3426 to 4326) | 24.54 (21.76 to 27.24) | 6103 (5453 to 6761) | 24.25 (21.67 to 26.9) | 0.57 (0.48 to 0.67) | 0.04 (-0.08 to 0.16) |
| Greenland | 5 (4 to 6) | 18.05 (16.14 to 20.22) | 13 (12 to 15) | 21.06 (18.99 to 23.5) | 1.67 (1.51 to 1.86) | 0.51 (0.49 to 0.54) |
| Grenada | 11 (10 to 12) | 15.65 (13.87 to 17.69) | 29 (26 to 33) | 25.93 (23.08 to 29.09) | 1.68 (1.51 to 1.89) | 1.64 (1.51 to 1.77) |
| Guam | 11 (9 to 13) | 13.92 (12.19 to 15.81) | 36 (31 to 41) | 18.93 (16.64 to 21.44) | 2.32 (2.09 to 2.56) | 0.89 (0.75 to 1.04) |
| Guatemala | 685 (590 to 799) | 18.7 (16.48 to 21.4) | 3322 (2928 to 3786) | 29.52 (26.13 to 33.24) | 3.85 (3.57 to 4.17) | 1.55 (1.45 to 1.65) |
| Guinea | 340 (300 to 384) | 10.36 (9.25 to 11.56) | 742 (648 to 843) | 13.64 (12.03 to 15.32) | 1.18 (1.08 to 1.3) | 1.01 (0.93 to 1.09) |
| Guinea-Bissau | 45 (39 to 52) | 11.02 (9.86 to 12.38) | 98 (83 to 116) | 13.46 (11.93 to 15.28) | 1.18 (1.07 to 1.29) | 0.78 (0.67 to 0.88) |

| | | | | | | |
|---|---|---|---|---|---|---|
| Guyana | 52<br>(45 to 60) | 13.19<br>(11.69 to 14.84) | 137<br>(119 to 156) | 21.33<br>(18.91 to 23.89) | 1.65<br>(1.5 to 1.82) | 1.48<br>(1.4 to 1.56) |
| Haiti | 380<br>(325 to 441) | 11.69<br>(10.23 to 13.27) | 1197<br>(1033 to 1376) | 16.99<br>(14.98 to 19.21) | 2.15<br>(2.02 to 2.29) | 1.29<br>(1.26 to 1.31) |
| Honduras | 364<br>(314 to 428) | 17.31<br>(15.19 to 20.06) | 1550<br>(1348 to 1803) | 25<br>(22.05 to 28.62) | 3.25<br>(3.04 to 3.48) | 1.24<br>(1.22 to 1.27) |
| Hungary | 1426<br>(1243 to 1625) | 9.77<br>(8.56 to 11.11) | 3501<br>(3078 to 3978) | 18.06<br>(15.94 to 20.41) | 1.45<br>(1.3 to 1.62) | 2.07<br>(1.98 to 2.17) |
| Iceland | 50<br>(45 to 56) | 16.85<br>(15.04 to 18.92) | 105<br>(93 to 117) | 18.25<br>(16.21 to 20.49) | 1.1<br>(0.96 to 1.26) | 0.1<br>(-0.03 to 0.22) |
| India | 55197<br>(49426 to 61289) | 12.14<br>(11 to 13.36) | 166500<br>(149302 to 183791) | 14.85<br>(13.42 to 16.33) | 2.02<br>(1.89 to 2.15) | 0.56<br>(0.47 to 0.66) |
| Indonesia | 8929<br>(7986 to 9917) | 9.24<br>(8.34 to 10.22) | 28163<br>(25362 to 31276) | 13.26<br>(11.96 to 14.66) | 2.15<br>(2.07 to 2.24) | 1.17<br>(1.09 to 1.26) |
| Iran (Islamic Republic of) | 6456<br>(5712 to 7268) | 24.85<br>(22.55 to 27.39) | 24228<br>(22158 to 26358) | 33.66<br>(30.87 to 36.58) | 2.75<br>(2.54 to 2.96) | 1.1<br>(1.01 to 1.18) |
| Iraq | 1824<br>(1616 to 2054) | 23.82<br>(21.23 to 26.73) | 8903<br>(7914 to 10093) | 38.71<br>(35.07 to 42.91) | 3.88<br>(3.69 to 4.08) | 1.86<br>(1.8 to 1.92) |
| Ireland | 1010<br>(910 to 1104) | 23.48<br>(21.34 to 25.44) | 1723<br>(1627 to 1815) | 22.56<br>(21.27 to 23.79) | 0.71<br>(0.58 to 0.85) | -0.06<br>(-0.1 to -0.03) |
| Israel | 1241<br>(1092 to 1390) | 24.62<br>(21.97 to 27.34) | 3177<br>(2849 to 3533) | 26.98<br>(24.23 to 30.02) | 1.56<br>(1.38 to 1.74) | 0.32<br>(0.22 to 0.43) |
| Italy | 17786<br>(15856 to 19969) | 19.21<br>(17.28 to 21.3) | 32250<br>(29179 to 35342) | 20.41<br>(18.48 to 22.55) | 0.81<br>(0.71 to 0.9) | 0.2<br>(0.15 to 0.25) |
| Jamaica | 267<br>(235 to 302) | 15.1<br>(13.27 to 17.16) | 615<br>(542 to 697) | 20.62<br>(18.2 to 23.39) | 1.3<br>(1.17 to 1.44) | 1.14<br>(0.96 to 1.33) |

| | | | | | | |
|---|---|---|---|---|---|---|
| Japan | 40707<br>(37196 to 44632) | 24.02<br>(22.08 to 26.16) | 97863<br>(89097 to 107229) | 26.43<br>(24.24 to 28.84) | 1.4<br>(1.3 to 1.51) | 0.19<br>(0.14 to 0.24) |
| Jordan | 306<br>(268 to 349) | 23.61<br>(21.1 to 26.2) | 2678<br>(2368 to 3034) | 40.13<br>(36.14 to 44.36) | 7.75<br>(7.23 to 8.26) | 1.99<br>(1.9 to 2.08) |
| Kazakhstan | 951<br>(799 to 1129) | 7.37<br>(6.28 to 8.62) | 2280<br>(1950 to 2665) | 12.85<br>(11.1 to 14.78) | 1.4<br>(1.25 to 1.55) | 2.27<br>(2 to 2.54) |
| Kenya | 591<br>(533 to 651) | 7.61<br>(6.9 to 8.39) | 1970<br>(1766 to 2190) | 9.57<br>(8.63 to 10.63) | 2.34<br>(2.27 to 2.4) | 0.75<br>(0.65 to 0.84) |
| Kiribati | 4<br>(3 to 5) | 10.93<br>(9.46 to 12.59) | 11<br>(9 to 13) | 15.31<br>(13.39 to 17.47) | 1.53<br>(1.41 to 1.67) | 1.11<br>(1.05 to 1.17) |
| Kuwait | 151<br>(130 to 176) | 24.52<br>(21.82 to 27.67) | 960<br>(831 to 1111) | 37.66<br>(33.53 to 41.9) | 5.34<br>(4.78 to 5.92) | 1.62<br>(1.57 to 1.67) |
| Kyrgyzstan | 222<br>(184 to 274) | 6.93<br>(5.85 to 8.33) | 505<br>(418 to 612) | 10.16<br>(8.6 to 12.01) | 1.28<br>(1.11 to 1.44) | 1.53<br>(1.29 to 1.76) |
| Lao People's Democratic Republic | 254<br>(219 to 294) | 12.28<br>(10.85 to 13.82) | 770<br>(674 to 882) | 17.51<br>(15.52 to 19.6) | 2.03<br>(1.9 to 2.18) | 1.26<br>(1.16 to 1.36) |
| Latvia | 278<br>(239 to 319) | 7.97<br>(6.83 to 9.17) | 450<br>(392 to 512) | 12.21<br>(10.57 to 14.09) | 0.62<br>(0.5 to 0.74) | 1.46<br>(1.25 to 1.68) |
| Lebanon | 485<br>(426 to 555) | 21.83<br>(19.34 to 24.59) | 2046<br>(1835 to 2260) | 39.2<br>(35.08 to 43.45) | 3.22<br>(2.95 to 3.5) | 2.17<br>(2 to 2.34) |
| Lesotho | 116<br>(101 to 131) | 12.02<br>(10.67 to 13.47) | 200<br>(173 to 230) | 15.97<br>(14.19 to 17.88) | 0.73<br>(0.65 to 0.83) | 0.86<br>(0.63 to 1.08) |
| Liberia | 113<br>(98 to 128) | 10.14<br>(9 to 11.37) | 273<br>(237 to 317) | 13.77<br>(12.15 to 15.54) | 1.43<br>(1.27 to 1.58) | 1.17<br>(1.06 to 1.29) |
| Libya | 400<br>(350 to 455) | 22.02<br>(19.53 to 24.6) | 1807<br>(1583 to 2032) | 36.73<br>(32.68 to 40.75) | 3.52<br>(3.33 to 3.72) | 1.91<br>(1.76 to 2.05) |

| | | | | | | |
|---|---|---|---|---|---|---|
| Lithuania | 383 (330 to 439) | 8.64 (7.45 to 9.9) | 586 (516 to 666) | 10.99 (9.62 to 12.52) | 0.53 (0.39 to 0.7) | 0.9 (0.71 to 1.08) |
| Luxembourg | 121 (108 to 135) | 21.18 (18.99 to 23.42) | 242 (218 to 266) | 23.84 (21.34 to 26.23) | 1 (0.87 to 1.16) | 0.43 (0.32 to 0.53) |
| Madagascar | 329 (287 to 378) | 6.78 (6 to 7.63) | 853 (737 to 1001) | 8.51 (7.59 to 9.57) | 1.59 (1.46 to 1.73) | 0.78 (0.7 to 0.85) |
| Malawi | 285 (251 to 325) | 7.93 (7.11 to 8.85) | 692 (605 to 786) | 10.1 (9.02 to 11.28) | 1.43 (1.3 to 1.57) | 0.95 (0.88 to 1.02) |
| Malaysia | 1230 (1072 to 1390) | 13.4 (11.92 to 14.99) | 5596 (4945 to 6284) | 20.59 (18.37 to 23.08) | 3.55 (3.28 to 3.85) | 1.58 (1.5 to 1.66) |
| Maldives | 14 (11 to 16) | 15.44 (13.49 to 17.44) | 72 (63 to 82) | 23.82 (21.01 to 27.1) | 4.3 (3.86 to 4.82) | 1.67 (1.57 to 1.78) |
| Mali | 398 (325 to 489) | 9.7 (8.11 to 11.66) | 1120 (915 to 1383) | 12.94 (10.81 to 15.86) | 1.82 (1.63 to 1.98) | 1.09 (0.98 to 1.19) |
| Malta | 95 (84 to 107) | 22.18 (19.85 to 24.77) | 236 (209 to 263) | 23.9 (21.3 to 26.6) | 1.48 (1.32 to 1.67) | 0.28 (0.2 to 0.35) |
| Marshall Islands | 2 (2 to 3) | 12.67 (11.17 to 14.46) | 6 (5 to 7) | 18.49 (16.44 to 20.88) | 1.92 (1.75 to 2.07) | 1.3 (1.27 to 1.32) |
| Mauritania | 116 (101 to 132) | 11.44 (10.15 to 12.86) | 324 (283 to 372) | 15.62 (13.77 to 17.6) | 1.8 (1.67 to 1.94) | 1.11 (1.06 to 1.17) |
| Mauritius | 145 (124 to 166) | 19.04 (16.55 to 21.5) | 525 (453 to 591) | 28.97 (25.01 to 32.57) | 2.63 (2.2 to 3.05) | 1.39 (1.29 to 1.48) |
| Mexico | 10288 (9283 to 11440) | 23.51 (21.19 to 26.18) | 41910 (38772 to 45160) | 34.89 (32.44 to 37.52) | 3.07 (2.82 to 3.34) | 1.29 (1.2 to 1.37) |
| Micronesia (Federated States of) | 7 (6 to 8) | 14.35 (12.4 to 16.35) | 16 (13 to 18) | 21.9 (19.21 to 24.99) | 1.29 (1.15 to 1.44) | 1.42 (1.37 to 1.46) |

| | | | | | | |
|---|---|---|---|---|---|---|
| Monaco | 14<br>(13 to 16) | 18.5<br>(16.4 to 20.77) | 21<br>(19 to 24) | 20.87<br>(18.59 to 23.25) | 0.47<br>(0.38 to 0.58) | 0.46<br>(0.43 to 0.5) |
| Mongolia | 119<br>(98 to 144) | 10.57<br>(9.07 to 12.33) | 340<br>(286 to 406) | 13.47<br>(11.62 to 15.47) | 1.87<br>(1.65 to 2.11) | 1.06<br>(0.92 to 1.19) |
| Montenegro | 93<br>(81 to 105) | 15.02<br>(13.18 to 16.98) | 213<br>(186 to 240) | 21.48<br>(18.94 to 24.24) | 1.3<br>(1.15 to 1.47) | 1.41<br>(1.35 to 1.46) |
| Morocco | 2245<br>(1938 to 2572) | 17.14<br>(14.99 to 19.37) | 10950<br>(9591 to 12408) | 35.5<br>(31.42 to 39.6) | 3.88<br>(3.65 to 4.12) | 2.67<br>(2.6 to 2.74) |
| Mozambique | 386<br>(338 to 439) | 7.07<br>(6.33 to 7.83) | 975<br>(847 to 1122) | 9.56<br>(8.47 to 10.7) | 1.53<br>(1.42 to 1.66) | 1.04<br>(1 to 1.09) |
| Myanmar | 2595<br>(2204 to 3045) | 11.22<br>(9.8 to 12.71) | 7712<br>(6695 to 8884) | 16.8<br>(14.84 to 19.05) | 1.97<br>(1.82 to 2.14) | 1.42<br>(1.31 to 1.53) |
| Namibia | 87<br>(76 to 100) | 12.17<br>(10.82 to 13.72) | 231<br>(203 to 263) | 16.73<br>(14.74 to 18.91) | 1.65<br>(1.52 to 1.78) | 1.17<br>(0.96 to 1.38) |
| Nauru | 1<br>(0 to 1) | 14.69<br>(12.82 to 16.83) | 1<br>(1 to 1) | 20.39<br>(17.94 to 23.22) | 0.55<br>(0.48 to 0.62) | 1.06<br>(0.99 to 1.12) |
| Nepal | 916<br>(786 to 1086) | 9.78<br>(8.57 to 11.26) | 3656<br>(3157 to 4228) | 16.12<br>(14.11 to 18.32) | 2.99<br>(2.78 to 3.21) | 1.53<br>(1.47 to 1.59) |
| Netherlands | 4148<br>(3766 to 4562) | 19.7<br>(17.97 to 21.51) | 8058<br>(7216 to 8938) | 22.26<br>(19.95 to 24.58) | 0.94<br>(0.8 to 1.09) | 0.51<br>(0.47 to 0.55) |
| New Zealand | 947<br>(842 to 1058) | 23.36<br>(20.93 to 25.9) | 2075<br>(1860 to 2304) | 25.75<br>(23.13 to 28.58) | 1.19<br>(1.04 to 1.38) | 0.4<br>(0.36 to 0.45) |
| Nicaragua | 331<br>(279 to 391) | 20.18<br>(17.41 to 23.31) | 1510<br>(1308 to 1742) | 31.52<br>(27.73 to 35.75) | 3.56<br>(3.31 to 3.84) | 1.55<br>(1.51 to 1.58) |
| Niger | 255<br>(221 to 298) | 9.59<br>(8.54 to 10.71) | 940<br>(814 to 1094) | 12.43<br>(11.02 to 14) | 2.68<br>(2.51 to 2.87) | 1.03<br>(0.94 to 1.12) |

| | | | | | | |
|---|---|---|---|---|---|---|
| Nigeria | 4482<br>(4049 to 4961) | 10.55<br>(9.59 to 11.61) | 11683<br>(10591 to 12911) | 14.1<br>(12.72 to 15.6) | 1.61<br>(1.55 to 1.67) | 1.22<br>(1.07 to 1.37) |
| Niue | 0<br>(0 to 0) | 14.56<br>(12.61 to 16.59) | 0<br>(0 to 1) | 22.15<br>(19.13 to 25.2) | 0.51<br>(0.45 to 0.59) | 1.42<br>(1.3 to 1.55) |
| North Macedonia | 218<br>(190 to 249) | 11.76<br>(10.33 to 13.35) | 693<br>(609 to 790) | 21.15<br>(18.73 to 23.94) | 2.19<br>(1.93 to 2.45) | 2.02<br>(1.93 to 2.11) |
| Northern Mariana Islands | 4<br>(3 to 5) | 19.32<br>(16.63 to 22.17) | 13<br>(11 to 15) | 24.42<br>(21.6 to 27.51) | 2.47<br>(1.93 to 3.08) | 0.69<br>(0.47 to 0.92) |
| Norway | 1152<br>(1011 to 1291) | 15.37<br>(13.67 to 17.09) | 1936<br>(1751 to 2138) | 19.15<br>(17.32 to 21.1) | 0.68<br>(0.57 to 0.8) | 0.8<br>(0.75 to 0.84) |
| Oman | 116<br>(101 to 134) | 18.74<br>(16.65 to 21.12) | 649<br>(560 to 746) | 37.27<br>(33.43 to 41.27) | 4.57<br>(4.23 to 4.92) | 2.34<br>(2.16 to 2.53) |
| Pakistan | 6405<br>(5675 to 7212) | 11.19<br>(9.96 to 12.47) | 16414<br>(14531 to 18524) | 14.88<br>(13.28 to 16.56) | 1.56<br>(1.47 to 1.65) | 1.1<br>(1.05 to 1.15) |
| Palau | 2<br>(1 to 2) | 16.61<br>(14.11 to 19.22) | 5<br>(4 to 6) | 23.57<br>(20.55 to 26.73) | 2.05<br>(1.81 to 2.3) | 1.08<br>(0.7 to 1.47) |
| Palestine | 207<br>(183 to 236) | 24.09<br>(21.56 to 26.93) | 901<br>(794 to 1020) | 38.32<br>(34.38 to 42.62) | 3.35<br>(3.15 to 3.56) | 1.57<br>(1.5 to 1.64) |
| Panama | 284<br>(248 to 326) | 18.86<br>(16.5 to 21.53) | 1141<br>(1024 to 1285) | 27.57<br>(24.74 to 31.04) | 3.01<br>(2.65 to 3.42) | 1.2<br>(1.13 to 1.26) |
| Papua New Guinea | 124<br>(104 to 150) | 6.71<br>(5.82 to 7.75) | 441<br>(371 to 530) | 9.36<br>(8.15 to 10.89) | 2.57<br>(2.37 to 2.79) | 1.02<br>(0.87 to 1.18) |
| Paraguay | 328<br>(283 to 376) | 14.74<br>(12.85 to 16.85) | 1321<br>(1160 to 1489) | 23.69<br>(20.93 to 26.58) | 3.03<br>(2.79 to 3.3) | 1.54<br>(1.5 to 1.59) |
| Peru | 1326<br>(1174 to 1499) | 11.47<br>(10.2 to 12.91) | 7012<br>(6279 to 7845) | 22.16<br>(19.73 to 24.8) | 4.29<br>(3.95 to 4.64) | 2.36<br>(2.24 to 2.47) |

| | | | | | | |
|---|---|---|---|---|---|---|
| Philippines | 4197 (3827 to 4585) | 14.28 (13.06 to 15.6) | 15924 (14530 to 17414) | 20.18 (18.4 to 21.99) | 2.79 (2.73 to 2.87) | 1.14 (1.12 to 1.15) |
| Poland | 5394 (4715 to 6193) | 12.42 (10.91 to 14.11) | 12086 (10645 to 13630) | 17.21 (15.29 to 19.3) | 1.24 (1.13 to 1.35) | 0.43 (0.2 to 0.67) |
| Portugal | 2608 (2269 to 2963) | 18.32 (16.17 to 20.59) | 5440 (4810 to 6095) | 21 (18.57 to 23.58) | 1.09 (0.95 to 1.24) | 0.46 (0.39 to 0.53) |
| Puerto Rico | 629 (551 to 713) | 17.3 (15.13 to 19.53) | 1706 (1509 to 1917) | 25.5 (22.43 to 28.8) | 1.71 (1.51 to 1.96) | 1.4 (1.21 to 1.6) |
| Qatar | 29 (25 to 35) | 26.84 (23.84 to 30.17) | 486 (407 to 582) | 44.21 (39.78 to 49.16) | 15.52 (14.46 to 16.63) | 1.8 (1.63 to 1.97) |
| Republic of Korea | 4913 (4278 to 5637) | 18.84 (16.63 to 21.38) | 19378 (17548 to 21335) | 21.65 (19.62 to 23.81) | 2.94 (2.55 to 3.38) | 0.5 (0.43 to 0.58) |
| Republic of Moldova | 256 (221 to 296) | 6.25 (5.44 to 7.17) | 499 (428 to 579) | 8.88 (7.62 to 10.34) | 0.95 (0.81 to 1.1) | 1.03 (0.82 to 1.25) |
| Romania | 2662 (2310 to 3066) | 9.71 (8.5 to 11.08) | 6113 (5406 to 6845) | 16.62 (14.71 to 18.67) | 1.3 (1.12 to 1.5) | 1.83 (1.72 to 1.95) |
| Russian Federation | 16501 (14745 to 18403) | 9.52 (8.6 to 10.57) | 31373 (28181 to 35221) | 13.96 (12.56 to 15.54) | 0.9 (0.85 to 0.96) | 1.48 (1.3 to 1.66) |
| Rwanda | 208 (180 to 239) | 7.6 (6.77 to 8.47) | 565 (490 to 647) | 10.08 (8.94 to 11.27) | 1.71 (1.56 to 1.86) | 1.21 (1.06 to 1.35) |
| Saint Kitts and Nevis | 7 (6 to 9) | 19.6 (16.33 to 23.58) | 19 (16 to 24) | 28.06 (23.58 to 33.4) | 1.73 (1.31 to 2.16) | 1.09 (0.91 to 1.28) |
| Saint Lucia | 13 (12 to 15) | 15.28 (13.6 to 17.26) | 50 (44 to 57) | 23.3 (20.68 to 26.46) | 2.72 (2.51 to 2.93) | 1.42 (1.28 to 1.56) |
| Saint Vincent and the Grenadines | 10 (9 to 12) | 14.25 (12.48 to 16.15) | 28 (25 to 32) | 21.02 (18.63 to 23.86) | 1.76 (1.62 to 1.93) | 1.34 (1.23 to 1.46) |

| | | | | | | |
|---|---|---|---|---|---|---|
| Samoa | 13<br>(11 to 15) | 14.54<br>(12.67 to 16.56) | 30<br>(26 to 34) | 20.45<br>(17.95 to 22.83) | 1.31<br>(1.19 to 1.42) | 1.06<br>(0.96 to 1.16) |
| San Marino | 6<br>(5 to 7) | 16.8<br>(14.84 to 18.93) | 13<br>(12 to 14) | 18.78<br>(16.68 to 20.96) | 1.24<br>(1.11 to 1.38) | 0.4<br>(0.36 to 0.44) |
| Sao Tome and Principe | 8<br>(7 to 9) | 12.55<br>(11.15 to 14.09) | 19<br>(16 to 22) | 17.93<br>(15.86 to 20.03) | 1.28<br>(1.16 to 1.43) | 1.33<br>(1.26 to 1.4) |
| Saudi Arabia | 1463<br>(1270 to 1678) | 24.97<br>(22.18 to 28.01) | 8910<br>(7606 to 10345) | 45.66<br>(41.39 to 50.22) | 5.09<br>(4.7 to 5.52) | 2.06<br>(1.94 to 2.18) |
| Senegal | 338<br>(295 to 383) | 10.64<br>(9.47 to 11.82) | 983<br>(867 to 1112) | 13.42<br>(11.96 to 14.92) | 1.91<br>(1.79 to 2.06) | 0.86<br>(0.8 to 0.92) |
| Serbia | 1303<br>(1140 to 1472) | 11.47<br>(10.09 to 12.9) | 3229<br>(2811 to 3668) | 19.85<br>(17.54 to 22.27) | 1.48<br>(1.26 to 1.73) | 2.01<br>(1.95 to 2.07) |
| Seychelles | 9<br>(8 to 11) | 16.64<br>(14.62 to 18.82) | 28<br>(24 to 31) | 25.15<br>(22.34 to 28) | 1.94<br>(1.73 to 2.16) | 1.47<br>(1.36 to 1.58) |
| Sierra Leone | 184<br>(162 to 209) | 9.67<br>(8.61 to 10.88) | 454<br>(397 to 523) | 12.84<br>(11.41 to 14.49) | 1.46<br>(1.34 to 1.6) | 1.05<br>(0.95 to 1.14) |
| Singapore | 452<br>(409 to 495) | 21.79<br>(19.93 to 23.79) | 1963<br>(1762 to 2163) | 25.54<br>(23.05 to 28.26) | 3.34<br>(3.06 to 3.67) | 0.61<br>(0.56 to 0.66) |
| Slovakia | 725<br>(637 to 838) | 12.19<br>(10.73 to 14.02) | 1691<br>(1462 to 1939) | 18.25<br>(15.99 to 20.88) | 1.33<br>(1.19 to 1.51) | 1.4<br>(1.36 to 1.43) |
| Slovenia | 274<br>(241 to 309) | 11.32<br>(9.99 to 12.78) | 776<br>(682 to 869) | 18.06<br>(15.94 to 20.15) | 1.83<br>(1.6 to 2.06) | 1.75<br>(1.71 to 1.79) |
| Solomon Islands | 17<br>(13 to 21) | 11.5<br>(9.96 to 13.38) | 43<br>(35 to 53) | 13.74<br>(11.94 to 15.83) | 1.59<br>(1.43 to 1.75) | 0.52<br>(0.44 to 0.6) |
| Somalia | 175<br>(152 to 204) | 7.63<br>(6.83 to 8.49) | 545<br>(470 to 630) | 8.91<br>(7.96 to 9.93) | 2.11<br>(1.96 to 2.26) | 0.64<br>(0.54 to 0.74) |

| | | | | | | |
|---|---|---|---|---|---|---|
| South Africa | 2811<br>(2528 to 3104) | 13.68<br>(12.35 to 15.08) | 8403<br>(7617 to 9246) | 19.12<br>(17.42 to 20.99) | 1.99<br>(1.89 to 2.09) | 1.12<br>(0.95 to 1.3) |
| South Sudan | 183<br>(160 to 206) | 8.06<br>(7.17 to 9.01) | 339<br>(295 to 383) | 9.56<br>(8.5 to 10.69) | 0.86<br>(0.76 to 0.95) | 0.64<br>(0.59 to 0.7) |
| Spain | 11818<br>(10574 to 13179) | 20.85<br>(18.82 to 22.97) | 22184<br>(19963 to 24521) | 21.47<br>(19.17 to 23.8) | 0.88<br>(0.77 to 1.03) | 0.15<br>(0.07 to 0.23) |
| Sri Lanka | 1585<br>(1386 to 1800) | 14.84<br>(13.17 to 16.52) | 5908<br>(5186 to 6623) | 22.51<br>(19.87 to 25.07) | 2.73<br>(2.48 to 2.98) | 1.51<br>(1.42 to 1.61) |
| Sudan | 1529<br>(1327 to 1751) | 17.16<br>(15.09 to 19.31) | 5610<br>(4902 to 6378) | 31.53<br>(27.95 to 35.42) | 2.67<br>(2.49 to 2.83) | 2.15<br>(2.11 to 2.18) |
| Suriname | 32<br>(26 to 39) | 12.02<br>(10.08 to 14.6) | 139<br>(117 to 168) | 22.82<br>(19.33 to 27.21) | 3.35<br>(2.98 to 3.73) | 2.34<br>(2.18 to 2.49) |
| Sweden | 2650<br>(2328 to 2985) | 15.93<br>(14.1 to 17.91) | 3721<br>(3271 to 4199) | 16.29<br>(14.4 to 18.29) | 0.4<br>(0.32 to 0.48) | 0.13<br>(0.11 to 0.15) |
| Switzerland | 2258<br>(2008 to 2544) | 20.36<br>(18.2 to 22.78) | 4257<br>(3814 to 4716) | 23.17<br>(20.76 to 25.74) | 0.89<br>(0.78 to 1) | 0.42<br>(0.37 to 0.48) |
| Syrian Arab Republic | 1122<br>(979 to 1281) | 22.09<br>(19.66 to 24.81) | 4482<br>(4010 to 5006) | 36.27<br>(32.86 to 40.12) | 2.99<br>(2.71 to 3.26) | 1.91<br>(1.73 to 2.09) |
| Taiwan (Province of China) | 3425<br>(3059 to 3808) | 22.16<br>(20.04 to 24.34) | 10198<br>(9363 to 11105) | 25.75<br>(23.65 to 28.07) | 1.98<br>(1.79 to 2.19) | 0.51<br>(0.43 to 0.59) |
| Tajikistan | 174<br>(144 to 211) | 5.91<br>(4.98 to 7.04) | 543<br>(440 to 666) | 10.15<br>(8.56 to 11.86) | 2.12<br>(1.9 to 2.34) | 2.09<br>(1.8 to 2.38) |
| Thailand | 5582<br>(4848 to 6429) | 15.72<br>(13.89 to 17.76) | 24734<br>(22180 to 27414) | 24.06<br>(21.56 to 26.69) | 3.43<br>(3.08 to 3.84) | 1.46<br>(1.36 to 1.57) |
| Timor-Leste | 31<br>(26 to 36) | 10.83<br>(9.62 to 12.18) | 129<br>(111 to 147) | 15.81<br>(13.96 to 17.89) | 3.2<br>(2.84 to 3.61) | 1.44<br>(1.34 to 1.54) |

| | | | | | | |
|---|---|---|---|---|---|---|
| Togo | 132<br>(114 to 152) | 10.84<br>(9.58 to 12.18) | 491<br>(425 to 561) | 13.84<br>(12.23 to 15.41) | 2.71<br>(2.54 to 2.89) | 0.94<br>(0.84 to 1.05) |
| Tokelau | 0<br>(0 to 0) | 12.03<br>(10.58 to 13.62) | 0<br>(0 to 0) | 19.35<br>(17 to 21.94) | 0.59<br>(0.5 to 0.68) | 1.62<br>(1.6 to 1.65) |
| Tonga | 8<br>(7 to 9) | 13.99<br>(12.29 to 15.88) | 15<br>(14 to 17) | 19.56<br>(17.35 to 22.06) | 0.98<br>(0.89 to 1.09) | 1.07<br>(1.02 to 1.12) |
| Trinidad and Tobago | 117<br>(102 to 134) | 13.8<br>(12.12 to 15.6) | 411<br>(361 to 467) | 22.05<br>(19.39 to 24.97) | 2.51<br>(2.24 to 2.82) | 1.66<br>(1.57 to 1.75) |
| Tunisia | 1047<br>(911 to 1188) | 21.21<br>(18.78 to 23.8) | 4718<br>(4185 to 5247) | 37.26<br>(33.3 to 41.16) | 3.51<br>(3.28 to 3.73) | 2.05<br>(1.92 to 2.19) |
| Turkey | 6401<br>(5740 to 7140) | 18.96<br>(17.14 to 20.97) | 31447<br>(28075 to 35198) | 35.87<br>(32.17 to 39.93) | 3.91<br>(3.54 to 4.31) | 2.45<br>(2.27 to 2.64) |
| Turkmenistan | 167<br>(137 to 206) | 8.02<br>(6.82 to 9.39) | 510<br>(430 to 610) | 12.13<br>(10.45 to 14.1) | 2.06<br>(1.83 to 2.36) | 1.59<br>(1.4 to 1.79) |
| Tuvalu | 1<br>(1 to 1) | 11.43<br>(10.03 to 13.05) | 2<br>(2 to 2) | 18.45<br>(16.28 to 20.99) | 1.42<br>(1.3 to 1.56) | 1.54<br>(1.48 to 1.6) |
| Uganda | 434<br>(380 to 492) | 7.19<br>(6.43 to 7.98) | 1177<br>(1029 to 1337) | 9.09<br>(8.08 to 10.16) | 1.71<br>(1.58 to 1.85) | 0.91<br>(0.81 to 1.01) |
| Ukraine | 5082<br>(4372 to 5881) | 7.44<br>(6.41 to 8.58) | 6806<br>(5969 to 7674) | 9.29<br>(8.13 to 10.59) | 0.34<br>(0.24 to 0.45) | 0.82<br>(0.59 to 1.05) |
| United Arab Emirates | 128<br>(105 to 152) | 28.49<br>(25.36 to 31.68) | 1963<br>(1590 to 2369) | 43.84<br>(39.65 to 48.39) | 14.37<br>(13.44 to 15.43) | 1.47<br>(1.36 to 1.58) |
| United Kingdom | 17804<br>(15901 to 19869) | 18.35<br>(16.51 to 20.29) | 26694<br>(24011 to 29547) | 19.96<br>(18.01 to 22.07) | 0.5<br>(0.47 to 0.53) | 0.1<br>(-0.01 to 0.22) |
| United Republic of Tanzania | 863<br>(755 to 983) | 8.37<br>(7.46 to 9.34) | 2441<br>(2150 to 2770) | 10.62<br>(9.43 to 11.79) | 1.83<br>(1.68 to 1.99) | 0.84<br>(0.78 to 0.89) |

| | | | | | | |
|---|---|---|---|---|---|---|
| United States of America | 85254<br>(77045 to 94075) | 25.85<br>(23.53 to 28.36) | 157609<br>(144239 to 172082) | 27.85<br>(25.53 to 30.31) | 0.85<br>(0.79 to 0.92) | 0.11<br>(0.04 to 0.19) |
| United States Virgin Islands | 13<br>(11 to 15) | 14.81<br>(13.08 to 16.8) | 42<br>(37 to 48) | 22.83<br>(20.18 to 25.92) | 2.3<br>(2.04 to 2.58) | 1.51<br>(1.34 to 1.68) |
| Uruguay | 686<br>(617 to 771) | 17.2<br>(15.54 to 19.19) | 1198<br>(1082 to 1337) | 20.88<br>(18.86 to 23.22) | 0.75<br>(0.62 to 0.88) | 0.66<br>(0.64 to 0.68) |
| Uzbekistan | 1229<br>(1034 to 1496) | 10.14<br>(8.74 to 11.81) | 3594<br>(2998 to 4364) | 15.17<br>(13.13 to 17.64) | 1.92<br>(1.75 to 2.11) | 1.46<br>(1.29 to 1.62) |
| Vanuatu | 7<br>(6 to 8) | 10.96<br>(9.62 to 12.53) | 27<br>(23 to 32) | 16.05<br>(14.07 to 18.26) | 2.81<br>(2.64 to 3.01) | 1.18<br>(1.09 to 1.26) |
| Venezuela (Bolivarian Republic of) | 1871<br>(1627 to 2165) | 18.83<br>(16.54 to 21.48) | 8443<br>(7483 to 9470) | 28.56<br>(25.59 to 31.94) | 3.51<br>(3.12 to 3.92) | 1.45<br>(1.35 to 1.55) |
| Viet Nam | 3982<br>(3495 to 4553) | 10.09<br>(8.97 to 11.35) | 14272<br>(12662 to 16117) | 15.94<br>(14.16 to 17.85) | 2.58<br>(2.36 to 2.83) | 1.74<br>(1.67 to 1.82) |
| Yemen | 767<br>(664 to 881) | 16.72<br>(14.8 to 18.87) | 3649<br>(3190 to 4227) | 28.89<br>(25.46 to 32.55) | 3.76<br>(3.46 to 4.19) | 2.08<br>(2.04 to 2.13) |
| Zambia | 237<br>(205 to 274) | 8.76<br>(7.84 to 9.8) | 719<br>(620 to 834) | 11.39<br>(10.08 to 12.79) | 2.04<br>(1.89 to 2.19) | 0.98<br>(0.88 to 1.09) |
| Zimbabwe | 523<br>(451 to 606) | 13.1<br>(11.58 to 14.78) | 1046<br>(900 to 1213) | 15.41<br>(13.68 to 17.32) | 1<br>(0.93 to 1.08) | 0.38<br>(0.09 to 0.67) |

Table S6. The prevalence count and ASPR of CKD due to hypertension in 1990 and 2019 for both sexes in 204 countries, and its temporal trends from 1990 to 2019.

| location | prevalence count_1990 (95% UI) | ASPR_1990 (95% UI) | prevalence count_2019 (95% UI) | ASPR_2019 (95% UI) | prevalence count_change (*100%) (95% CI) | EAPC (95% CI) |
|---|---|---|---|---|---|---|
| Afghanistan | 23343 (20777 to 26448) | 359.16 (320.92 to 402.99) | 68351 (59805 to 78896) | 580.15 (519.55 to 650.41) | 1.93 (1.75 to 2.13) | 1.8 (1.7 to 1.9) |
| Albania | 5027 (4433 to 5766) | 239.74 (214.41 to 269.38) | 14483 (13040 to 16017) | 352.15 (316.11 to 390.03) | 1.88 (1.6 to 2.16) | 1.39 (1.35 to 1.44) |
| Algeria | 47361 (41941 to 54367) | 416.52 (372.9 to 466.92) | 219693 (196131 to 245300) | 711.47 (639.5 to 789.49) | 3.64 (3.41 to 3.87) | 1.89 (1.81 to 1.97) |
| American Samoa | 99 (86 to 115) | 409.68 (365.64 to 460.01) | 264 (236 to 296) | 572.37 (515.73 to 634.01) | 1.67 (1.48 to 1.86) | 1.13 (1.04 to 1.23) |
| Andorra | 149 (132 to 167) | 315.93 (282.32 to 351.79) | 477 (429 to 527) | 328.97 (293.96 to 365.44) | 2.21 (2.01 to 2.42) | 0.14 (0.04 to 0.23) |
| Angola | 6220 (5193 to 7766) | 148.16 (131.04 to 169.62) | 24372 (20627 to 29546) | 211.16 (187.09 to 238.1) | 2.92 (2.72 to 3.1) | 1.34 (1.23 to 1.45) |
| Antigua and Barbuda | 205 (178 to 237) | 373.89 (325.73 to 430.77) | 609 (532 to 707) | 620.9 (543.14 to 720.54) | 1.97 (1.8 to 2.19) | 1.87 (1.74 to 2) |
| Argentina | 98092 (87778 to 109457) | 321.25 (288.92 to 357.34) | 218246 (197483 to 240775) | 398.65 (360.43 to 440.61) | 1.22 (1.08 to 1.39) | 0.8 (0.7 to 0.89) |
| Armenia | 6154 (5306 to 7273) | 227.63 (198.95 to 262.74) | 14205 (12658 to 15983) | 357.58 (318.1 to 403.66) | 1.31 (1.11 to 1.52) | 1.72 (1.6 to 1.84) |
| Australia | 68759 (63907 to 73661) | 357.91 (332.97 to 383.54) | 179279 (164355 to 196325) | 405.31 (370.67 to 445.09) | 1.61 (1.45 to 1.77) | 0.4 (0.36 to 0.44) |
| Austria | 37166 (33150 to 41640) | 305.36 (273.7 to 341.16) | 74012 (66514 to 81432) | 380.43 (342.75 to 417.26) | 0.99 (0.87 to 1.13) | 0.74 (0.69 to 0.8) |
| Azerbaijan | 13052 (11260 to 15352) | 249.51 (219.8 to 285.97) | 35823 (31102 to 41035) | 392.4 (346.61 to 440.62) | 1.74 (1.57 to 1.94) | 1.68 (1.51 to 1.86) |

| | | | | | | |
|---|---|---|---|---|---|---|
| Bahamas | 556<br>(487 to 637) | 346.22<br>(309.34 to 384.61) | 1860<br>(1659 to 2051) | 492.62<br>(442.49 to 541.89) | 2.35<br>(2.15 to 2.55) | 1.18<br>(1.03 to 1.33) |
| Bahrain | 785<br>(684 to 915) | 472.51<br>(426.68 to 527.1) | 6694<br>(5870 to 7539) | 806.47<br>(726.89 to 889.9) | 7.53<br>(6.83 to 8.18) | 2.01<br>(1.88 to 2.14) |
| Bangladesh | 88774<br>(74964 to 110652) | 182.25<br>(160.26 to 209.41) | 342611<br>(301596 to 393685) | 267.45<br>(237.56 to 304.43) | 2.86<br>(2.48 to 3.19) | 1.38<br>(1.25 to 1.5) |
| Barbados | 989<br>(880 to 1096) | 336.21<br>(298.72 to 374.27) | 2344<br>(2111 to 2597) | 496.21<br>(445.96 to 549.68) | 1.37<br>(1.24 to 1.51) | 1.33<br>(1.21 to 1.44) |
| Belarus | 29609<br>(26043 to 34118) | 239.47<br>(209 to 278.16) | 45469<br>(40688 to 51072) | 303.85<br>(268.98 to 350.42) | 0.54<br>(0.44 to 0.64) | 0.91<br>(0.66 to 1.15) |
| Belgium | 50991<br>(45550 to 56453) | 326.62<br>(292.7 to 360.26) | 89009<br>(80904 to 97496) | 353.63<br>(322.08 to 387.03) | 0.75<br>(0.6 to 0.9) | 0.31<br>(0.24 to 0.39) |
| Belize | 328<br>(290 to 375) | 334.52<br>(297.81 to 374.8) | 1456<br>(1297 to 1646) | 516<br>(465.63 to 572.41) | 3.44<br>(3.14 to 3.74) | 1.49<br>(1.42 to 1.56) |
| Benin | 4547<br>(4001 to 5271) | 215.41<br>(193.91 to 241.53) | 14807<br>(13011 to 17293) | 289.16<br>(259.7 to 324.38) | 2.26<br>(2.14 to 2.38) | 1.08<br>(1.01 to 1.15) |
| Bermuda | 200<br>(180 to 222) | 324.91<br>(292.33 to 360.82) | 632<br>(571 to 700) | 497.84<br>(448.9 to 550.33) | 2.17<br>(1.97 to 2.36) | 1.5<br>(1.34 to 1.66) |
| Bhutan | 615<br>(525 to 743) | 235.07<br>(209.58 to 263.3) | 2023<br>(1791 to 2289) | 356.99<br>(320.08 to 396.7) | 2.29<br>(1.99 to 2.52) | 1.62<br>(1.55 to 1.69) |
| Bolivia (Plurinational State of) | 8320<br>(7371 to 9445) | 262.29<br>(236.96 to 290.54) | 33341<br>(29926 to 36908) | 396.61<br>(358.85 to 437.41) | 3.01<br>(2.8 to 3.24) | 1.46<br>(1.41 to 1.51) |
| Bosnia and Herzegovina | 9168<br>(8144 to 10442) | 239.27<br>(214.64 to 267.47) | 21148<br>(19091 to 23299) | 372.92<br>(334.93 to 410.58) | 1.31<br>(1.11 to 1.52) | 1.81<br>(1.7 to 1.91) |
| Botswana | 1445<br>(1249 to 1686) | 254.15<br>(225.11 to 285.7) | 5154<br>(4453 to 5952) | 385.66<br>(344.25 to 433.7) | 2.57<br>(2.41 to 2.71) | 1.37<br>(1.22 to 1.52) |

| | | | | | | |
|---|---|---|---|---|---|---|
| Brazil | 280171 (256653 to 303871) | 321.36 (294.61 to 348.39) | 942266 (868875 to 1021494) | 407.65 (376.53 to 441.88) | 2.36 (2.24 to 2.51) | 0.76 (0.7 to 0.82) |
| Brunei Darussalam | 416 (360 to 487) | 461.49 (412.31 to 512.5) | 1262 (1098 to 1450) | 495.68 (440.55 to 556.96) | 2.04 (1.84 to 2.2) | 0.26 (0.12 to 0.41) |
| Bulgaria | 30243 (26746 to 33811) | 265.79 (235.75 to 295.98) | 56348 (50632 to 62195) | 403.19 (361.22 to 445.52) | 0.86 (0.73 to 1.02) | 1.49 (1.44 to 1.53) |
| Burkina Faso | 8363 (7279 to 9620) | 195.36 (174.18 to 218.45) | 24828 (21413 to 28648) | 266.7 (238.71 to 296.72) | 1.97 (1.85 to 2.1) | 1.19 (1.1 to 1.29) |
| Burundi | 3965 (3403 to 4744) | 158.53 (141.02 to 179.16) | 9250 (7933 to 11072) | 195.12 (173.79 to 219.97) | 1.33 (1.25 to 1.42) | 0.87 (0.78 to 0.95) |
| Cabo Verde | 481 (429 to 545) | 200.64 (180.08 to 225.7) | 1361 (1215 to 1547) | 310.78 (278.96 to 347.43) | 1.83 (1.71 to 1.95) | 1.73 (1.61 to 1.84) |
| Cambodia | 11544 (9872 to 13943) | 240.86 (213.79 to 276.25) | 40534 (35627 to 46989) | 343.28 (307.87 to 387.2) | 2.51 (2.25 to 2.74) | 1.27 (1.19 to 1.35) |
| Cameroon | 12398 (10754 to 14309) | 274.19 (244.55 to 307.32) | 48726 (42253 to 56451) | 393.4 (351.45 to 440.56) | 2.93 (2.79 to 3.09) | 1.39 (1.29 to 1.48) |
| Canada | 113579 (101218 to 127655) | 353.76 (317.22 to 395.73) | 258008 (233650 to 286272) | 364.81 (331.52 to 405.9) | 1.27 (1.04 to 1.49) | 0.35 (0.26 to 0.45) |
| Central African Republic | 1800 (1503 to 2167) | 148.56 (130.89 to 168.52) | 4185 (3500 to 5015) | 184.77 (162.99 to 208.19) | 1.33 (1.25 to 1.41) | 0.87 (0.78 to 0.95) |
| Chad | 5752 (5016 to 6694) | 199.17 (176.46 to 226.1) | 15001 (12864 to 17770) | 254.68 (226.03 to 288.83) | 1.61 (1.5 to 1.72) | 0.91 (0.79 to 1.03) |
| Chile | 30326 (27033 to 34124) | 324.05 (290.39 to 361.28) | 110366 (99979 to 121182) | 464.53 (419.71 to 510.79) | 2.64 (2.36 to 2.94) | 1.36 (1.26 to 1.46) |
| China | 2245024 (2071687 to 2438393) | 279.14 (257.23 to 302.84) | 5577334 (5161604 to 6034387) | 300.76 (278.72 to 325.76) | 1.48 (1.41 to 1.55) | 0.67 (0.55 to 0.8) |

| | | | | | | |
|---|---|---|---|---|---|---|
| Colombia | 70077<br>(62182 to 79252) | 396.89<br>(357.64 to 440.4) | 283284<br>(256603 to 313896) | 531.74<br>(480.37 to 589.56) | 3.04<br>(2.72 to 3.38) | 0.94<br>(0.88 to 1.01) |
| Comoros | 400<br>(350 to 468) | 171.74<br>(153.48 to 192.87) | 1091<br>(964 to 1251) | 221.84<br>(198.93 to 247.78) | 1.73<br>(1.62 to 1.86) | 1<br>(0.92 to 1.07) |
| Congo | 1877<br>(1594 to 2258) | 169.83<br>(150.28 to 193.51) | 6220<br>(5384 to 7402) | 233.39<br>(207.13 to 262.25) | 2.31<br>(2.18 to 2.45) | 1.29<br>(1.17 to 1.4) |
| Cook Islands | 46<br>(41 to 53) | 366.79<br>(327.22 to 416.57) | 127<br>(114 to 141) | 536.14<br>(479.89 to 600.17) | 1.74<br>(1.53 to 1.91) | 1.25<br>(1.19 to 1.32) |
| Costa Rica | 12938<br>(12020 to 13991) | 741.03<br>(697.19 to 790.13) | 40102<br>(37569 to 42862) | 783.04<br>(735.18 to 836.44) | 2.1<br>(2 to 2.2) | 0.2<br>(0.19 to 0.21) |
| Côte d'Ivoire | 10447<br>(8847 to 12723) | 239.95<br>(214.86 to 267.78) | 34226<br>(29408 to 40770) | 308<br>(275.49 to 344.5) | 2.28<br>(2.13 to 2.43) | 0.97<br>(0.88 to 1.06) |
| Croatia | 17415<br>(15540 to 19277) | 288.79<br>(258.02 to 321.41) | 36440<br>(32972 to 40326) | 413.43<br>(374.89 to 456.91) | 1.09<br>(0.93 to 1.28) | 1.46<br>(1.39 to 1.53) |
| Cuba | 28578<br>(25503 to 32051) | 276.54<br>(247.57 to 308.92) | 81346<br>(73494 to 90098) | 437.53<br>(394.56 to 485.45) | 1.85<br>(1.66 to 2.04) | 1.6<br>(1.54 to 1.67) |
| Cyprus | 2683<br>(2348 to 3035) | 370.51<br>(330.91 to 413.99) | 7544<br>(6665 to 8497) | 398.76<br>(355.27 to 447.21) | 1.81<br>(1.67 to 1.96) | 0.3<br>(0.2 to 0.41) |
| Czechia | 34951<br>(31474 to 38711) | 262.28<br>(235.17 to 291) | 75075<br>(67356 to 83246) | 362.62<br>(326 to 401.44) | 1.15<br>(1.04 to 1.26) | 1.11<br>(1.03 to 1.18) |
| Democratic People's Republic of Korea | 45920<br>(40343 to 52910) | 303.08<br>(272.13 to 339.36) | 108178<br>(96846 to 120654) | 353.91<br>(317.84 to 394.67) | 1.36<br>(1.22 to 1.5) | 0.58<br>(0.46 to 0.71) |
| Democratic Republic of the Congo | 25406<br>(21535 to 31077) | 157.8<br>(139.59 to 181.04) | 75253<br>(64333 to 90830) | 204.01<br>(182.31 to 231.82) | 1.96<br>(1.86 to 2.08) | 0.9<br>(0.8 to 0.99) |
| Denmark | 24467<br>(21808 to 27376) | 291.48<br>(260.15 to 324.23) | 41684<br>(37488 to 46036) | 347.64<br>(313.78 to 383.67) | 0.7<br>(0.57 to 0.84) | 0.71<br>(0.66 to 0.76) |

| | | | | | | |
|---|---|---|---|---|---|---|
| Djibouti | 278<br>(228 to 353) | 169.73<br>(150.39 to 193.28) | 1417<br>(1222 to 1685) | 232.79<br>(208.48 to 262.95) | 4.1<br>(3.73 to 4.46) | 1.26<br>(1.13 to 1.38) |
| Dominica | 285<br>(254 to 315) | 398.16<br>(355.08 to 441.27) | 476<br>(430 to 524) | 539.51<br>(486.14 to 597.26) | 0.67<br>(0.59 to 0.74) | 1.04<br>(0.89 to 1.19) |
| Dominican Republic | 9627<br>(8340 to 11266) | 245.78<br>(218.26 to 277.16) | 38016<br>(33907 to 42277) | 409.46<br>(366.06 to 454.29) | 2.95<br>(2.67 to 3.23) | 1.6<br>(1.49 to 1.71) |
| Ecuador | 14966<br>(13166 to 17211) | 275.07<br>(245.46 to 309.34) | 77897<br>(70137 to 86090) | 528.14<br>(477.28 to 581.99) | 4.2<br>(3.81 to 4.64) | 2.25<br>(2.13 to 2.37) |
| Egypt | 111011<br>(97688 to 126522) | 416.44<br>(372.19 to 466.81) | 414949<br>(367942 to 469758) | 746.85<br>(670.93 to 827.79) | 2.74<br>(2.57 to 2.9) | 2<br>(1.9 to 2.11) |
| El Salvador | 12304<br>(10808 to 14223) | 405.12<br>(358.91 to 461.89) | 40675<br>(36335 to 45275) | 671.91<br>(599.8 to 749.27) | 2.31<br>(2.07 to 2.54) | 1.92<br>(1.78 to 2.05) |
| Equatorial Guinea | 288<br>(244 to 345) | 145.46<br>(127.74 to 166.29) | 1351<br>(1133 to 1616) | 254.12<br>(225.49 to 284.18) | 3.68<br>(3.43 to 3.96) | 2.25<br>(2.11 to 2.4) |
| Eritrea | 1640<br>(1345 to 2048) | 147.56<br>(129.54 to 168.44) | 5852<br>(4921 to 7067) | 203.73<br>(180.77 to 228.57) | 2.57<br>(2.39 to 2.75) | 1.11<br>(1.05 to 1.17) |
| Estonia | 5417<br>(4787 to 6155) | 277.11<br>(242.72 to 319.03) | 10015<br>(8934 to 11173) | 399.5<br>(352.04 to 452.51) | 0.85<br>(0.73 to 0.97) | 1.48<br>(1.33 to 1.63) |
| Eswatini | 868<br>(753 to 1016) | 283.75<br>(252.03 to 321.33) | 2229<br>(1954 to 2561) | 377.83<br>(337.93 to 421.54) | 1.57<br>(1.46 to 1.68) | 0.9<br>(0.68 to 1.12) |
| Ethiopia | 32924<br>(29591 to 36666) | 157.65<br>(143.79 to 172.91) | 90943<br>(81828 to 101009) | 202.26<br>(183.73 to 222.62) | 1.76<br>(1.69 to 1.84) | 0.91<br>(0.82 to 0.99) |
| Fiji | 1492<br>(1272 to 1738) | 382.93<br>(338.74 to 430.84) | 3537<br>(3118 to 3974) | 491.43<br>(440.56 to 547.38) | 1.37<br>(1.21 to 1.54) | 0.78<br>(0.75 to 0.81) |
| Finland | 16499<br>(14750 to 18432) | 234.06<br>(209.43 to 260.69) | 36312<br>(33012 to 40029) | 277.26<br>(251.46 to 304.46) | 1.2<br>(1.07 to 1.36) | 0.41<br>(0.33 to 0.49) |

| | | | | | | |
|---|---|---|---|---|---|---|
| France | 244044 (218001 to 269250) | 280.43 (252.49 to 308.93) | 503102 (452500 to 553221) | 328.55 (295.44 to 362.61) | 1.06 (0.93 to 1.2) | 0.49 (0.44 to 0.54) |
| Gabon | 1051 (911 to 1232) | 189.65 (167.75 to 214.01) | 2907 (2542 to 3366) | 281.49 (250.89 to 313.59) | 1.77 (1.66 to 1.88) | 1.45 (1.38 to 1.52) |
| Gambia | 809 (691 to 985) | 214.11 (190.6 to 240.86) | 2923 (2549 to 3418) | 286.44 (255.35 to 320) | 2.61 (2.43 to 2.78) | 1.04 (0.98 to 1.09) |
| Georgia | 14457 (12675 to 16655) | 246.68 (216.12 to 284.53) | 20186 (18245 to 22379) | 353.69 (314.27 to 399.77) | 0.4 (0.29 to 0.5) | 1.18 (1.1 to 1.27) |
| Germany | 428147 (386866 to 475444) | 330.48 (299.68 to 366.72) | 783944 (718963 to 847879) | 374.26 (346.12 to 402.87) | 0.83 (0.71 to 0.96) | 0.32 (0.27 to 0.37) |
| Ghana | 12814 (10843 to 15507) | 197.54 (174.86 to 223.46) | 46143 (39816 to 54093) | 282.1 (249.65 to 314.91) | 2.6 (2.42 to 2.77) | 1.24 (1.17 to 1.3) |
| Greece | 58613 (51536 to 65333) | 394.12 (348.03 to 440.72) | 105300 (93855 to 116806) | 392.43 (349.62 to 438.1) | 0.8 (0.69 to 0.9) | 0.07 (-0.06 to 0.19) |
| Greenland | 89 (78 to 103) | 304.96 (273.94 to 339.83) | 204 (185 to 227) | 350.01 (316.29 to 389.85) | 1.31 (1.1 to 1.5) | 0.46 (0.43 to 0.49) |
| Grenada | 266 (238 to 296) | 358.85 (320.14 to 399.41) | 617 (549 to 689) | 574.29 (510.33 to 639.61) | 1.32 (1.19 to 1.46) | 1.49 (1.36 to 1.62) |
| Guam | 290 (250 to 348) | 363.64 (323.65 to 410.84) | 905 (810 to 1010) | 491.87 (437.43 to 549.63) | 2.12 (1.84 to 2.37) | 0.91 (0.77 to 1.04) |
| Guatemala | 14448 (12703 to 16792) | 404.82 (361.17 to 462.08) | 73201 (64720 to 82997) | 645.15 (574.17 to 722.51) | 4.07 (3.79 to 4.34) | 1.6 (1.49 to 1.7) |
| Guinea | 6919 (6072 to 7897) | 208.46 (185.95 to 232.72) | 15802 (13805 to 18253) | 275.68 (247.11 to 306.02) | 1.28 (1.2 to 1.38) | 1.02 (0.94 to 1.09) |
| Guinea-Bissau | 937 (808 to 1103) | 219.54 (194.74 to 246.69) | 2206 (1878 to 2632) | 281.8 (250.28 to 315.95) | 1.35 (1.27 to 1.44) | 0.95 (0.87 to 1.04) |

| | | | | | | |
|---|---|---|---|---|---|---|
| Guyana | 1270<br>(1108 to 1472) | 314.85<br>(283.61 to 352.03) | 3031<br>(2677 to 3414) | 495.64<br>(443.52 to 550.57) | 1.39<br>(1.22 to 1.56) | 1.39<br>(1.3 to 1.47) |
| Haiti | 8819<br>(7613 to 10173) | 268.59<br>(238.11 to 303.8) | 27055<br>(23591 to 31189) | 382.35<br>(342.2 to 429.66) | 2.07<br>(1.96 to 2.18) | 1.27<br>(1.26 to 1.29) |
| Honduras | 8397<br>(7374 to 9650) | 398.52<br>(354.55 to 452.21) | 33868<br>(30173 to 38244) | 560.32<br>(504.55 to 622.98) | 3.03<br>(2.83 to 3.25) | 1.15<br>(1.13 to 1.18) |
| Hungary | 32414<br>(28912 to 36334) | 232.32<br>(207 to 262.63) | 70795<br>(63379 to 79361) | 369.7<br>(329.05 to 415.46) | 1.18<br>(1.06 to 1.32) | 1.59<br>(1.52 to 1.66) |
| Iceland | 796<br>(723 to 886) | 268.91<br>(243.86 to 298.77) | 1678<br>(1517 to 1848) | 286.15<br>(258.82 to 315.94) | 1.11<br>(0.98 to 1.25) | 0.11<br>(-0.01 to 0.22) |
| India | 1358638<br>(1252191 to 1473753) | 304.48<br>(281.53 to 330.11) | 3717836<br>(3446782 to 4010903) | 338.69<br>(314.03 to 364.86) | 1.74<br>(1.67 to 1.81) | 0.29<br>(0.19 to 0.39) |
| Indonesia | 283249<br>(262099 to 309184) | 269.3<br>(248.69 to 289.75) | 776074<br>(717420 to 840412) | 361.96<br>(334.72 to 390.39) | 1.74<br>(1.66 to 1.81) | 0.98<br>(0.92 to 1.03) |
| Iran (Islamic Republic of) | 119518<br>(108697 to 131051) | 507.8<br>(462.78 to 556.19) | 466110<br>(428623 to 504067) | 672.36<br>(618.4 to 729.13) | 2.9<br>(2.77 to 3.06) | 0.95<br>(0.87 to 1.03) |
| Iraq | 36103<br>(32136 to 40485) | 472.56<br>(423.1 to 526.13) | 163783<br>(146060 to 183222) | 770.13<br>(696.13 to 853.34) | 3.54<br>(3.36 to 3.7) | 1.9<br>(1.84 to 1.97) |
| Ireland | 15660<br>(13842 to 17321) | 387.15<br>(345.21 to 425.76) | 28665<br>(27371 to 30112) | 380.74<br>(362.54 to 403.31) | 0.83<br>(0.68 to 1.05) | -0.1<br>(-0.14 to -0.07) |
| Israel | 18277<br>(16246 to 20462) | 383.55<br>(342.98 to 427.27) | 51037<br>(45983 to 56567) | 425.17<br>(383.94 to 471.05) | 1.79<br>(1.61 to 1.97) | 0.38<br>(0.27 to 0.49) |
| Italy | 270156<br>(243742 to 299616) | 308.17<br>(280.24 to 340.9) | 506483<br>(460940 to 560030) | 314.97<br>(288.53 to 345.08) | 0.87<br>(0.8 to 0.95) | 0.08<br>(0.01 to 0.15) |
| Jamaica | 6425<br>(5738 to 7221) | 351.51<br>(313.22 to 393.56) | 14648<br>(13175 to 16216) | 481.5<br>(433.35 to 534.3) | 1.28<br>(1.17 to 1.39) | 1.16<br>(0.98 to 1.34) |

| | | | | | | |
|---|---|---|---|---|---|---|
| Japan | 701987 (650816 to 759408) | 432.58 (401.87 to 466.92) | 1799081 (1652762 to 1964976) | 458.58 (425.82 to 498.25) | 1.56 (1.48 to 1.65) | 0.16 (0.13 to 0.19) |
| Jordan | 5762 (5099 to 6572) | 460.29 (413.92 to 509.48) | 46466 (41798 to 51575) | 784.99 (716.79 to 860.81) | 7.06 (6.61 to 7.52) | 2.04 (1.97 to 2.12) |
| Kazakhstan | 32122 (27659 to 38018) | 247.4 (217.07 to 286.62) | 61575 (53504 to 70570) | 363.3 (320.49 to 413.71) | 0.92 (0.81 to 1.02) | 1.63 (1.41 to 1.84) |
| Kenya | 14785 (13564 to 16060) | 165.81 (152.33 to 180.85) | 48174 (44123 to 52364) | 207.11 (189.53 to 227.15) | 2.26 (2.2 to 2.32) | 0.72 (0.64 to 0.8) |
| Kiribati | 127 (107 to 152) | 311.91 (273.94 to 356.92) | 300 (255 to 355) | 413.02 (365.39 to 467.38) | 1.36 (1.26 to 1.46) | 0.93 (0.9 to 0.96) |
| Kuwait | 3007 (2607 to 3461) | 495.02 (446.58 to 552.1) | 17620 (15658 to 19740) | 726.98 (653.11 to 808.8) | 4.86 (4.39 to 5.35) | 1.47 (1.42 to 1.51) |
| Kyrgyzstan | 8147 (7018 to 9536) | 253.39 (221.95 to 289.47) | 15826 (13613 to 18355) | 325.26 (287.18 to 367.01) | 0.94 (0.85 to 1.03) | 1.03 (0.87 to 1.18) |
| Lao People's Democratic Republic | 6416 (5573 to 7627) | 300.5 (265.67 to 343.15) | 18859 (16432 to 22084) | 421.4 (375.39 to 472.64) | 1.94 (1.83 to 2.05) | 1.21 (1.13 to 1.3) |
| Latvia | 8414 (7376 to 9561) | 247.17 (214.69 to 285.02) | 12698 (11391 to 14106) | 342.63 (302.46 to 386.28) | 0.51 (0.41 to 0.61) | 1.17 (1.02 to 1.33) |
| Lebanon | 8849 (7859 to 9969) | 427.82 (382.96 to 478.15) | 39911 (36014 to 44026) | 764.61 (690.67 to 843.74) | 3.51 (3.28 to 3.74) | 2.19 (2.04 to 2.33) |
| Lesotho | 2315 (2026 to 2659) | 239.12 (212.65 to 267.87) | 4047 (3526 to 4637) | 322.92 (289.04 to 359) | 0.75 (0.68 to 0.82) | 0.91 (0.7 to 1.13) |
| Liberia | 2283 (2001 to 2660) | 208.46 (185.94 to 235.59) | 6170 (5324 to 7391) | 285.94 (254.19 to 320.6) | 1.7 (1.57 to 1.84) | 1.24 (1.11 to 1.36) |
| Libya | 7835 (6941 to 8838) | 432.61 (388.05 to 481.32) | 33808 (30259 to 37438) | 709.42 (636.74 to 787.66) | 3.32 (3.13 to 3.48) | 1.87 (1.73 to 2.01) |

| | | | | | | |
|---|---|---|---|---|---|---|
| Lithuania | 11477 (10094 to 13146) | 262.15 (229.76 to 302.57) | 16848 (15103 to 18842) | 315.88 (280.91 to 361.13) | 0.47 (0.36 to 0.57) | 0.74 (0.6 to 0.88) |
| Luxembourg | 1778 (1595 to 1966) | 327.12 (294.88 to 360.96) | 3864 (3510 to 4236) | 365.37 (331.94 to 400.74) | 1.17 (1.05 to 1.33) | 0.43 (0.32 to 0.54) |
| Madagascar | 8439 (7190 to 10251) | 155.06 (136.32 to 177.45) | 21974 (18546 to 26877) | 191.41 (169.83 to 216.63) | 1.6 (1.51 to 1.7) | 0.72 (0.66 to 0.79) |
| Malawi | 8512 (6844 to 11262) | 208.69 (175.42 to 257.39) | 20161 (16635 to 26028) | 255.94 (219.34 to 306.35) | 1.37 (1.26 to 1.49) | 0.8 (0.74 to 0.86) |
| Malaysia | 31043 (27024 to 36101) | 325.07 (291.77 to 366.24) | 124521 (111800 to 138819) | 481.33 (436.6 to 531.23) | 3.01 (2.74 to 3.25) | 1.46 (1.38 to 1.53) |
| Maldives | 312 (268 to 369) | 352.47 (312.07 to 397.9) | 1682 (1479 to 1959) | 530.95 (477.92 to 593.47) | 4.39 (4.09 to 4.73) | 1.59 (1.49 to 1.7) |
| Mali | 8029 (6953 to 9312) | 196.52 (175.36 to 220.16) | 23354 (20237 to 26931) | 265.44 (236.47 to 296.04) | 1.91 (1.81 to 2.01) | 1.09 (1 to 1.18) |
| Malta | 1402 (1251 to 1550) | 345.81 (309.95 to 382.35) | 3638 (3264 to 4003) | 376.72 (339.21 to 414.76) | 1.6 (1.44 to 1.77) | 0.32 (0.25 to 0.39) |
| Marshall Islands | 61 (53 to 73) | 329.55 (292.6 to 368.62) | 159 (139 to 183) | 454.99 (405.88 to 504.92) | 1.6 (1.47 to 1.72) | 1.12 (1.08 to 1.15) |
| Mauritania | 2449 (2129 to 2849) | 237.42 (210.23 to 268.16) | 6838 (5987 to 7886) | 323.91 (287.63 to 362.53) | 1.79 (1.69 to 1.9) | 1.1 (1.06 to 1.15) |
| Mauritius | 3443 (3029 to 3921) | 462.3 (409.63 to 515) | 11717 (10434 to 13104) | 699.19 (622.14 to 778.42) | 2.4 (2.08 to 2.75) | 1.37 (1.28 to 1.46) |
| Mexico | 232922 (210628 to 257091) | 534.67 (483.95 to 595.31) | 940862 (864126 to 1018762) | 811.09 (745.97 to 878.73) | 3.04 (2.81 to 3.27) | 1.36 (1.29 to 1.44) |
| Micronesia (Federated States of) | 181 (155 to 211) | 361.7 (320.95 to 409.7) | 376 (326 to 429) | 540.75 (480.94 to 603.37) | 1.08 (0.97 to 1.2) | 1.38 (1.32 to 1.43) |

| | | | | | | |
|---|---|---|---|---|---|---|
| Monaco | 224<br>(199 to 251) | 292.21<br>(262.35 to 326.43) | 341<br>(305 to 379) | 327.47<br>(294.18 to 363.23) | 0.52<br>(0.44 to 0.61) | 0.43<br>(0.4 to 0.46) |
| Mongolia | 3372<br>(2873 to 4069) | 293.62<br>(257.15 to 338.51) | 9212<br>(7958 to 10806) | 378<br>(334.71 to 427.68) | 1.73<br>(1.58 to 1.89) | 1.08<br>(0.97 to 1.19) |
| Montenegro | 1992<br>(1783 to 2227) | 330.9<br>(297.01 to 368.57) | 4181<br>(3762 to 4644) | 443.43<br>(397.76 to 493.65) | 1.1<br>(0.97 to 1.23) | 1.19<br>(1.13 to 1.25) |
| Morocco | 42624<br>(37746 to 48200) | 331.41<br>(295.86 to 370.63) | 192376<br>(170987 to 213158) | 684.37<br>(613.17 to 756.22) | 3.51<br>(3.26 to 3.77) | 2.69<br>(2.64 to 2.74) |
| Mozambique | 9247<br>(7825 to 11413) | 151.76<br>(133.7 to 175.75) | 24380<br>(20649 to 29858) | 208.11<br>(185.53 to 236.52) | 1.64<br>(1.53 to 1.76) | 1.11<br>(1.07 to 1.14) |
| Myanmar | 71816<br>(61801 to 84289) | 298.65<br>(264.21 to 339.45) | 191862<br>(168920 to 218340) | 422.98<br>(377.76 to 474.73) | 1.67<br>(1.52 to 1.81) | 1.24<br>(1.15 to 1.33) |
| Namibia | 1726<br>(1514 to 1992) | 241.78<br>(216.39 to 271.57) | 4643<br>(4099 to 5318) | 332.85<br>(298.11 to 373.87) | 1.69<br>(1.59 to 1.8) | 1.18<br>(0.98 to 1.37) |
| Nauru | 17<br>(14 to 21) | 389.09<br>(345.57 to 441.32) | 25<br>(21 to 30) | 534.68<br>(477.16 to 602.08) | 0.48<br>(0.42 to 0.54) | 1.01<br>(0.92 to 1.11) |
| Nepal | 21285<br>(18474 to 24760) | 228.64<br>(203.88 to 256.7) | 81465<br>(72617 to 91908) | 374.26<br>(338 to 416.06) | 2.83<br>(2.58 to 3.05) | 1.39<br>(1.29 to 1.49) |
| Netherlands | 60953<br>(55772 to 67184) | 300.92<br>(275.92 to 332.63) | 121955<br>(109835 to 134844) | 345.34<br>(311.98 to 382.36) | 1<br>(0.86 to 1.15) | 0.57<br>(0.52 to 0.62) |
| New Zealand | 14488<br>(13085 to 16057) | 373<br>(338.33 to 412.28) | 33265<br>(30182 to 36827) | 412.15<br>(374.09 to 456.71) | 1.3<br>(1.14 to 1.46) | 0.4<br>(0.36 to 0.44) |
| Nicaragua | 7919<br>(6932 to 9142) | 486.77<br>(432.01 to 547.83) | 34718<br>(30767 to 38969) | 768.16<br>(691.02 to 851) | 3.38<br>(3.13 to 3.66) | 1.57<br>(1.52 to 1.61) |
| Niger | 6762<br>(5580 to 8195) | 219.7<br>(190.06 to 255.19) | 23344<br>(19430 to 28065) | 281.93<br>(243.66 to 327.72) | 2.45<br>(2.3 to 2.6) | 0.96<br>(0.85 to 1.06) |

| | | | | | | |
|---|---|---|---|---|---|---|
| Nigeria | 98369<br>(90507 to 106526) | 224.98<br>(206.98 to 245.16) | 260869<br>(239044 to 282889) | 296.43<br>(271.37 to 323.47) | 1.65<br>(1.6 to 1.7) | 1.12<br>(1.02 to 1.23) |
| Niue | 8<br>(7 to 9) | 381.38<br>(339.54 to 426.32) | 12<br>(10 to 13) | 558.29<br>(497.14 to 621.7) | 0.38<br>(0.33 to 0.45) | 1.33<br>(1.22 to 1.43) |
| North Macedonia | 4844<br>(4320 to 5433) | 273.54<br>(245.19 to 304.33) | 13074<br>(11784 to 14593) | 438.95<br>(397.15 to 489.5) | 1.7<br>(1.51 to 1.89) | 1.68<br>(1.58 to 1.78) |
| Northern Mariana Islands | 100<br>(81 to 122) | 468.83<br>(410.87 to 530.93) | 279<br>(246 to 315) | 591.11<br>(526.81 to 659.51) | 1.81<br>(1.43 to 2.23) | 0.7<br>(0.48 to 0.92) |
| Norway | 17159<br>(15502 to 18994) | 240.84<br>(219.72 to 264.63) | 29855<br>(27211 to 32899) | 294.76<br>(269.04 to 322.39) | 0.74<br>(0.65 to 0.83) | 0.77<br>(0.73 to 0.81) |
| Oman | 2269<br>(1969 to 2638) | 368.26<br>(331.86 to 411.72) | 11411<br>(9959 to 13097) | 707.81<br>(643.87 to 778.98) | 4.03<br>(3.78 to 4.3) | 2.23<br>(2.08 to 2.39) |
| Pakistan | 155514<br>(138247 to 173787) | 266.58<br>(239.01 to 294.91) | 387983<br>(344110 to 437772) | 343.14<br>(307.58 to 378.08) | 1.49<br>(1.42 to 1.56) | 0.99<br>(0.94 to 1.04) |
| Palau | 44<br>(38 to 51) | 430.9<br>(374.68 to 488.7) | 119<br>(104 to 133) | 602.36<br>(535.1 to 669.9) | 1.7<br>(1.51 to 1.88) | 1.07<br>(0.77 to 1.36) |
| Palestine | 4065<br>(3623 to 4574) | 477.27<br>(430.2 to 531.18) | 16084<br>(14375 to 18019) | 741.6<br>(671 to 821.62) | 2.96<br>(2.81 to 3.12) | 1.5<br>(1.45 to 1.55) |
| Panama | 6682<br>(5864 to 7590) | 439.25<br>(389.81 to 490.97) | 25634<br>(23018 to 28359) | 615.83<br>(552.58 to 679.71) | 2.84<br>(2.49 to 3.2) | 1.05<br>(0.98 to 1.11) |
| Papua New Guinea | 4474<br>(3728 to 5504) | 217.87<br>(189.98 to 250.02) | 14872<br>(12621 to 17868) | 280.7<br>(247.87 to 318.23) | 2.32<br>(2.2 to 2.46) | 0.79<br>(0.7 to 0.87) |
| Paraguay | 7051<br>(6205 to 7971) | 315.33<br>(279.67 to 351.17) | 26907<br>(24089 to 29940) | 488.95<br>(440.35 to 540.57) | 2.82<br>(2.59 to 3.07) | 1.44<br>(1.4 to 1.48) |
| Peru | 31922<br>(27622 to 37912) | 262.68<br>(230.82 to 302.29) | 146703<br>(131357 to 164674) | 454.39<br>(406.8 to 509.37) | 3.6<br>(3.23 to 4) | 1.98<br>(1.89 to 2.07) |

| | | | | | | |
|---|---|---|---|---|---|---|
| Philippines | 105969<br>(98182 to 114430) | 339.97<br>(315.72 to 367.02) | 372292<br>(343574 to 402332) | 479.91<br>(442.59 to 518.49) | 2.51<br>(2.44 to 2.58) | 1.13<br>(1.11 to 1.15) |
| Poland | 119690<br>(108118 to 133272) | 284.07<br>(257.45 to 315.15) | 236892<br>(214348 to 263447) | 343.48<br>(311.45 to 379.7) | 0.98<br>(0.91 to 1.05) | 0.32<br>(0.2 to 0.45) |
| Portugal | 38012<br>(33667 to 43080) | 288.37<br>(256.59 to 324.46) | 86688<br>(76816 to 97072) | 328.74<br>(290.76 to 367.72) | 1.28<br>(1.14 to 1.43) | 0.5<br>(0.38 to 0.63) |
| Puerto Rico | 14405<br>(12863 to 15949) | 398.32<br>(356.49 to 441.26) | 40596<br>(36337 to 45100) | 578.62<br>(518.26 to 644.59) | 1.82<br>(1.6 to 2.05) | 1.38<br>(1.2 to 1.57) |
| Qatar | 573<br>(477 to 695) | 511.14<br>(456.35 to 573.28) | 7659<br>(6467 to 9003) | 852.19<br>(766.61 to 946.09) | 12.38<br>(11.34 to 13.35) | 1.9<br>(1.75 to 2.05) |
| Republic of Korea | 85710<br>(75383 to 96621) | 327.98<br>(293.56 to 365.9) | 305077<br>(279399 to 330438) | 355.83<br>(324.78 to 386.36) | 2.56<br>(2.23 to 2.89) | 0.36<br>(0.29 to 0.43) |
| Republic of Moldova | 8504<br>(7293 to 10144) | 202.64<br>(174.9 to 239.2) | 15180<br>(13375 to 17349) | 276.4<br>(241.62 to 321.36) | 0.79<br>(0.66 to 0.92) | 0.95<br>(0.82 to 1.08) |
| Romania | 61064<br>(54054 to 68613) | 233.79<br>(207.68 to 262.61) | 129593<br>(116971 to 142462) | 352.99<br>(318.11 to 389.99) | 1.12<br>(0.98 to 1.28) | 1.42<br>(1.34 to 1.49) |
| Russian Federation | 530336<br>(489299 to 572498) | 310.53<br>(287.39 to 334.78) | 906488<br>(829545 to 987662) | 407.9<br>(374.69 to 442.84) | 0.71<br>(0.67 to 0.75) | 1.12<br>(1.02 to 1.23) |
| Rwanda | 4948<br>(4193 to 6053) | 161.34<br>(142.27 to 185.52) | 12987<br>(11208 to 15481) | 213.28<br>(189.45 to 240.18) | 1.62<br>(1.52 to 1.75) | 1.21<br>(1.07 to 1.35) |
| Saint Kitts and Nevis | 157<br>(139 to 176) | 425.13<br>(378.88 to 474.32) | 368<br>(326 to 414) | 592.96<br>(528.71 to 661.72) | 1.34<br>(1.19 to 1.49) | 1.09<br>(0.92 to 1.25) |
| Saint Lucia | 313<br>(278 to 350) | 351.18<br>(314.65 to 388.88) | 1101<br>(991 to 1226) | 523.54<br>(472.04 to 582.83) | 2.52<br>(2.32 to 2.73) | 1.35<br>(1.21 to 1.48) |
| Saint Vincent and the Grenadines | 247<br>(218 to 277) | 337.54<br>(300.91 to 374.6) | 634<br>(565 to 701) | 486.63<br>(434.98 to 536.99) | 1.57<br>(1.43 to 1.71) | 1.27<br>(1.16 to 1.38) |

| | | | | | | |
|---|---|---|---|---|---|---|
| Samoa | 334<br>(292 to 387) | 372.04<br>(331.19 to 417.69) | 736<br>(650 to 833) | 505.06<br>(448.66 to 562.04) | 1.2<br>(1.11 to 1.3) | 0.96<br>(0.87 to 1.06) |
| San Marino | 86<br>(76 to 95) | 264.34<br>(237.02 to 293.31) | 208<br>(186 to 231) | 292.67<br>(261.91 to 326.77) | 1.43<br>(1.3 to 1.57) | 0.37<br>(0.34 to 0.39) |
| Sao Tome and Principe | 168<br>(147 to 192) | 261.69<br>(232.57 to 292.33) | 409<br>(357 to 468) | 377.73<br>(336.88 to 418.63) | 1.44<br>(1.33 to 1.53) | 1.38<br>(1.31 to 1.45) |
| Saudi Arabia | 28066<br>(24688 to 31854) | 489.75<br>(439.36 to 539.91) | 151419<br>(133307 to 170121) | 926.25<br>(841.29 to 1008.65) | 4.4<br>(4.08 to 4.77) | 2.21<br>(2.1 to 2.32) |
| Senegal | 7176<br>(6300 to 8294) | 213.71<br>(191.28 to 238.97) | 20451<br>(18047 to 23247) | 269.23<br>(241.4 to 299.39) | 1.85<br>(1.75 to 1.96) | 0.85<br>(0.79 to 0.91) |
| Serbia | 27615<br>(24955 to 31245) | 260.08<br>(234.99 to 292.88) | 61549<br>(55403 to 68453) | 397.17<br>(357.74 to 441.31) | 1.23<br>(1.07 to 1.4) | 1.61<br>(1.55 to 1.67) |
| Seychelles | 222<br>(197 to 250) | 384.48<br>(344.06 to 432.8) | 594<br>(535 to 659) | 566.36<br>(513.2 to 628.94) | 1.68<br>(1.53 to 1.85) | 1.38<br>(1.29 to 1.47) |
| Sierra Leone | 4003<br>(3478 to 4603) | 202.43<br>(180.1 to 226.86) | 10298<br>(8946 to 11949) | 270.67<br>(243.79 to 302.46) | 1.57<br>(1.47 to 1.68) | 1.07<br>(0.98 to 1.15) |
| Singapore | 8805<br>(8018 to 9865) | 421.51<br>(389.91 to 455.82) | 35350<br>(32357 to 39226) | 473.37<br>(433.8 to 522.48) | 3.01<br>(2.74 to 3.32) | 0.48<br>(0.43 to 0.53) |
| Slovakia | 16237<br>(14473 to 18160) | 279.61<br>(249.24 to 313.04) | 33888<br>(30243 to 37869) | 379.31<br>(338.32 to 422.44) | 1.09<br>(0.98 to 1.21) | 1.07<br>(1.04 to 1.1) |
| Slovenia | 5874<br>(5256 to 6556) | 247.09<br>(221.66 to 276.44) | 15395<br>(13847 to 16823) | 350.98<br>(315.1 to 387.03) | 1.62<br>(1.43 to 1.81) | 1.34<br>(1.31 to 1.38) |
| Solomon Islands | 507<br>(428 to 609) | 331.1<br>(292.8 to 378.81) | 1360<br>(1150 to 1610) | 394.97<br>(349.44 to 446.79) | 1.68<br>(1.59 to 1.78) | 0.52<br>(0.46 to 0.58) |
| Somalia | 4406<br>(3703 to 5378) | 160.26<br>(142.25 to 182.76) | 13912<br>(11591 to 17098) | 190.36<br>(169.87 to 214.97) | 2.16<br>(2.05 to 2.28) | 0.73<br>(0.64 to 0.82) |

| | | | | | | |
|---|---|---|---|---|---|---|
| South Africa | 60763 (55410 to 66190) | 284.9 (259.17 to 311.85) | 166759 (153669 to 181243) | 384.57 (354.56 to 418.42) | 1.74 (1.67 to 1.83) | 1.01 (0.83 to 1.18) |
| South Sudan | 4186 (3585 to 5011) | 164.33 (146.05 to 185.25) | 7884 (6769 to 9356) | 198.68 (176.38 to 222.77) | 0.88 (0.82 to 0.96) | 0.73 (0.68 to 0.79) |
| Spain | 170575 (154027 to 188425) | 317.14 (286.87 to 348.85) | 339906 (302825 to 377576) | 309.82 (278.47 to 343.25) | 0.99 (0.86 to 1.14) | 0.05 (-0.03 to 0.13) |
| Sri Lanka | 37975 (33387 to 43745) | 349.76 (313.5 to 391.94) | 124966 (112288 to 138607) | 516.75 (464.42 to 571.35) | 2.29 (2.02 to 2.53) | 1.43 (1.35 to 1.52) |
| Sudan | 28954 (25659 to 33542) | 328.18 (292.86 to 370.04) | 102848 (90810 to 116471) | 587.91 (522.23 to 653.78) | 2.55 (2.4 to 2.72) | 2.04 (2 to 2.09) |
| Suriname | 881 (775 to 992) | 331.23 (293.69 to 370.92) | 2935 (2628 to 3247) | 502.37 (450.61 to 554.5) | 2.33 (2.13 to 2.56) | 1.44 (1.4 to 1.47) |
| Sweden | 42086 (37473 to 46967) | 268.5 (240.73 to 299.77) | 61891 (55418 to 68684) | 277.04 (248.17 to 309.04) | 0.47 (0.4 to 0.54) | 0.16 (0.13 to 0.19) |
| Switzerland | 35645 (32117 to 39309) | 326.01 (295.31 to 358.54) | 67393 (60815 to 73869) | 352.61 (317.68 to 386.64) | 0.89 (0.79 to 1) | 0.29 (0.24 to 0.34) |
| Syrian Arab Republic | 22454 (19762 to 25470) | 438.15 (392.52 to 487.58) | 75315 (68058 to 83847) | 697.04 (635.48 to 769.66) | 2.35 (2.13 to 2.59) | 1.83 (1.65 to 2) |
| Taiwan (Province of China) | 64646 (58541 to 72882) | 443.72 (405.5 to 490.59) | 228806 (213163 to 245694) | 590.47 (546.63 to 640.27) | 2.54 (2.29 to 2.79) | 1.04 (0.96 to 1.12) |
| Tajikistan | 6240 (5244 to 7561) | 202.76 (175.49 to 237.57) | 15966 (13265 to 19434) | 293.18 (255.55 to 339.3) | 1.56 (1.42 to 1.7) | 1.42 (1.21 to 1.64) |
| Thailand | 144139 (125011 to 166797) | 395.01 (352.38 to 438.91) | 536813 (489807 to 589731) | 546.32 (497.21 to 603.23) | 2.72 (2.39 to 3.04) | 1.22 (1.13 to 1.32) |
| Timor-Leste | 881 (736 to 1084) | 274.28 (242.76 to 311.31) | 3084 (2709 to 3519) | 384.04 (341.26 to 429.58) | 2.5 (2.15 to 2.83) | 1.29 (1.21 to 1.37) |

| | | | | | | |
|---|---|---|---|---|---|---|
| Togo | 2999<br>(2565 to 3626) | 223.48<br>(198.39 to 251.68) | 10559<br>(9168 to 12352) | 288.72<br>(258.78 to 322.85) | 2.52<br>(2.36 to 2.68) | 0.99<br>(0.89 to 1.09) |
| Tokelau | 4<br>(4 to 5) | 317.29<br>(279.45 to 359.01) | 6<br>(5 to 7) | 482.52<br>(428.5 to 539.2) | 0.48<br>(0.42 to 0.54) | 1.45<br>(1.43 to 1.46) |
| Tonga | 202<br>(178 to 234) | 360.56<br>(322.2 to 409.61) | 387<br>(346 to 437) | 484.42<br>(434.14 to 541.65) | 0.92<br>(0.83 to 1) | 0.96<br>(0.92 to 1.01) |
| Trinidad and Tobago | 2798<br>(2490 to 3171) | 327.48<br>(293.48 to 366.96) | 9066<br>(8202 to 10106) | 505.44<br>(457.12 to 563.08) | 2.24<br>(1.99 to 2.53) | 1.56<br>(1.47 to 1.65) |
| Tunisia | 18768<br>(16733 to 21045) | 405.85<br>(366.81 to 450.39) | 83360<br>(75227 to 91878) | 705.36<br>(638.31 to 776.84) | 3.44<br>(3.22 to 3.67) | 2.04<br>(1.93 to 2.15) |
| Turkey | 121599<br>(109802 to 136500) | 357.96<br>(327.13 to 394.11) | 566157<br>(512888 to 625916) | 669.5<br>(607.49 to 737.27) | 3.66<br>(3.31 to 4.03) | 2.44<br>(2.22 to 2.65) |
| Turkmenistan | 5556<br>(4739 to 6715) | 264.07<br>(231.9 to 303.32) | 15373<br>(13435 to 17967) | 378.63<br>(336.8 to 429.69) | 1.77<br>(1.59 to 1.96) | 1.4<br>(1.26 to 1.54) |
| Tuvalu | 21<br>(18 to 24) | 309.53<br>(273.27 to 351.24) | 46<br>(40 to 52) | 462.49<br>(412.86 to 517.59) | 1.2<br>(1.1 to 1.32) | 1.3<br>(1.25 to 1.35) |
| Uganda | 10384<br>(8807 to 12789) | 151.79<br>(133.55 to 173.92) | 29047<br>(24656 to 35724) | 191.83<br>(170.59 to 216.57) | 1.8<br>(1.69 to 1.9) | 0.98<br>(0.88 to 1.09) |
| Ukraine | 152097<br>(133295 to 173992) | 229.88<br>(200.79 to 267.24) | 198630<br>(176261 to 221869) | 280.16<br>(246.18 to 319) | 0.31<br>(0.24 to 0.37) | 0.78<br>(0.63 to 0.93) |
| United Arab Emirates | 2504<br>(2093 to 3025) | 539.95<br>(485.44 to 600.92) | 30300<br>(25532 to 35325) | 856.34<br>(772.87 to 945.78) | 11.1<br>(10.14 to 11.96) | 1.55<br>(1.42 to 1.68) |
| United Kingdom | 264860<br>(240016 to 292597) | 288.41<br>(263.03 to 316.3) | 417667<br>(377893 to 462243) | 314.59<br>(286.24 to 346.35) | 0.58<br>(0.55 to 0.6) | 0.13<br>(0.04 to 0.22) |
| United Republic of Tanzania | 19491<br>(16811 to 22846) | 176.34<br>(156.63 to 198.3) | 57595<br>(50095 to 66860) | 226.6<br>(200.71 to 253) | 1.96<br>(1.82 to 2.12) | 0.91<br>(0.83 to 0.98) |

| | | | | | | |
|---|---|---|---|---|---|---|
| United States of America | 1533786<br>(1397951 to 1696595) | 466<br>(426.19 to 513.32) | 2857254<br>(2625010 to 3117828) | 504.07<br>(463.64 to 548.07) | 0.86<br>(0.8 to 0.93) | 0.12<br>(0.01 to 0.22) |
| United States Virgin Islands | 290<br>(259 to 326) | 348.33<br>(313.51 to 388.12) | 898<br>(804 to 1003) | 516.84<br>(463.64 to 572.82) | 2.1<br>(1.89 to 2.32) | 1.39<br>(1.22 to 1.55) |
| Uruguay | 11183<br>(10104 to 12395) | 292.69<br>(264.52 to 325.29) | 19978<br>(18264 to 21980) | 344.86<br>(314.96 to 379.87) | 0.79<br>(0.68 to 0.91) | 0.57<br>(0.54 to 0.6) |
| Uzbekistan | 37356<br>(32375 to 44018) | 305.51<br>(270.42 to 346.09) | 92322<br>(79571 to 109022) | 424.7<br>(378.47 to 479.95) | 1.47<br>(1.38 to 1.57) | 1.23<br>(1.1 to 1.37) |
| Vanuatu | 210<br>(178 to 250) | 295.03<br>(260.21 to 332.96) | 711<br>(620 to 820) | 403.36<br>(359.22 to 452.64) | 2.38<br>(2.19 to 2.57) | 0.98<br>(0.91 to 1.05) |
| Venezuela (Bolivarian Republic of) | 43895<br>(38761 to 50061) | 442.32<br>(396.89 to 496.3) | 180913<br>(162648 to 198519) | 635.7<br>(575.18 to 695.3) | 3.12<br>(2.76 to 3.49) | 1.28<br>(1.18 to 1.38) |
| Viet Nam | 107560<br>(94397 to 126146) | 262.79<br>(234.81 to 297.55) | 336229<br>(300978 to 380209) | 377.39<br>(339.45 to 419.66) | 2.13<br>(1.94 to 2.31) | 1.42<br>(1.34 to 1.5) |
| Yemen | 14545<br>(12597 to 16735) | 323.29<br>(287.64 to 363.9) | 65565<br>(57429 to 74583) | 536.38<br>(477.69 to 599.26) | 3.51<br>(3.25 to 3.9) | 1.94<br>(1.89 to 1.99) |
| Zambia | 5495<br>(4701 to 6469) | 178.52<br>(159.37 to 200.19) | 17428<br>(14850 to 20692) | 240.25<br>(214.09 to 268.84) | 2.17<br>(2.06 to 2.3) | 1.14<br>(1.04 to 1.24) |
| Zimbabwe | 10816<br>(9082 to 12834) | 260.99<br>(225.58 to 303.53) | 23287<br>(19755 to 27359) | 329.07<br>(286.26 to 383.18) | 1.15<br>(1.08 to 1.23) | 0.6<br>(0.32 to 0.88) |

Table S7. The DALYs count and ASDR of CKD due to hypertension in 1990 and 2019 for both sexes in 204 countries, and its temporal trends from 1990 to 2019.

| location | DALYs count_1990 (95% UI) | ASDR_1990 (95% UI) | DALYs count_2019 (95% UI) | ASDR_2019 (95% UI) | DALYs count_change (*100%) (95% CI) | EAPC (95% CI) |
|---|---|---|---|---|---|---|
| Afghanistan | 25249 (17671 to 36165) | 365.54 (256.08 to 525.75) | 40034 (28452 to 57037) | 322.53 (231.92 to 469.78) | 0.59 (0.13 to 1.11) | -0.38 (-0.43 to -0.32) |
| Albania | 1354 (1081 to 1645) | 64.18 (51.33 to 77.91) | 2040 (1533 to 2684) | 50.34 (38.15 to 65.7) | 0.51 (0.17 to 0.9) | -1.31 (-1.61 to -1.02) |
| Algeria | 24706 (17548 to 35241) | 231.73 (169.33 to 325.59) | 59156 (44407 to 77436) | 195.54 (148.13 to 254.4) | 1.39 (0.82 to 2.16) | -0.48 (-0.62 to -0.33) |
| American Samoa | 74 (58 to 93) | 313.77 (246.72 to 382.42) | 218 (172 to 272) | 458.43 (367.79 to 566.92) | 1.93 (1.38 to 2.55) | 1.43 (1.31 to 1.56) |
| Andorra | 25 (18 to 33) | 55.76 (41.67 to 73.18) | 75 (58 to 96) | 49.56 (38.3 to 63.96) | 2.03 (1.22 to 3) | -0.39 (-0.53 to -0.24) |
| Angola | 7990 (5637 to 10633) | 199.51 (143.93 to 260.25) | 19498 (13621 to 26272) | 174.41 (121.95 to 228.49) | 1.44 (0.81 to 2.33) | -0.54 (-0.58 to -0.5) |
| Antigua and Barbuda | 82 (67 to 99) | 152.39 (124.52 to 183.88) | 215 (172 to 265) | 219.35 (177.2 to 267.66) | 1.62 (1.2 to 2.14) | 1.53 (1.35 to 1.71) |
| Argentina | 46909 (38927 to 56291) | 149.47 (124.65 to 178.39) | 93201 (77382 to 112054) | 170.82 (142.2 to 205.21) | 0.99 (0.73 to 1.24) | 0.39 (0.15 to 0.64) |
| Armenia | 426 (319 to 561) | 15.63 (12.1 to 19.96) | 1506 (1176 to 1888) | 38.39 (29.86 to 47.85) | 2.54 (1.86 to 3.42) | 3.31 (3.19 to 3.42) |
| Australia | 5971 (5151 to 6945) | 32.34 (27.64 to 37.91) | 19091 (14677 to 24385) | 41.68 (32.59 to 52.89) | 2.2 (1.62 to 2.86) | 0.96 (0.88 to 1.04) |
| Austria | 4511 (3668 to 5439) | 37.87 (31.04 to 45.18) | 14505 (11529 to 17609) | 70 (56.22 to 84.01) | 2.22 (1.77 to 2.76) | 3.79 (3.24 to 4.34) |

| | | | | | | |
|---|---|---|---|---|---|---|
| Azerbaijan | 1731<br>(1346 to 2192) | 31.14<br>(24.53 to 38.98) | 4791<br>(3670 to 6114) | 54.13<br>(41.95 to 69.24) | 1.77<br>(1.3 to 2.3) | 2.43<br>(2.09 to 2.76) |
| Bahamas | 231<br>(190 to 284) | 142.85<br>(117.75 to 173.11) | 697<br>(542 to 892) | 179.6<br>(140.33 to 228.33) | 2.01<br>(1.44 to 2.72) | 1.14<br>(1 to 1.29) |
| Bahrain | 340<br>(269 to 429) | 227.7<br>(180.37 to 287.44) | 1554<br>(1218 to 1950) | 208.84<br>(164.18 to 258.79) | 3.57<br>(2.61 to 4.73) | -0.18<br>(-0.63 to 0.29) |
| Bangladesh | 34489<br>(25413 to 45410) | 71.21<br>(52.65 to 93.08) | 79577<br>(60561 to 102816) | 62.01<br>(47.73 to 79.85) | 1.31<br>(0.77 to 2.02) | -0.02<br>(-0.29 to 0.25) |
| Barbados | 315<br>(260 to 374) | 109.43<br>(90.31 to 131.34) | 722<br>(573 to 882) | 152.86<br>(122.43 to 186.12) | 1.29<br>(0.93 to 1.75) | 1.17<br>(0.99 to 1.35) |
| Belarus | 2035<br>(1641 to 2510) | 16.86<br>(13.58 to 20.87) | 3411<br>(2711 to 4249) | 23.47<br>(18.38 to 29.39) | 0.68<br>(0.44 to 0.97) | 1.45<br>(1.12 to 1.78) |
| Belgium | 8023<br>(6630 to 9615) | 51.77<br>(43.16 to 61.8) | 14434<br>(11427 to 17561) | 54.14<br>(43.65 to 65.07) | 0.8<br>(0.56 to 1.07) | -0.05<br>(-0.2 to 0.1) |
| Belize | 127<br>(104 to 155) | 131.16<br>(108.01 to 159.36) | 667<br>(532 to 817) | 231.79<br>(187.31 to 281.44) | 4.26<br>(3.43 to 5.28) | 2.17<br>(1.75 to 2.59) |
| Benin | 5919<br>(4757 to 7315) | 284.36<br>(230.16 to 344.4) | 13567<br>(9881 to 18335) | 259.06<br>(194.8 to 338.01) | 1.29<br>(0.7 to 2.02) | -0.24<br>(-0.29 to -0.18) |
| Bermuda | 63<br>(52 to 76) | 101.8<br>(84.41 to 121.96) | 121<br>(98 to 148) | 97.23<br>(78.71 to 118.97) | 0.92<br>(0.61 to 1.29) | 0.12<br>(0 to 0.24) |
| Bhutan | 350<br>(232 to 506) | 128.86<br>(85.86 to 184.6) | 846<br>(612 to 1142) | 148.73<br>(106.96 to 198.79) | 1.42<br>(0.69 to 2.47) | 0.59<br>(0.51 to 0.67) |

| | | | | | | |
|---|---|---|---|---|---|---|
| Bolivia (Plurinational State of) | 6177 (4754 to 7954) | 198.52 (153.6 to 255.27) | 23401 (17454 to 31597) | 280.74 (209.42 to 371.88) | 2.79 (1.92 to 3.85) | 1.28 (1.17 to 1.38) |
| Bosnia and Herzegovina | 1954 (1564 to 2426) | 49.23 (40.01 to 60.5) | 3340 (2554 to 4386) | 58.59 (45.28 to 76.05) | 0.71 (0.37 to 1.15) | 0.77 (0.19 to 1.36) |
| Botswana | 1446 (1018 to 2045) | 246.4 (174.18 to 343.73) | 4422 (3018 to 6137) | 309.96 (215.98 to 423.6) | 2.06 (1.07 to 3.35) | 0.38 (0.03 to 0.72) |
| Brazil | 96119 (79577 to 113954) | 107.04 (90.04 to 125.59) | 252237 (213713 to 298127) | 107.92 (91.31 to 127.25) | 1.62 (1.38 to 1.9) | 0.02 (-0.08 to 0.11) |
| Brunei Darussalam | 175 (139 to 222) | 188.01 (149.07 to 235.4) | 457 (357 to 587) | 180.86 (143.06 to 226.1) | 1.62 (1.15 to 2.23) | 0.47 (0.23 to 0.71) |
| Bulgaria | 4990 (4006 to 6180) | 43.16 (35.09 to 52.69) | 11114 (8540 to 14368) | 83.07 (64.2 to 106.61) | 1.23 (0.76 to 1.74) | 2.44 (2.15 to 2.74) |
| Burkina Faso | 10003 (7783 to 12461) | 236.58 (187.18 to 289.49) | 22811 (17485 to 29124) | 242.68 (192.38 to 301.55) | 1.28 (0.79 to 1.89) | -0.01 (-0.13 to 0.11) |
| Burundi | 4974 (3644 to 6564) | 208.1 (153.43 to 272.74) | 7768 (5762 to 10429) | 171.45 (129.22 to 225.89) | 0.56 (0.17 to 1.08) | -0.95 (-1.06 to -0.84) |
| Cabo Verde | 314 (262 to 378) | 132.17 (110.55 to 159) | 840 (697 to 1000) | 190.86 (159.36 to 225.44) | 1.67 (1.14 to 2.25) | 0.57 (0.3 to 0.85) |
| Cambodia | 11681 (8907 to 14848) | 217.19 (169.22 to 272.73) | 23093 (17986 to 29225) | 183.14 (144.85 to 225.98) | 0.98 (0.48 to 1.58) | -0.68 (-0.77 to -0.6) |
| Cameroon | 17550 (13363 to 22520) | 380.46 (292.19 to 482.39) | 44680 (31484 to 61061) | 341.18 (251.71 to 452.04) | 1.55 (0.85 to 2.56) | -0.47 (-0.75 to -0.19) |

| | | | | | | |
|---|---|---|---|---|---|---|
| Canada | 19131 (16208 to 22168) | 59.78 (50.59 to 69.06) | 48870 (40585 to 56789) | 67.68 (56.98 to 78.7) | 1.55 (1.29 to 1.87) | 0.34 (0.27 to 0.41) |
| Central African Republic | 2843 (2071 to 3769) | 235.34 (173.85 to 304.22) | 5249 (3726 to 7300) | 231.37 (166.44 to 314.87) | 0.85 (0.38 to 1.49) | -0.03 (-0.14 to 0.09) |
| Chad | 7575 (5523 to 10814) | 262.54 (189.99 to 372.47) | 15477 (11315 to 21069) | 257.8 (189.91 to 342.55) | 1.04 (0.57 to 1.64) | -0.02 (-0.12 to 0.07) |
| Chile | 9681 (8063 to 11509) | 101.29 (85.41 to 119.63) | 30972 (25829 to 36768) | 129.93 (108.33 to 154.01) | 2.2 (1.82 to 2.61) | 1.15 (0.83 to 1.46) |
| China | 885572 (738270 to 1057438) | 101.36 (85.27 to 118.83) | 1692128 (1393406 to 2014210) | 88.48 (73.38 to 104.46) | 0.91 (0.59 to 1.31) | 0 (-0.14 to 0.15) |
| Colombia | 21491 (17881 to 26194) | 121.56 (101.07 to 146.67) | 58764 (45408 to 76057) | 110.25 (85.05 to 143.43) | 1.73 (1.19 to 2.3) | -0.33 (-0.42 to -0.24) |
| Comoros | 412 (241 to 563) | 188.74 (118.04 to 252.86) | 826 (625 to 1054) | 173.88 (134.25 to 220.94) | 1.01 (0.47 to 2.28) | -0.46 (-0.6 to -0.33) |
| Congo | 2816 (1964 to 3750) | 261.99 (180.28 to 342.15) | 5581 (3886 to 7613) | 218.87 (151.83 to 292.1) | 0.98 (0.48 to 1.71) | -0.79 (-0.9 to -0.67) |
| Cook Islands | 24 (19 to 30) | 189.25 (152.32 to 230.76) | 56 (46 to 69) | 237.41 (192.5 to 291.49) | 1.32 (0.88 to 1.9) | 0.95 (0.88 to 1.02) |
| Costa Rica | 1996 (1625 to 2432) | 110.2 (90.82 to 132.95) | 8709 (6770 to 11277) | 169.05 (131.44 to 218.26) | 3.36 (2.54 to 4.37) | 1.46 (1.13 to 1.78) |
| Côte d'Ivoire | 14475 (10970 to 18664) | 335.07 (261.38 to 417.63) | 32712 (23826 to 43142) | 284.75 (218.51 to 364.49) | 1.26 (0.64 to 2.04) | -0.64 (-0.86 to -0.42) |

| | | | | | | |
|---|---|---|---|---|---|---|
| Croatia | 2740<br>(2218 to 3285) | 44.63<br>(36.28 to 53.5) | 5028<br>(3892 to 6367) | 56.26<br>(44.19 to 70.49) | 0.84<br>(0.49 to 1.27) | 2.92<br>(2.03 to 3.82) |
| Cuba | 5800<br>(4812 to 7012) | 55.52<br>(46.18 to 67.26) | 17949<br>(14128 to 22265) | 98.54<br>(77.87 to 122.8) | 2.09<br>(1.56 to 2.68) | 2.4<br>(2.17 to 2.63) |
| Cyprus | 853<br>(648 to 1116) | 137.37<br>(104.75 to 175.62) | 1530<br>(1219 to 1916) | 87.38<br>(70.18 to 108.64) | 0.79<br>(0.48 to 1.18) | -1.84<br>(-2 to -1.68) |
| Czechia | 6459<br>(5242 to 7983) | 47.95<br>(39.03 to 58.94) | 7836<br>(6204 to 9883) | 38.42<br>(30.25 to 47.89) | 0.21<br>(-0.01 to 0.46) | -0.64<br>(-0.74 to -0.55) |
| Democratic People's Republic of Korea | 23507<br>(17686 to 30693) | 140.55<br>(108.67 to 178.44) | 42368<br>(33151 to 53707) | 133.5<br>(105.08 to 167.73) | 0.8<br>(0.42 to 1.29) | -0.1<br>(-0.2 to 0.01) |
| Democratic Republic of the Congo | 32076<br>(24881 to 41196) | 209.91<br>(167.97 to 260.39) | 62783<br>(47144 to 81989) | 174.54<br>(130.62 to 228.68) | 0.96<br>(0.46 to 1.56) | -0.74<br>(-0.78 to -0.7) |
| Denmark | 2360<br>(1929 to 2880) | 29.2<br>(23.91 to 35.38) | 5564<br>(4419 to 6734) | 46.4<br>(37.24 to 55.52) | 1.36<br>(1.06 to 1.67) | 1.1<br>(0.85 to 1.36) |
| Djibouti | 228<br>(160 to 320) | 159.3<br>(113.43 to 218.5) | 1047<br>(756 to 1479) | 186.14<br>(137.17 to 253.27) | 3.6<br>(2.29 to 5.36) | 0.45<br>(0.35 to 0.55) |
| Dominica | 140<br>(115 to 172) | 197.38<br>(162.7 to 244.63) | 232<br>(184 to 295) | 262.52<br>(209.07 to 333.23) | 0.66<br>(0.35 to 1.06) | 1.26<br>(1.16 to 1.36) |
| Dominican Republic | 3364<br>(2730 to 4131) | 86.32<br>(71.08 to 105.24) | 14077<br>(10384 to 19174) | 150.06<br>(111 to 202.85) | 3.18<br>(2.21 to 4.52) | 2.72<br>(2.44 to 3.01) |
| Ecuador | 5980<br>(4988 to 7195) | 111.66<br>(93.21 to 133.38) | 36870<br>(28505 to 48009) | 255.59<br>(198.4 to 329.07) | 5.17<br>(3.95 to 6.52) | 3.22<br>(2.6 to 3.85) |
| Egypt | 67869<br>(47115 to 87803) | 253.03<br>(175.06 to 325.71) | 167205<br>(104137 to 239958) | 289.39<br>(181.46 to 407.25) | 1.46<br>(0.8 to 2.26) | 0.57<br>(0.48 to 0.66) |

| | | | | | | |
|---|---|---|---|---|---|---|
| El Salvador | 3948<br>(3272 to 4876) | 127.37<br>(104.86 to 156.08) | 22420<br>(16459 to 30282) | 376.8<br>(275.52 to 511.58) | 4.68<br>(3.3 to 6.36) | 4.11<br>(3.41 to 4.82) |
| Equatorial Guinea | 427<br>(298 to 575) | 214.2<br>(151.75 to 285.08) | 1032<br>(728 to 1480) | 211.91<br>(150.32 to 295.59) | 1.42<br>(0.67 to 2.63) | 0.08<br>(-0.12 to 0.28) |
| Eritrea | 1831<br>(1189 to 2550) | 174.27<br>(111.21 to 249.14) | 5108<br>(3526 to 7151) | 188.24<br>(129.95 to 254.46) | 1.79<br>(1.05 to 2.91) | 0.12<br>(-0.04 to 0.27) |
| Estonia | 419<br>(339 to 509) | 21.8<br>(17.63 to 26.48) | 1719<br>(1343 to 2211) | 61.88<br>(48.55 to 78.37) | 3.1<br>(2.28 to 4.19) | 4.29<br>(3.92 to 4.66) |
| Eswatini | 975<br>(739 to 1267) | 318.07<br>(243.08 to 408.04) | 2552<br>(1758 to 3474) | 419.09<br>(298.1 to 557.88) | 1.62<br>(0.85 to 2.72) | 1.15<br>(0.57 to 1.73) |
| Ethiopia | 57217<br>(44193 to 73751) | 277.24<br>(216.25 to 346.46) | 72212<br>(58986 to 87349) | 173.61<br>(141.72 to 208.6) | 0.26<br>(-0.01 to 0.61) | -1.74<br>(-1.85 to -1.63) |
| Fiji | 1038<br>(804 to 1335) | 265.99<br>(208.27 to 339.82) | 2549<br>(1921 to 3280) | 348.62<br>(268.23 to 442.43) | 1.46<br>(0.87 to 2.26) | 0.51<br>(0.19 to 0.83) |
| Finland | 1220<br>(968 to 1503) | 17.68<br>(14.15 to 21.52) | 3029<br>(2353 to 3753) | 23.23<br>(18.3 to 28.48) | 1.48<br>(1.12 to 1.91) | -0.32<br>(-0.97 to 0.33) |
| France | 39953<br>(32629 to 47569) | 45.84<br>(38.06 to 54.24) | 64875<br>(52675 to 77515) | 39.92<br>(32.5 to 47.56) | 0.62<br>(0.41 to 0.86) | -0.23<br>(-0.32 to -0.14) |
| Gabon | 1355<br>(960 to 1758) | 245.59<br>(173.37 to 318.01) | 2710<br>(1761 to 3695) | 270.51<br>(173 to 361.4) | 1<br>(0.41 to 1.71) | 0.29<br>(0.09 to 0.48) |
| Gambia | 961<br>(691 to 1297) | 261.06<br>(196.14 to 339.31) | 2799<br>(2122 to 3700) | 274.57<br>(212.03 to 352.52) | 1.91<br>(1.04 to 3.14) | 0.12<br>(-0.02 to 0.25) |
| Georgia | 1406<br>(1110 to 1770) | 24.01<br>(19.07 to 30.1) | 2307<br>(1816 to 2891) | 41.48<br>(32.28 to 52.26) | 0.64<br>(0.35 to 0.97) | 1.82<br>(1.37 to 2.28) |

| | | | | | | |
|---|---|---|---|---|---|---|
| Germany | 50001 (40451 to 61053) | 38.98 (31.82 to 47.24) | 134881 (104238 to 168946) | 60.72 (47.67 to 74.23) | 1.7 (1.27 to 2.24) | 2.49 (2.23 to 2.76) |
| Ghana | 14104 (10088 to 19098) | 215.85 (156.45 to 285.09) | 38903 (27169 to 51874) | 233.43 (164.99 to 300.43) | 1.76 (0.92 to 2.71) | 0.4 (0.33 to 0.47) |
| Greece | 13542 (10650 to 16651) | 91.86 (73.42 to 111.6) | 19813 (15721 to 24501) | 70.71 (57.13 to 86.13) | 0.46 (0.27 to 0.66) | -1.04 (-1.45 to -0.63) |
| Greenland | 26 (21 to 32) | 91.27 (74.66 to 109.74) | 59 (47 to 72) | 98.89 (79.32 to 120.89) | 1.24 (0.81 to 1.71) | 0.4 (0.3 to 0.49) |
| Grenada | 140 (114 to 169) | 193.24 (157.73 to 236.49) | 293 (237 to 360) | 267.78 (220.33 to 323.93) | 1.1 (0.78 to 1.47) | 1.48 (1.34 to 1.62) |
| Guam | 143 (117 to 174) | 188.99 (157.6 to 225.48) | 480 (378 to 595) | 254.53 (201.55 to 313.65) | 2.34 (1.76 to 3.1) | 1.27 (1.06 to 1.48) |
| Guatemala | 6189 (4964 to 7789) | 174.36 (142.43 to 213.62) | 33165 (25375 to 43933) | 298.81 (228.18 to 392.98) | 4.36 (3.22 to 5.83) | 2.58 (2.21 to 2.95) |
| Guinea | 10159 (7702 to 13415) | 304.42 (231.83 to 402.87) | 16240 (12094 to 21207) | 277.21 (209.14 to 355.77) | 0.6 (0.2 to 1.13) | 0.14 (0 to 0.28) |
| Guinea-Bissau | 1869 (1386 to 2447) | 427.26 (326.34 to 541.53) | 2761 (2065 to 3651) | 337.14 (255.15 to 432.71) | 0.48 (0.09 to 0.99) | -0.75 (-0.79 to -0.71) |
| Guyana | 660 (525 to 821) | 165.48 (131.63 to 204.49) | 1700 (1274 to 2230) | 266.47 (201.37 to 344.13) | 1.57 (1 to 2.29) | 2.32 (2.11 to 2.53) |

| | | | | | | |
|---|---|---|---|---|---|---|
| Haiti | 6144<br>(4321 to 9038) | 183.46<br>(129.06 to 277.19) | 14106<br>(9748 to 21495) | 195.41<br>(136.57 to 300.61) | 1.3<br>(0.75 to 1.97) | 0.59<br>(0.45 to 0.73) |
| Honduras | 3205<br>(2420 to 4469) | 149.19<br>(110.75 to 215.65) | 18307<br>(13830 to 24627) | 305.7<br>(231.48 to 409.08) | 4.71<br>(3.54 to 6.16) | 2.9<br>(2.65 to 3.16) |
| Hungary | 5097<br>(4201 to 6130) | 36.69<br>(30.53 to 44.03) | 9973<br>(7940 to 12416) | 52.17<br>(41.37 to 64.37) | 0.96<br>(0.64 to 1.32) | 1.98<br>(1.63 to 2.33) |
| Iceland | 89<br>(73 to 105) | 30.06<br>(24.87 to 35.51) | 200<br>(161 to 238) | 33.3<br>(27.01 to 39.9) | 1.25<br>(0.98 to 1.55) | 0.78<br>(0.52 to 1.04) |
| India | 623007<br>(501557 to 775325) | 135.05<br>(109.68 to 165.58) | 1394020<br>(1111090 to 1716100) | 120.59<br>(97.2 to 147.93) | 1.24<br>(0.85 to 1.74) | -0.49<br>(-0.67 to -0.32) |
| Indonesia | 231732<br>(190494 to 278922) | 197.76<br>(164.71 to 235.94) | 459213<br>(375974 to 567784) | 198.01<br>(164.17 to 239.21) | 0.98<br>(0.62 to 1.38) | 0.02<br>(-0.08 to 0.13) |
| Iran (Islamic Republic of) | 38311<br>(31891 to 46021) | 157.16<br>(132.22 to 187.39) | 92636<br>(79012 to 107257) | 133.05<br>(113.68 to 153.27) | 1.42<br>(1.1 to 1.72) | -0.74<br>(-0.86 to -0.62) |
| Iraq | 22314<br>(16976 to 29882) | 291.37<br>(221.77 to 393.42) | 58755<br>(43660 to 76188) | 270.72<br>(204.15 to 346.28) | 1.63<br>(1.01 to 2.31) | -0.33<br>(-0.38 to -0.28) |
| Ireland | 2298<br>(1841 to 2823) | 58.03<br>(46.83 to 70.3) | 3522<br>(2831 to 4292) | 46.57<br>(37.63 to 56.77) | 0.53<br>(0.31 to 0.76) | -1.01<br>(-1.14 to -0.87) |
| Israel | 5336<br>(4320 to 6404) | 115.04<br>(94.32 to 136.32) | 13536<br>(10909 to 16375) | 110.3<br>(88.95 to 133.1) | 1.54<br>(1.22 to 1.88) | 0.03<br>(-0.16 to 0.21) |
| Italy | 50547<br>(42267 to 59296) | 57.58<br>(48.29 to 67.02) | 80197<br>(66015 to 94101) | 47.68<br>(39.4 to 55.46) | 0.59<br>(0.39 to 0.78) | -0.47<br>(-0.61 to -0.33) |

| | | | | | | |
|---|---|---|---|---|---|---|
| Jamaica | 2378 (1976 to 2832) | 130.6 (109.2 to 155) | 5369 (4197 to 6804) | 177.53 (138.58 to 226.66) | 1.26 (0.84 to 1.81) | 0.64 (0.23 to 1.06) |
| Japan | 116094 (96531 to 138373) | 72.04 (59.98 to 85.14) | 224116 (176825 to 269013) | 55.1 (44.76 to 64.94) | 0.93 (0.68 to 1.16) | -1.44 (-1.74 to -1.15) |
| Jordan | 3004 (2356 to 3714) | 246.28 (192.68 to 305.88) | 13807 (10988 to 16979) | 233.57 (188.38 to 280.83) | 3.6 (2.66 to 4.68) | -0.08 (-0.2 to 0.03) |
| Kazakhstan | 3695 (2931 to 4648) | 26.97 (21.65 to 33.78) | 7230 (5734 to 9197) | 42.36 (33.56 to 53.76) | 0.96 (0.66 to 1.29) | 1.45 (1.28 to 1.62) |
| Kenya | 10720 (8531 to 13793) | 130.6 (103.52 to 166.16) | 34337 (27283 to 42931) | 158.97 (127.8 to 196.59) | 2.2 (1.68 to 2.79) | 0.75 (0.66 to 0.84) |
| Kiribati | 145 (112 to 188) | 351.7 (274.78 to 442.83) | 329 (239 to 441) | 440.05 (322.88 to 574.65) | 1.27 (0.67 to 2.02) | 0.54 (0.18 to 0.91) |
| Kuwait | 1099 (902 to 1326) | 182.15 (149.53 to 219.57) | 2902 (2311 to 3562) | 117.56 (94.16 to 143.21) | 1.64 (1.22 to 2.12) | -1.8 (-2.41 to -1.18) |
| Kyrgyzstan | 1323 (1019 to 1730) | 38.99 (30.2 to 50.03) | 1920 (1476 to 2480) | 36.5 (28.58 to 46.8) | 0.45 (0.23 to 0.69) | -0.6 (-0.9 to -0.3) |
| Lao People's Democratic Republic | 10512 (7658 to 14319) | 462.33 (342.46 to 625.38) | 17001 (12674 to 22520) | 364.02 (277.72 to 472.02) | 0.62 (0.2 to 1.27) | -0.97 (-1.02 to -0.92) |
| Latvia | 553 (435 to 681) | 16.61 (13.01 to 20.5) | 1259 (968 to 1587) | 33.57 (25.95 to 42.22) | 1.28 (0.92 to 1.72) | 3.24 (2.55 to 3.93) |
| Lebanon | 4386 (3419 to 5673) | 207.62 (163.83 to 265.19) | 8388 (6191 to 10880) | 161.67 (118.85 to 209.51) | 0.91 (0.43 to 1.48) | -0.81 (-0.94 to -0.67) |

| | | | | | | |
|---|---|---|---|---|---|---|
| Lesotho | 2237 (1737 to 2866) | 225.33 (175.52 to 287.86) | 5433 (3776 to 7465) | 418.15 (298.54 to 557.66) | 1.43 (0.69 to 2.33) | 2.76 (2.46 to 3.05) |
| Liberia | 3626 (2822 to 4716) | 331.1 (261.68 to 426.49) | 5856 (4138 to 8268) | 264.81 (193.31 to 368.65) | 0.62 (0.14 to 1.22) | -0.58 (-0.84 to -0.32) |
| Libya | 3584 (2687 to 4729) | 196.4 (146.3 to 261.48) | 10243 (7163 to 13807) | 207.55 (146.35 to 280.47) | 1.86 (1.1 to 2.72) | 0.4 (0.26 to 0.53) |
| Lithuania | 862 (695 to 1042) | 19.91 (16.06 to 24.08) | 1541 (1234 to 1925) | 29 (23.16 to 35.9) | 0.79 (0.5 to 1.16) | 1 (0.62 to 1.37) |
| Luxembourg | 273 (219 to 334) | 51.39 (41.68 to 62.47) | 550 (435 to 674) | 50.52 (40.43 to 61.72) | 1.01 (0.69 to 1.34) | 0.13 (-0.07 to 0.32) |
| Madagascar | 8034 (5960 to 10505) | 152.89 (112.2 to 201.34) | 15325 (11286 to 20804) | 140.65 (103.31 to 188.75) | 0.91 (0.43 to 1.48) | -0.37 (-0.45 to -0.3) |
| Malawi | 6792 (5350 to 8526) | 179.68 (142.05 to 221.37) | 12856 (9965 to 16368) | 174.56 (136.21 to 216.02) | 0.89 (0.48 to 1.41) | -0.2 (-0.35 to -0.05) |
| Malaysia | 21679 (18026 to 25757) | 221.62 (185.15 to 260.73) | 59963 (46875 to 75717) | 224.15 (175.81 to 281.82) | 1.77 (1.21 to 2.39) | -0.37 (-0.55 to -0.19) |
| Maldives | 425 (334 to 543) | 465.51 (377.64 to 581.02) | 816 (654 to 988) | 265.4 (214.15 to 319.05) | 0.92 (0.45 to 1.42) | -2.3 (-2.57 to -2.04) |
| Mali | 12804 (9723 to 17161) | 305.59 (236.91 to 402.48) | 22663 (17083 to 29930) | 251.13 (193.68 to 323.23) | 0.77 (0.34 to 1.35) | -0.57 (-0.73 to -0.4) |

| | | | | | | |
|---|---|---|---|---|---|---|
| Malta | 267<br>(212 to 330) | 66.48<br>(52.63 to 81.65) | 518<br>(409 to 642) | 53.86<br>(42.94 to 66.31) | 0.94<br>(0.65 to 1.28) | -0.74<br>(-0.92 to -0.55) |
| Marshall Islands | 57<br>(43 to 77) | 315.66<br>(236.79 to 425.37) | 156<br>(109 to 216) | 418.21<br>(300.23 to 574.68) | 1.73<br>(1.09 to 2.48) | 0.99<br>(0.76 to 1.23) |
| Mauritania | 3961<br>(3056 to 4947) | 387.2<br>(302 to 477.37) | 5075<br>(3706 to 6647) | 242.09<br>(179.79 to 312.28) | 0.28<br>(-0.04 to 0.66) | -1.57<br>(-1.66 to -1.47) |
| Mauritius | 3040<br>(2527 to 3593) | 396.45<br>(333.39 to 465.58) | 11101<br>(8642 to 13948) | 635.53<br>(496.72 to 794.12) | 2.65<br>(1.92 to 3.59) | 1.93<br>(1.59 to 2.28) |
| Mexico | 73476<br>(62435 to 86191) | 169.69<br>(144.98 to 196.71) | 430226<br>(345558 to 527876) | 365.93<br>(294.2 to 448.22) | 4.86<br>(4.05 to 5.76) | 3.01<br>(2.58 to 3.43) |
| Micronesia (Federated States of) | 196<br>(146 to 258) | 391.31<br>(297.71 to 514.78) | 436<br>(305 to 599) | 592.52<br>(429.89 to 782.39) | 1.23<br>(0.54 to 2.1) | 1.39<br>(1.05 to 1.74) |
| Monaco | 20<br>(16 to 25) | 27.24<br>(21.3 to 33.47) | 37<br>(30 to 47) | 35.37<br>(28.55 to 43.58) | 0.86<br>(0.55 to 1.24) | 1.17<br>(0.9 to 1.44) |
| Mongolia | 1038<br>(803 to 1342) | 94.36<br>(73.86 to 122.2) | 1401<br>(1053 to 1864) | 56.82<br>(44.02 to 74.05) | 0.35<br>(0.04 to 0.73) | -2.68<br>(-3.05 to -2.31) |
| Montenegro | 444<br>(364 to 552) | 73<br>(59.91 to 90.71) | 777<br>(600 to 978) | 82.66<br>(64.43 to 103.31) | 0.75<br>(0.4 to 1.11) | 1.43<br>(1.09 to 1.77) |
| Morocco | 23876<br>(18645 to 31866) | 183.91<br>(143.93 to 251.98) | 65737<br>(49401 to 84433) | 227.75<br>(174.8 to 293.99) | 1.75<br>(1.05 to 2.5) | 0.78<br>(0.67 to 0.88) |
| Mozambique | 8401<br>(6375 to 11156) | 148.33<br>(115.01 to 194.9) | 19171<br>(14448 to 25348) | 171.42<br>(132.03 to 221.93) | 1.28<br>(0.61 to 2.07) | 0.6<br>(0.45 to 0.75) |

| | | | | | | |
|---|---|---|---|---|---|---|
| Myanmar | 78045 (56578 to 104661) | 290.63 (216.34 to 382.04) | 117090 (92502 to 150155) | 243.76 (196.63 to 307.42) | 0.5 (0.11 to 1.06) | -0.68 (-0.8 to -0.57) |
| Namibia | 1792 (1284 to 2539) | 250.26 (182.95 to 353.35) | 3364 (2318 to 4757) | 238.18 (168.09 to 334.23) | 0.88 (0.36 to 1.53) | -0.45 (-0.78 to -0.12) |
| Nauru | 17 (12 to 23) | 380.66 (281.91 to 496.67) | 25 (18 to 34) | 514.66 (373.89 to 671.33) | 0.52 (0.18 to 0.93) | 0.92 (0.6 to 1.25) |
| Nepal | 11138 (7869 to 15396) | 109.75 (78.86 to 151.87) | 31760 (22832 to 42728) | 142.36 (103.27 to 191.2) | 1.85 (1.06 to 2.79) | 0.91 (0.63 to 1.18) |
| Netherlands | 9648 (7918 to 11437) | 47.97 (39.54 to 56.65) | 19422 (15827 to 23496) | 53.3 (43.82 to 64.04) | 1.01 (0.77 to 1.27) | 0.99 (0.83 to 1.16) |
| New Zealand | 1507 (1227 to 1851) | 39.32 (32.16 to 47.71) | 3996 (3237 to 4788) | 49.7 (40.48 to 59.67) | 1.65 (1.35 to 2.01) | 0.87 (0.74 to 1) |
| Nicaragua | 3508 (2835 to 4321) | 218.79 (176.07 to 268.74) | 20204 (15186 to 25916) | 457.5 (346.17 to 581.8) | 4.76 (3.66 to 6.05) | 2.95 (2.61 to 3.28) |
| Niger | 8427 (6353 to 11357) | 288.16 (218.08 to 380.5) | 18990 (13879 to 25816) | 234.07 (176.21 to 308.2) | 1.25 (0.74 to 2) | -0.73 (-0.81 to -0.64) |
| Nigeria | 92119 (71676 to 119295) | 212.91 (168.47 to 275.48) | 160341 (121962 to 208010) | 181.02 (141.78 to 229.82) | 0.74 (0.3 to 1.29) | -0.41 (-0.53 to -0.29) |
| Niue | 6 (5 to 8) | 279.25 (209.88 to 356.04) | 8 (6 to 10) | 369.97 (267.72 to 483.43) | 0.29 (0.01 to 0.63) | 0.85 (0.61 to 1.09) |
| North Macedonia | 1427 (1139 to 1734) | 77.62 (62.61 to 93.53) | 2574 (1995 to 3356) | 84.74 (66.17 to 108.81) | 0.8 (0.43 to 1.27) | 0.53 (0.15 to 0.92) |

| | | | | | | |
|---|---|---|---|---|---|---|
| Northern Mariana Islands | 74 (57 to 95) | 352.55 (283.51 to 432.5) | 222 (172 to 278) | 435.28 (349.88 to 529.82) | 2 (1.31 to 2.81) | 0.89 (0.8 to 0.99) |
| Norway | 2276 (1910 to 2673) | 32.43 (27.27 to 37.91) | 3995 (3332 to 4670) | 38.4 (32.05 to 44.79) | 0.76 (0.61 to 0.94) | 1.13 (0.78 to 1.49) |
| Oman | 694 (524 to 929) | 115.3 (87.38 to 152.89) | 2087 (1641 to 2598) | 135.63 (111.58 to 162.41) | 2.01 (1.28 to 2.85) | 0.9 (0.77 to 1.03) |
| Pakistan | 90178 (64899 to 124002) | 152.34 (108.94 to 211.46) | 266139 (189370 to 358926) | 222.18 (159.96 to 293.29) | 1.95 (1.25 to 2.83) | 1.35 (1.09 to 1.62) |
| Palau | 44 (33 to 57) | 423.15 (326.92 to 547.49) | 114 (86 to 150) | 535.49 (414.22 to 687.28) | 1.61 (0.94 to 2.44) | 0.77 (0.56 to 0.97) |
| Palestine | 2591 (1957 to 3333) | 306.11 (234.28 to 390.57) | 5024 (4078 to 6160) | 225.01 (180.8 to 274.29) | 0.94 (0.51 to 1.49) | -0.79 (-1.03 to -0.54) |
| Panama | 1495 (1230 to 1836) | 96.19 (79.82 to 117.66) | 6820 (5280 to 8849) | 163.63 (126.19 to 213.18) | 3.56 (2.72 to 4.63) | 1.92 (1.61 to 2.23) |
| Papua New Guinea | 2331 (1800 to 3016) | 105.66 (82.74 to 133.39) | 7279 (5355 to 9750) | 126.67 (94.91 to 166.61) | 2.12 (1.41 to 3) | 0.59 (0.48 to 0.7) |
| Paraguay | 1972 (1603 to 2391) | 88.03 (71.59 to 105.9) | 10567 (7939 to 13939) | 190.98 (144.4 to 252.47) | 4.36 (3.16 to 5.89) | 3.26 (3.05 to 3.47) |
| Peru | 15046 (12032 to 18870) | 126.66 (101.82 to 158.21) | 43327 (32531 to 56916) | 134.2 (100.8 to 176.65) | 1.88 (1.17 to 2.74) | 0.31 (0.09 to 0.54) |
| Philippines | 100771 (83020 to 120773) | 314.36 (262.91 to 370.15) | 339663 (263727 to 438671) | 406.98 (320.98 to 518) | 2.37 (1.71 to 3.15) | 1.29 (1.03 to 1.56) |
| Poland | 27287 (22592 to 32634) | 63.5 (52.86 to 75.81) | 28526 (23297 to 34997) | 41.85 (33.95 to 51.09) | 0.05 (-0.12 to 0.23) | -0.59 (-1.07 to -0.12) |

| | | | | | | |
|---|---|---|---|---|---|---|
| Portugal | 8423<br>(6811 to 10374) | 64.13<br>(52.3 to 78.4) | 17703<br>(13898 to 21943) | 63.63<br>(51.36 to 78.13) | 1.1<br>(0.79 to 1.44) | 0.24<br>(-0.22 to 0.71) |
| Puerto Rico | 5698<br>(4772 to 6773) | 158.89<br>(133.01 to 188.34) | 11358<br>(8878 to 14238) | 164.54<br>(128.7 to 206.23) | 0.99<br>(0.62 to 1.46) | 0.63<br>(0.41 to 0.86) |
| Qatar | 229<br>(172 to 325) | 277.21<br>(205.38 to 446.38) | 1527<br>(1161 to 1978) | 258.14<br>(203.79 to 324.77) | 5.65<br>(3.78 to 7.71) | 0<br>(-0.25 to 0.26) |
| Republic of Korea | 20383<br>(17385 to 24760) | 70.25<br>(60.94 to 83.04) | 44898<br>(38039 to 52690) | 52.35<br>(44.5 to 61.24) | 1.2<br>(0.87 to 1.6) | -0.61<br>(-0.8 to -0.43) |
| Republic of Moldova | 728<br>(586 to 892) | 18.22<br>(14.73 to 22.17) | 1530<br>(1233 to 1872) | 28.28<br>(22.63 to 34.38) | 1.1<br>(0.81 to 1.42) | 1.49<br>(1.25 to 1.74) |
| Romania | 10866<br>(9394 to 12436) | 40.92<br>(35.72 to 46.66) | 17809<br>(13934 to 22424) | 49.57<br>(38.43 to 61.89) | 0.64<br>(0.33 to 1) | -0.02<br>(-0.53 to 0.5) |
| Russian Federation | 60226<br>(50089 to 72571) | 35.17<br>(29.38 to 42.17) | 78702<br>(63752 to 95112) | 36.3<br>(29.55 to 43.72) | 0.31<br>(0.16 to 0.48) | -0.49<br>(-0.99 to 0.01) |
| Rwanda | 6370<br>(4908 to 8140) | 216.83<br>(170.01 to 273.23) | 9235<br>(7044 to 11890) | 160.3<br>(124.58 to 203.06) | 0.45<br>(0.12 to 0.9) | -1.77<br>(-2.06 to -1.49) |
| Saint Kitts and Nevis | 96<br>(78 to 116) | 262.58<br>(217.84 to 317.21) | 191<br>(147 to 242) | 297.43<br>(234.94 to 370.69) | 1<br>(0.62 to 1.47) | 0.74<br>(0.59 to 0.89) |
| Saint Lucia | 150<br>(123 to 184) | 171.94<br>(141.72 to 208.56) | 419<br>(337 to 523) | 197.97<br>(159.68 to 245.8) | 1.79<br>(1.33 to 2.35) | 0.59<br>(0.41 to 0.78) |
| Saint Vincent and the Grenadines | 103<br>(84 to 125) | 141.54<br>(116.75 to 172.61) | 268<br>(218 to 334) | 203.7<br>(166.58 to 252.85) | 1.6<br>(1.22 to 2.07) | 1.51<br>(1.34 to 1.67) |
| Samoa | 265<br>(200 to 346) | 295.34<br>(224.22 to 384.48) | 523<br>(403 to 669) | 350.94<br>(274.38 to 445.22) | 0.97<br>(0.51 to 1.66) | 0.51<br>(0.31 to 0.72) |

| | | | | | | |
|---|---|---|---|---|---|---|
| San Marino | 8 (6 to 10) | 25.67 (20.34 to 31.47) | 21 (15 to 29) | 29.39 (21.61 to 39.52) | 1.63 (1.01 to 2.37) | 0.89 (0.73 to 1.04) |
| Sao Tome and Principe | 228 (175 to 278) | 361.12 (285.84 to 433.97) | 459 (352 to 589) | 421.96 (327.92 to 523.3) | 1.02 (0.57 to 1.65) | 0.39 (0.22 to 0.56) |
| Saudi Arabia | 18510 (13718 to 24648) | 324.21 (243.98 to 427.43) | 60596 (46068 to 77728) | 360.89 (283.05 to 452.77) | 2.27 (1.32 to 3.41) | 0.58 (0.37 to 0.79) |
| Senegal | 10987 (8209 to 14744) | 332.24 (256.7 to 442.49) | 21112 (15527 to 28329) | 274.6 (208.6 to 362.21) | 0.92 (0.44 to 1.55) | -0.56 (-0.72 to -0.41) |
| Serbia | 8452 (6692 to 10720) | 79.06 (63.1 to 99.48) | 14038 (10533 to 18599) | 89.28 (68.3 to 114.9) | 0.66 (0.29 to 1.12) | 1.29 (0.97 to 1.61) |
| Seychelles | 153 (129 to 181) | 267.07 (225.75 to 317.3) | 401 (330 to 483) | 371.24 (310.12 to 443.56) | 1.62 (1.25 to 2.02) | 0.68 (0.34 to 1.03) |
| Sierra Leone | 5136 (3957 to 6591) | 264.06 (205.46 to 334.37) | 9086 (6774 to 12123) | 233.62 (177.97 to 306.15) | 0.77 (0.34 to 1.35) | -0.22 (-0.3 to -0.13) |
| Singapore | 1994 (1642 to 2407) | 91.79 (75.71 to 110.17) | 5066 (4119 to 6094) | 67.41 (54.61 to 81.38) | 1.54 (1.15 to 1.92) | 0.19 (-0.61 to 1) |
| Slovakia | 4039 (3217 to 4944) | 68.39 (55.24 to 83.17) | 4671 (3627 to 5948) | 52.39 (41.06 to 66.42) | 0.16 (-0.08 to 0.44) | -1.23 (-1.8 to -0.65) |
| Slovenia | 559 (444 to 689) | 23.56 (18.69 to 29.06) | 1333 (1020 to 1708) | 29.97 (22.98 to 38.14) | 1.38 (0.82 to 2.06) | 1.33 (1.15 to 1.51) |
| Solomon Islands | 630 (457 to 886) | 366.82 (267.65 to 513) | 915 (700 to 1191) | 230.25 (181.37 to 292.27) | 0.45 (0.04 to 0.96) | -1.79 (-2.15 to -1.43) |
| Somalia | 5386 (3829 to 7262) | 211.56 (149.12 to 282.65) | 13829 (9869 to 19168) | 201.44 (145.86 to 272.79) | 1.57 (0.92 to 2.45) | -0.08 (-0.14 to -0.03) |

| | | | | | | |
|---|---|---|---|---|---|---|
| South Africa | 38706 (32344 to 46042) | 169.39 (142.46 to 199.09) | 104250 (87822 to 122361) | 231.46 (197.09 to 273.26) | 1.69 (1.39 to 2.05) | 0.26 (-0.41 to 0.93) |
| South Sudan | 4243 (2943 to 5921) | 180.51 (125.08 to 250.06) | 6424 (4421 to 9159) | 176.72 (123.98 to 249.79) | 0.51 (0.11 to 1.11) | -0.13 (-0.17 to -0.08) |
| Spain | 35008 (28136 to 42764) | 65.97 (53.51 to 80.17) | 56699 (43287 to 71354) | 47.37 (36.97 to 58.77) | 0.62 (0.36 to 0.91) | -1.37 (-1.57 to -1.17) |
| Sri Lanka | 25071 (20676 to 30389) | 220.61 (183.4 to 261.91) | 52300 (38955 to 68859) | 211.14 (158.89 to 275.61) | 1.09 (0.58 to 1.8) | -0.04 (-0.33 to 0.24) |
| Sudan | 18713 (13865 to 26663) | 204.79 (152.65 to 296.61) | 37094 (26003 to 55367) | 204.51 (144.28 to 315.03) | 0.98 (0.35 to 1.88) | 0.33 (0.18 to 0.48) |
| Suriname | 427 (349 to 518) | 160.55 (131.33 to 195.02) | 1488 (1174 to 1872) | 250.89 (199.03 to 313.52) | 2.49 (1.82 to 3.17) | 1.57 (1.31 to 1.83) |
| Sweden | 3785 (3099 to 4562) | 24.37 (19.99 to 29.29) | 8444 (6825 to 10018) | 35.39 (28.87 to 41.61) | 1.23 (0.96 to 1.56) | 1.29 (1.15 to 1.43) |
| Switzerland | 3522 (2850 to 4281) | 32.62 (26.4 to 39.6) | 9592 (7427 to 11871) | 46.12 (36.24 to 56.53) | 1.72 (1.33 to 2.17) | 2.15 (1.85 to 2.45) |
| Syrian Arab Republic | 13009 (10072 to 16899) | 244.83 (190 to 318.55) | 21144 (15877 to 28221) | 190.21 (144.96 to 246.92) | 0.63 (0.2 to 1.21) | -1.48 (-1.76 to -1.19) |
| Taiwan (Province of China) | 27822 (23778 to 32329) | 187.28 (162.11 to 214.97) | 70042 (56111 to 86965) | 178.77 (143.45 to 221.95) | 1.52 (1.08 to 2.06) | -0.11 (-0.24 to 0.02) |
| Tajikistan | 516 (391 to 687) | 15.31 (11.97 to 19.61) | 1780 (1339 to 2337) | 33.18 (26.03 to 42.67) | 2.45 (2.01 to 3.01) | 3.08 (2.77 to 3.4) |

| | | | | | | |
|---|---|---|---|---|---|---|
| Thailand | 97609 (78692 to 125523) | 250.2 (205.93 to 308.45) | 246384 (182485 to 325591) | 248.25 (184.34 to 326.61) | 1.52 (0.84 to 2.41) | -0.13 (-0.25 to -0.01) |
| Timor-Leste | 939 (673 to 1315) | 283.26 (210.19 to 396) | 2289 (1602 to 3058) | 275.45 (199.2 to 361.4) | 1.44 (0.73 to 2.3) | -0.12 (-0.32 to 0.08) |
| Togo | 3811 (2916 to 4997) | 284 (220.97 to 366.57) | 9664 (7232 to 12750) | 251.16 (193.9 to 320.32) | 1.54 (0.87 to 2.35) | -0.4 (-0.43 to -0.37) |
| Tokelau | 3 (3 to 5) | 259.04 (193.32 to 348.93) | 4 (3 to 6) | 318.17 (239.6 to 426.77) | 0.23 (-0.09 to 0.6) | 0.66 (0.45 to 0.87) |
| Tonga | 128 (100 to 168) | 226.57 (179.48 to 293.4) | 260 (196 to 348) | 325.61 (247.02 to 433.98) | 1.03 (0.58 to 1.6) | 1.23 (0.9 to 1.56) |
| Trinidad and Tobago | 1074 (894 to 1299) | 127.15 (106.47 to 151.71) | 3751 (2770 to 4977) | 204.91 (152.46 to 268.84) | 2.49 (1.71 to 3.49) | 2.01 (1.77 to 2.26) |
| Tunisia | 7245 (5721 to 9060) | 154.25 (122.46 to 193.36) | 18801 (13969 to 25207) | 157.42 (116.53 to 209.11) | 1.6 (0.89 to 2.47) | -0.08 (-0.16 to 0.01) |
| Turkey | 74723 (56552 to 104297) | 217.63 (163.85 to 309.12) | 131450 (101783 to 164104) | 154.17 (119.44 to 192.38) | 0.76 (0.22 to 1.36) | -1.21 (-1.5 to -0.92) |
| Turkmenistan | 922 (732 to 1164) | 41 (32.97 to 51.04) | 2412 (1821 to 3163) | 56.27 (42.85 to 73.02) | 1.62 (1.17 to 2.18) | 0.78 (0.56 to 1.01) |
| Tuvalu | 21 (16 to 28) | 305.43 (230.08 to 396.38) | 39 (27 to 54) | 379.74 (271.4 to 520.92) | 0.81 (0.35 to 1.48) | 0.71 (0.52 to 0.91) |
| Uganda | 10274 (7544 to 13719) | 163.11 (121.15 to 216.85) | 23524 (17498 to 30967) | 166.64 (125.3 to 213.84) | 1.29 (0.72 to 2) | -0.18 (-0.32 to -0.03) |
| Ukraine | 10470 (8566 to 12833) | 16.42 (13.39 to 20.18) | 17437 (14015 to 21215) | 26.03 (20.85 to 32.19) | 0.67 (0.46 to 0.9) | 1.93 (1.7 to 2.15) |

| | | | | | | |
|---|---|---|---|---|---|---|
| United Arab Emirates | 1683 (1185 to 2215) | 385.53 (237.78 to 495.71) | 13933 (8944 to 22277) | 325.57 (206.54 to 490.1) | 7.28 (4.52 to 11.01) | -0.58 (-1.05 to -0.11) |
| United Kingdom | 28073 (23289 to 33469) | 31.42 (26.06 to 37.15) | 40865 (33461 to 47996) | 31.2 (25.66 to 36.85) | 0.46 (0.35 to 0.58) | -0.33 (-0.62 to -0.04) |
| United Republic of Tanzania | 16133 (12541 to 20780) | 155.55 (122.49 to 199.32) | 29434 (24263 to 35489) | 121.94 (101.27 to 146.76) | 0.82 (0.41 to 1.39) | -1.17 (-1.39 to -0.95) |
| United States of America | 263040 (225164 to 299590) | 81.13 (69.52 to 92.23) | 713768 (605863 to 824688) | 127.03 (108.07 to 146.66) | 1.71 (1.48 to 1.99) | 1.83 (1.67 to 2) |
| United States Virgin Islands | 106 (85 to 132) | 125.28 (99.53 to 154.81) | 314 (251 to 387) | 179.38 (144.95 to 221.38) | 1.95 (1.35 to 2.67) | 1.74 (1.52 to 1.96) |
| Uruguay | 3382 (3051 to 3706) | 87.54 (79.11 to 95.99) | 5847 (5008 to 6786) | 101.43 (87.2 to 117.31) | 0.73 (0.51 to 0.98) | 0.86 (0.73 to 0.99) |
| Uzbekistan | 5920 (4505 to 7939) | 45.87 (34.81 to 62.31) | 14038 (10666 to 18143) | 61.8 (48.62 to 77.99) | 1.37 (0.82 to 1.89) | 0.74 (0.12 to 1.37) |
| Vanuatu | 153 (106 to 215) | 215.85 (151.5 to 301.04) | 630 (448 to 874) | 348.74 (252.4 to 480.52) | 3.12 (1.95 to 4.8) | 1.77 (1.64 to 1.89) |
| Venezuela (Bolivarian Republic of) | 10684 (8749 to 13180) | 104.82 (86.93 to 127.38) | 62281 (46607 to 82490) | 215.19 (162.01 to 284.34) | 4.83 (3.58 to 6.56) | 2.17 (1.7 to 2.65) |
| Viet Nam | 99917 (76513 to 129027) | 242.45 (187.5 to 312.11) | 194098 (147956 to 247934) | 215.95 (163.51 to 270.89) | 0.94 (0.37 to 1.59) | -0.4 (-0.83 to 0.02) |
| Yemen | 8394 (5890 to 11567) | 176.11 (125.14 to 241.04) | 21945 (16531 to 29696) | 168.64 (128.03 to 226.51) | 1.61 (0.99 to 2.5) | -0.2 (-0.29 to -0.11) |

| | | | | | | |
|---|---|---|---|---|---|---|
| Zambia | 6162 (4845 to 7775) | 214.37 (169.01 to 268.97) | 14256 (10707 to 18749) | 204.61 (155.99 to 264.06) | 1.31 (0.7 to 2.08) | -0.46 (-0.64 to -0.27) |
| Zimbabwe | 9584 (7013 to 14261) | 236 (174.29 to 349.05) | 23142 (16397 to 33904) | 322.58 (230.87 to 474.73) | 1.41 (0.82 to 2.12) | 1.43 (1.13 to 1.74) |

Table S8. The deaths count and ASMR of CKD due to hypertension in 1990 and 2019 for both sexes in 204 countries, and its temporal trends from 1990 to 2019.

| location | deaths count_1990 (95% UI) | ASMR_1990 (95% UI) | deaths count_2019 (95% UI) | ASMR_2019 (95% UI) | deaths count_change (*100%) (95% CI) | EAPC (95% CI) |
|---|---|---|---|---|---|---|
| Afghanistan | 1065 (731 to 1598) | 18.27 (12.63 to 28.81) | 1492 (1045 to 2240) | 16.39 (11.54 to 26.05) | 0.4 (0.01 to 0.85) | -0.3 (-0.35 to -0.26) |
| Albania | 54 (42 to 67) | 3.01 (2.36 to 3.76) | 91 (62 to 128) | 2.18 (1.51 to 3.03) | 0.7 (0.24 to 1.26) | -1.95 (-2.37 to -1.52) |
| Algeria | 1059 (742 to 1558) | 14.07 (10.19 to 19.77) | 2770 (1988 to 3739) | 11.63 (8.52 to 15.44) | 1.62 (0.94 to 2.5) | -0.46 (-0.62 to -0.29) |
| American Samoa | 3 (2 to 3) | 14.77 (11.56 to 18.11) | 9 (7 to 11) | 21.38 (17.14 to 26.06) | 2.37 (1.76 to 3.13) | 1.4 (1.26 to 1.54) |
| Andorra | 1 (1 to 2) | 3.88 (2.68 to 5.25) | 6 (4 to 8) | 3.48 (2.49 to 4.65) | 3.75 (2.27 to 5.7) | -0.31 (-0.49 to -0.12) |
| Angola | 285 (201 to 377) | 9.9 (7.05 to 12.72) | 715 (492 to 955) | 9.27 (6.2 to 12.16) | 1.51 (0.83 to 2.51) | -0.3 (-0.34 to -0.27) |
| Antigua and Barbuda | 4 (3 to 5) | 6.92 (5.57 to 8.38) | 10 (8 to 12) | 11.03 (8.61 to 13.61) | 1.48 (1.04 to 2.05) | 1.98 (1.74 to 2.23) |
| Argentina | 2357 (1909 to 2865) | 8.06 (6.58 to 9.66) | 5782 (4676 to 6933) | 10.32 (8.33 to 12.37) | 1.45 (1.1 to 1.85) | 0.78 (0.47 to 1.1) |
| Armenia | 9 (7 to 12) | 0.48 (0.35 to 0.65) | 61 (43 to 84) | 1.59 (1.12 to 2.19) | 5.61 (4.28 to 7.18) | 4.4 (4.13 to 4.67) |
| Australia | 355 (288 to 434) | 2.11 (1.67 to 2.61) | 1504 (1052 to 2066) | 2.95 (2.07 to 4.03) | 3.23 (2.3 to 4.23) | 1.36 (1.26 to 1.46) |
| Austria | 250 (191 to 312) | 2.06 (1.6 to 2.53) | 1273 (949 to 1617) | 5.41 (4.06 to 6.86) | 4.1 (3.26 to 5.11) | 5.71 (4.93 to 6.5) |
| Azerbaijan | 45 (34 to 61) | 0.97 (0.71 to 1.33) | 135 (96 to 186) | 2.27 (1.59 to 3.2) | 1.98 (1.19 to 2.92) | 3.93 (3.37 to 4.49) |

| | | | | | | |
|---|---|---|---|---|---|---|
| Bahamas | 8<br>(7 to 10) | 6.12<br>(4.93 to 7.45) | 27<br>(21 to 35) | 7.86<br>(6.08 to 9.96) | 2.24<br>(1.57 to 3.12) | 1.33<br>(1.13 to 1.54) |
| Bahrain | 13<br>(10 to 17) | 13.46<br>(10.37 to 17.22) | 55<br>(40 to 70) | 12.58<br>(9.56 to 15.96) | 3.12<br>(2.19 to 4.35) | 0.09<br>(-0.46 to 0.64) |
| Bangladesh | 1259<br>(892 to 1676) | 3.32<br>(2.31 to 4.39) | 3004<br>(2173 to 4028) | 2.84<br>(2.09 to 3.77) | 1.39<br>(0.77 to 2.28) | -0.36<br>(-0.77 to 0.05) |
| Barbados | 15<br>(12 to 18) | 4.89<br>(3.99 to 5.91) | 35<br>(27 to 43) | 7.03<br>(5.44 to 8.67) | 1.31<br>(0.88 to 1.82) | 1.29<br>(1.06 to 1.52) |
| Belarus | 77<br>(59 to 99) | 0.67<br>(0.51 to 0.84) | 139<br>(103 to 180) | 0.85<br>(0.63 to 1.09) | 0.8<br>(0.44 to 1.24) | 1<br>(0.68 to 1.33) |
| Belgium | 479<br>(372 to 598) | 3.09<br>(2.43 to 3.83) | 1177<br>(882 to 1498) | 3.76<br>(2.86 to 4.75) | 1.46<br>(1.05 to 1.94) | 0.4<br>(0.21 to 0.59) |
| Belize | 5<br>(4 to 7) | 6.01<br>(4.81 to 7.34) | 25<br>(20 to 31) | 10.27<br>(8.06 to 12.55) | 3.76<br>(2.97 to 4.82) | 2.13<br>(1.63 to 2.63) |
| Benin | 258<br>(209 to 313) | 14.68<br>(11.88 to 17.77) | 534<br>(403 to 695) | 13.43<br>(10.2 to 17.17) | 1.07<br>(0.57 to 1.69) | -0.21<br>(-0.29 to -0.14) |
| Bermuda | 3<br>(2 to 3) | 4.75<br>(3.87 to 5.72) | 6<br>(5 to 8) | 4.31<br>(3.29 to 5.45) | 1.26<br>(0.85 to 1.81) | 0.06<br>(-0.1 to 0.22) |
| Bhutan | 11<br>(7 to 17) | 5.67<br>(3.61 to 8.33) | 35<br>(24 to 48) | 7.2<br>(4.96 to 9.81) | 2.05<br>(1.07 to 3.41) | 0.95<br>(0.85 to 1.04) |
| Bolivia (Plurinational State of) | 269<br>(204 to 353) | 10.42<br>(7.94 to 13.5) | 1177<br>(851 to 1575) | 16.24<br>(11.88 to 21.57) | 3.38<br>(2.32 to 4.75) | 1.64<br>(1.53 to 1.74) |
| Bosnia and Herzegovina | 70<br>(54 to 89) | 2.14<br>(1.65 to 2.7) | 160<br>(111 to 223) | 2.82<br>(2 to 3.89) | 1.28<br>(0.73 to 2.02) | 1.03<br>(0.25 to 1.81) |
| Botswana | 54<br>(38 to 77) | 11.82<br>(8.44 to 16.74) | 162<br>(110 to 222) | 15.21<br>(10.44 to 20.77) | 1.99<br>(1.08 to 3.15) | 0.5<br>(0.21 to 0.79) |

| | | | | | | |
|---|---|---|---|---|---|---|
| Brazil | 3586<br>(2981 to 4211) | 5.08<br>(4.23 to 5.95) | 11665<br>(9627 to 13987) | 5.23<br>(4.28 to 6.25) | 2.25<br>(1.88 to 2.63) | 0.29<br>(0.22 to 0.36) |
| Brunei Darussalam | 7<br>(5 to 9) | 10.21<br>(7.89 to 12.9) | 19<br>(14 to 24) | 10.86<br>(8.34 to 13.74) | 1.81<br>(1.28 to 2.53) | 0.96<br>(0.72 to 1.2) |
| Bulgaria | 176<br>(135 to 226) | 1.69<br>(1.32 to 2.1) | 529<br>(395 to 699) | 3.58<br>(2.71 to 4.69) | 2<br>(1.28 to 2.81) | 2.89<br>(2.52 to 3.27) |
| Burkina Faso | 419<br>(329 to 515) | 12.64<br>(10.14 to 15.49) | 909<br>(710 to 1132) | 12.79<br>(10.21 to 15.77) | 1.17<br>(0.73 to 1.73) | -0.03<br>(-0.13 to 0.06) |
| Burundi | 204<br>(148 to 271) | 10.51<br>(7.69 to 13.91) | 297<br>(219 to 398) | 9.05<br>(6.72 to 11.88) | 0.46<br>(0.1 to 0.95) | -0.75<br>(-0.84 to -0.66) |
| Cabo Verde | 15<br>(13 to 19) | 6.37<br>(5.29 to 7.89) | 44<br>(36 to 52) | 10.45<br>(8.67 to 12.43) | 1.85<br>(1.09 to 2.44) | 0.85<br>(0.49 to 1.21) |
| Cambodia | 354<br>(273 to 447) | 8.51<br>(6.76 to 10.49) | 795<br>(619 to 993) | 7.79<br>(6.08 to 9.46) | 1.24<br>(0.64 to 1.94) | -0.39<br>(-0.48 to -0.3) |
| Cameroon | 684<br>(519 to 867) | 19.43<br>(14.7 to 24.54) | 1628<br>(1177 to 2174) | 17.13<br>(12.68 to 22.36) | 1.38<br>(0.77 to 2.24) | -0.51<br>(-0.71 to -0.31) |
| Canada | 1125<br>(934 to 1327) | 3.65<br>(3 to 4.3) | 3558<br>(2806 to 4251) | 4.41<br>(3.52 to 5.25) | 2.16<br>(1.74 to 2.63) | 0.44<br>(0.31 to 0.57) |
| Central African Republic | 100<br>(73 to 132) | 11.13<br>(8.28 to 14.33) | 178<br>(124 to 247) | 11.12<br>(7.84 to 14.85) | 0.77<br>(0.31 to 1.39) | 0.05<br>(-0.06 to 0.16) |
| Chad | 327<br>(236 to 463) | 13.32<br>(9.59 to 19.08) | 598<br>(434 to 798) | 13.01<br>(9.64 to 17.22) | 0.83<br>(0.43 to 1.33) | -0.06<br>(-0.15 to 0.02) |
| Chile | 457<br>(372 to 548) | 5.54<br>(4.54 to 6.61) | 1954<br>(1576 to 2348) | 8.22<br>(6.64 to 9.88) | 3.27<br>(2.71 to 3.88) | 1.84<br>(1.47 to 2.2) |
| China | 28727<br>(23475 to 34466) | 4.29<br>(3.55 to 5.07) | 70260<br>(56866 to 84481) | 4.09<br>(3.32 to 4.88) | 1.45<br>(0.99 to 2) | 0.21<br>(0.07 to 0.35) |

| | | | | | | |
|---|---|---|---|---|---|---|
| Colombia | 858<br>(700 to 1050) | 6.1<br>(4.95 to 7.39) | 2906<br>(2084 to 3881) | 5.22<br>(3.75 to 7.03) | 2.39<br>(1.59 to 3.22) | -0.35<br>(-0.51 to -0.18) |
| Comoros | 18<br>(12 to 25) | 10.23<br>(7.07 to 13.79) | 39<br>(30 to 49) | 9.53<br>(7.45 to 11.98) | 1.1<br>(0.56 to 2.1) | -0.38<br>(-0.48 to -0.28) |
| Congo | 106<br>(72 to 139) | 13.17<br>(8.86 to 17.06) | 219<br>(148 to 296) | 11.9<br>(7.99 to 15.93) | 1.07<br>(0.58 to 1.74) | -0.49<br>(-0.59 to -0.38) |
| Cook Islands | 1<br>(1 to 1) | 8.37<br>(6.67 to 10.26) | 3<br>(2 to 3) | 10.73<br>(8.51 to 13.25) | 1.77<br>(1.22 to 2.52) | 1.03<br>(0.95 to 1.11) |
| Costa Rica | 69<br>(56 to 86) | 4.37<br>(3.54 to 5.36) | 402<br>(292 to 537) | 7.75<br>(5.64 to 10.39) | 4.79<br>(3.54 to 6.31) | 1.97<br>(1.55 to 2.39) |
| Côte d'Ivoire | 502<br>(389 to 628) | 17.25<br>(13.7 to 21.27) | 1180<br>(892 to 1520) | 14.64<br>(11.47 to 18.27) | 1.35<br>(0.79 to 2.09) | -0.62<br>(-0.82 to -0.43) |
| Croatia | 114<br>(89 to 144) | 2<br>(1.56 to 2.51) | 292<br>(212 to 391) | 3<br>(2.19 to 3.96) | 1.56<br>(0.99 to 2.32) | 4.44<br>(3.12 to 5.77) |
| Cuba | 198<br>(163 to 238) | 2<br>(1.66 to 2.39) | 795<br>(605 to 1015) | 4<br>(3.03 to 5.14) | 3.01<br>(2.17 to 3.91) | 3.09<br>(2.79 to 3.39) |
| Cyprus | 52<br>(38 to 70) | 11<br>(8.08 to 14.56) | 108<br>(81 to 141) | 7.04<br>(5.28 to 9.1) | 1.06<br>(0.63 to 1.59) | -1.84<br>(-2.05 to -1.63) |
| Czechia | 277<br>(212 to 348) | 2.07<br>(1.61 to 2.58) | 401<br>(299 to 533) | 1.78<br>(1.32 to 2.36) | 0.45<br>(0.12 to 0.81) | -0.35<br>(-0.5 to -0.2) |
| Democratic People's Republic of Korea | 733<br>(552 to 962) | 5.42<br>(4.19 to 6.91) | 1505<br>(1151 to 1913) | 5.05<br>(3.89 to 6.4) | 1.05<br>(0.6 to 1.67) | -0.14<br>(-0.3 to 0.02) |
| Democratic Republic of the Congo | 1209<br>(946 to 1529) | 11.12<br>(8.9 to 13.6) | 2421<br>(1777 to 3212) | 9.18<br>(6.78 to 12.02) | 1<br>(0.47 to 1.64) | -0.77<br>(-0.82 to -0.73) |
| Denmark | 110<br>(84 to 140) | 1.26<br>(0.97 to 1.6) | 381<br>(284 to 480) | 2.85<br>(2.15 to 3.58) | 2.47<br>(1.89 to 3.03) | 2.35<br>(2.05 to 2.65) |

| | | | | | | |
|---|---|---|---|---|---|---|
| Djibouti | 8<br>(5 to 11) | 8.48<br>(5.99 to 11.51) | 39<br>(28 to 55) | 10.2<br>(7.51 to 13.61) | 4.1<br>(2.65 to 6.06) | 0.57<br>(0.48 to 0.66) |
| Dominica | 7<br>(6 to 8) | 9.37<br>(7.5 to 11.4) | 12<br>(9 to 15) | 12.81<br>(9.83 to 16.22) | 0.68<br>(0.35 to 1.12) | 1.48<br>(1.34 to 1.62) |
| Dominican Republic | 123<br>(99 to 152) | 4<br>(3.2 to 4.97) | 584<br>(417 to 804) | 6.78<br>(4.89 to 9.26) | 3.74<br>(2.51 to 5.37) | 2.98<br>(2.62 to 3.34) |
| Ecuador | 260<br>(213 to 314) | 5.86<br>(4.82 to 7.03) | 1974<br>(1485 to 2550) | 15.4<br>(11.71 to 19.74) | 6.59<br>(5.04 to 8.35) | 3.85<br>(3.15 to 4.56) |
| Egypt | 2964<br>(1962 to 3938) | 13.88<br>(9.24 to 18.28) | 7014<br>(4061 to 10224) | 15.47<br>(9.38 to 22.05) | 1.37<br>(0.67 to 2.17) | 0.5<br>(0.38 to 0.63) |
| El Salvador | 140<br>(113 to 171) | 4.97<br>(4.02 to 6.07) | 1045<br>(751 to 1428) | 16.81<br>(11.98 to 23.2) | 6.44<br>(4.54 to 8.58) | 4.48<br>(3.72 to 5.24) |
| Equatorial Guinea | 16<br>(11 to 22) | 10.36<br>(7.43 to 13.54) | 43<br>(30 to 60) | 12.29<br>(8.62 to 16.69) | 1.63<br>(0.77 to 2.84) | 0.75<br>(0.61 to 0.9) |
| Eritrea | 59<br>(37 to 86) | 8.26<br>(4.96 to 12.55) | 178<br>(120 to 248) | 9.73<br>(6.65 to 13.57) | 2<br>(1.14 to 3.28) | 0.42<br>(0.24 to 0.59) |
| Estonia | 15<br>(12 to 19) | 0.82<br>(0.65 to 1.03) | 117<br>(87 to 156) | 3.63<br>(2.72 to 4.78) | 6.69<br>(5.02 to 8.81) | 6.23<br>(5.75 to 6.7) |
| Eswatini | 37<br>(28 to 47) | 15.3<br>(11.67 to 19.24) | 92<br>(65 to 126) | 19.96<br>(14.42 to 26.23) | 1.52<br>(0.81 to 2.51) | 1.06<br>(0.5 to 1.63) |
| Ethiopia | 2064<br>(1587 to 2619) | 13.34<br>(10.42 to 16.55) | 3121<br>(2521 to 3818) | 9.6<br>(7.74 to 11.7) | 0.51<br>(0.18 to 0.94) | -1.24<br>(-1.33 to -1.16) |
| Fiji | 33<br>(25 to 43) | 11.68<br>(9.05 to 15.02) | 92<br>(68 to 119) | 16.3<br>(12.54 to 20.6) | 1.8<br>(1.04 to 2.81) | 0.64<br>(0.26 to 1.02) |
| Finland | 51<br>(38 to 67) | 0.74<br>(0.56 to 0.97) | 210<br>(147 to 283) | 1.32<br>(0.93 to 1.76) | 3.14<br>(2.31 to 4) | -0.16<br>(-1.68 to 1.37) |

| | | | | | | |
|---|---|---|---|---|---|---|
| France | 2767<br>(2138 to 3409) | 3.08<br>(2.4 to 3.76) | 5340<br>(4060 to 6703) | 2.62<br>(2.01 to 3.28) | 0.93<br>(0.59 to 1.3) | -0.11<br>(-0.31 to 0.09) |
| Gabon | 58<br>(40 to 76) | 12.64<br>(8.63 to 16.39) | 121<br>(74 to 164) | 15.27<br>(9.1 to 20.57) | 1.09<br>(0.47 to 1.82) | 0.59<br>(0.4 to 0.79) |
| Gambia | 38<br>(28 to 49) | 13.88<br>(10.69 to 17.57) | 118<br>(91 to 151) | 14.44<br>(11.53 to 18.26) | 2.11<br>(1.23 to 3.26) | 0.09<br>(0 to 0.19) |
| Georgia | 38<br>(28 to 50) | 0.73<br>(0.55 to 0.97) | 93<br>(66 to 127) | 1.42<br>(1.01 to 1.91) | 1.47<br>(0.91 to 2.1) | 1.99<br>(1.24 to 2.76) |
| Germany | 2485<br>(1878 to 3201) | 1.85<br>(1.42 to 2.36) | 11441<br>(8207 to 14944) | 4.65<br>(3.39 to 6.05) | 3.6<br>(2.74 to 4.64) | 4.94<br>(4.44 to 5.45) |
| Ghana | 529<br>(378 to 705) | 11.02<br>(8.15 to 14.73) | 1529<br>(1052 to 1975) | 12.18<br>(8.54 to 15.65) | 1.89<br>(1.03 to 2.86) | 0.45<br>(0.38 to 0.53) |
| Greece | 879<br>(650 to 1117) | 6.39<br>(4.77 to 8.05) | 1552<br>(1156 to 1999) | 4.73<br>(3.58 to 6.05) | 0.76<br>(0.48 to 1.07) | -1.36<br>(-1.87 to -0.84) |
| Greenland | 1<br>(1 to 1) | 5.3<br>(4.19 to 6.45) | 3<br>(2 to 4) | 6.09<br>(4.74 to 7.49) | 1.74<br>(1.15 to 2.39) | 0.59<br>(0.48 to 0.71) |
| Grenada | 7<br>(6 to 8) | 8.84<br>(7.08 to 10.79) | 12<br>(10 to 15) | 12.34<br>(9.95 to 15.06) | 0.73<br>(0.43 to 1.06) | 1.52<br>(1.34 to 1.71) |
| Guam | 5<br>(4 to 6) | 9.48<br>(7.82 to 11.21) | 20<br>(16 to 25) | 10.56<br>(8.29 to 13.18) | 3.08<br>(2.24 to 4.08) | 0.59<br>(0.27 to 0.91) |
| Guatemala | 233<br>(186 to 292) | 8.81<br>(7.16 to 10.89) | 1433<br>(1053 to 1913) | 15.22<br>(11.37 to 19.89) | 5.14<br>(3.68 to 6.87) | 2.45<br>(2.14 to 2.75) |
| Guinea | 435<br>(329 to 581) | 15.45<br>(11.73 to 20.61) | 666<br>(497 to 848) | 13.9<br>(10.52 to 17.55) | 0.53<br>(0.17 to 1.03) | 0.04<br>(-0.08 to 0.16) |
| Guinea-Bissau | 68<br>(52 to 87) | 20.04<br>(15.59 to 25.3) | 94<br>(71 to 123) | 16.1<br>(12.32 to 20.25) | 0.39<br>(0.04 to 0.86) | -0.74<br>(-0.77 to -0.71) |

| | | | | | | |
|---|---|---|---|---|---|---|
| Guyana | 24<br>(18 to 29) | 7.33<br>(5.75 to 9.01) | 62<br>(45 to 81) | 11.61<br>(8.67 to 15.05) | 1.63<br>(1.01 to 2.44) | 2.27<br>(2.04 to 2.49) |
| Haiti | 217<br>(146 to 343) | 8.13<br>(5.46 to 13.14) | 494<br>(325 to 785) | 8.72<br>(5.77 to 13.88) | 1.28<br>(0.67 to 2.08) | 0.55<br>(0.42 to 0.69) |
| Honduras | 120<br>(85 to 187) | 6.75<br>(4.71 to 10.94) | 776<br>(571 to 1048) | 15.02<br>(11.13 to 20.4) | 5.47<br>(3.81 to 7.37) | 3.27<br>(2.95 to 3.58) |
| Hungary | 206<br>(161 to 257) | 1.54<br>(1.23 to 1.89) | 583<br>(432 to 755) | 2.76<br>(2.06 to 3.56) | 1.83<br>(1.26 to 2.52) | 3.29<br>(2.68 to 3.9) |
| Iceland | 5<br>(4 to 6) | 1.63<br>(1.29 to 2) | 14<br>(11 to 18) | 2.06<br>(1.54 to 2.57) | 1.87<br>(1.41 to 2.37) | 1.38<br>(1.05 to 1.71) |
| India | 20274<br>(15874 to 25642) | 6.06<br>(4.73 to 7.61) | 50909<br>(39097 to 64888) | 5.2<br>(4.02 to 6.51) | 1.51<br>(1.01 to 2.17) | -0.87<br>(-1.1 to -0.65) |
| Indonesia | 6966<br>(5713 to 8439) | 7.69<br>(6.39 to 9.16) | 14999<br>(12108 to 18123) | 8<br>(6.56 to 9.47) | 1.15<br>(0.73 to 1.59) | 0.15<br>(0.01 to 0.3) |
| Iran (Islamic Republic of) | 1458<br>(1203 to 1777) | 8.23<br>(6.79 to 10.3) | 4322<br>(3587 to 5063) | 7.08<br>(5.85 to 8.25) | 1.96<br>(1.46 to 2.42) | -0.7<br>(-0.81 to -0.59) |
| Iraq | 1006<br>(757 to 1401) | 15.2<br>(11.31 to 21.54) | 2443<br>(1810 to 3250) | 14.35<br>(10.63 to 19.39) | 1.43<br>(0.86 to 2.04) | -0.24<br>(-0.31 to -0.16) |
| Ireland | 131<br>(99 to 167) | 3.61<br>(2.76 to 4.55) | 247<br>(184 to 321) | 3.11<br>(2.33 to 4.03) | 0.88<br>(0.54 to 1.24) | -0.44<br>(-0.53 to -0.36) |
| Israel | 337<br>(263 to 413) | 8.09<br>(6.32 to 9.82) | 1039<br>(784 to 1294) | 7.85<br>(5.98 to 9.74) | 2.08<br>(1.63 to 2.59) | 0.13<br>(-0.11 to 0.38) |
| Italy | 2883<br>(2332 to 3458) | 3.37<br>(2.74 to 4) | 6504<br>(5105 to 7929) | 3.23<br>(2.54 to 3.91) | 1.26<br>(0.9 to 1.58) | 0.27<br>(0.11 to 0.43) |
| Jamaica | 120<br>(98 to 144) | 6.56<br>(5.33 to 7.84) | 245<br>(187 to 313) | 7.63<br>(5.76 to 9.82) | 1.04<br>(0.62 to 1.54) | -0.08<br>(-0.55 to 0.4) |

| | | | | | | |
|---|---|---|---|---|---|---|
| Japan | 6208<br>(4901 to 7504) | 4.2<br>(3.34 to 5.05) | 16599<br>(11772 to 20812) | 3.04<br>(2.24 to 3.75) | 1.67<br>(1.19 to 2.11) | -1.85<br>(-2.25 to -1.46) |
| Jordan | 127<br>(98 to 160) | 13.89<br>(10.7 to 17.41) | 576<br>(451 to 713) | 13.01<br>(10.16 to 16.07) | 3.54<br>(2.53 to 4.75) | -0.09<br>(-0.21 to 0.03) |
| Kazakhstan | 83<br>(64 to 108) | 0.74<br>(0.57 to 0.96) | 210<br>(152 to 283) | 1.51<br>(1.1 to 2.04) | 1.51<br>(1.01 to 2.08) | 2.35<br>(2.02 to 2.68) |
| Kenya | 456<br>(355 to 585) | 7.15<br>(5.56 to 9.12) | 1375<br>(1084 to 1723) | 8.67<br>(6.9 to 10.71) | 2.01<br>(1.5 to 2.56) | 0.71<br>(0.67 to 0.76) |
| Kiribati | 4<br>(3 to 6) | 14<br>(10.71 to 17.56) | 10<br>(7 to 14) | 18.66<br>(13.23 to 24.49) | 1.27<br>(0.63 to 2.09) | 0.74<br>(0.33 to 1.16) |
| Kuwait | 41<br>(33 to 50) | 9.67<br>(7.64 to 11.7) | 110<br>(86 to 139) | 6.01<br>(4.63 to 7.65) | 1.7<br>(1.23 to 2.29) | -2.04<br>(-2.74 to -1.33) |
| Kyrgyzstan | 32<br>(24 to 42) | 1.04<br>(0.79 to 1.37) | 43<br>(31 to 56) | 1.04<br>(0.77 to 1.41) | 0.35<br>(0.1 to 0.63) | -0.35<br>(-0.81 to 0.11) |
| Lao People's Democratic Republic | 359<br>(263 to 492) | 19.25<br>(14.21 to 26.13) | 608<br>(464 to 801) | 16.29<br>(12.6 to 20.87) | 0.7<br>(0.28 to 1.35) | -0.76<br>(-0.82 to -0.69) |
| Latvia | 19<br>(14 to 25) | 0.58<br>(0.43 to 0.75) | 65<br>(46 to 87) | 1.41<br>(1.01 to 1.88) | 2.38<br>(1.68 to 3.23) | 4.31<br>(3.01 to 5.63) |
| Lebanon | 203<br>(157 to 260) | 11.43<br>(8.82 to 14.59) | 427<br>(301 to 569) | 8.51<br>(6 to 11.4) | 1.1<br>(0.47 to 1.8) | -0.98<br>(-1.08 to -0.87) |
| Lesotho | 90<br>(69 to 116) | 10.62<br>(8.36 to 13.6) | 205<br>(144 to 275) | 19.9<br>(14.43 to 25.86) | 1.28<br>(0.62 to 2.05) | 2.75<br>(2.46 to 3.04) |
| Liberia | 157<br>(124 to 203) | 17.2<br>(13.65 to 21.72) | 228<br>(165 to 318) | 13.89<br>(10.17 to 19.14) | 0.45<br>(0.07 to 0.94) | -0.53<br>(-0.76 to -0.31) |
| Libya | 164<br>(118 to 223) | 10.35<br>(7.44 to 14.41) | 450<br>(295 to 632) | 10.56<br>(6.92 to 14.89) | 1.75<br>(1 to 2.67) | 0.36<br>(0.18 to 0.53) |

| | | | | | | |
|---|---|---|---|---|---|---|
| Lithuania | 32<br>(25 to 41) | 0.75<br>(0.58 to 0.94) | 74<br>(56 to 99) | 1.14<br>(0.87 to 1.5) | 1.31<br>(0.82 to 1.88) | 1.13<br>(0.71 to 1.56) |
| Luxembourg | 16<br>(12 to 21) | 3.3<br>(2.47 to 4.2) | 42<br>(30 to 54) | 3.44<br>(2.51 to 4.42) | 1.53<br>(1.03 to 2.08) | 0.35<br>(0.13 to 0.57) |
| Madagascar | 323<br>(231 to 430) | 7.96<br>(5.73 to 10.6) | 550<br>(390 to 753) | 7.34<br>(5.29 to 9.84) | 0.7<br>(0.23 to 1.29) | -0.38<br>(-0.47 to -0.29) |
| Malawi | 268<br>(210 to 336) | 9.59<br>(7.57 to 11.81) | 523<br>(404 to 655) | 9.38<br>(7.28 to 11.65) | 0.95<br>(0.54 to 1.5) | -0.14<br>(-0.27 to -0.02) |
| Malaysia | 800<br>(663 to 934) | 9.82<br>(8.15 to 11.46) | 2376<br>(1822 to 3034) | 10.38<br>(8.1 to 13.1) | 1.97<br>(1.3 to 2.7) | -0.39<br>(-0.66 to -0.13) |
| Maldives | 15<br>(12 to 19) | 23.23<br>(18.81 to 29.04) | 36<br>(28 to 44) | 14.47<br>(11.55 to 17.72) | 1.32<br>(0.7 to 1.97) | -2.01<br>(-2.24 to -1.79) |
| Mali | 498<br>(382 to 655) | 15.2<br>(11.77 to 20.09) | 895<br>(688 to 1160) | 12.87<br>(10.01 to 16.31) | 0.8<br>(0.38 to 1.33) | -0.44<br>(-0.58 to -0.29) |
| Malta | 16<br>(12 to 20) | 4.29<br>(3.23 to 5.46) | 35<br>(26 to 46) | 3.35<br>(2.49 to 4.34) | 1.27<br>(0.84 to 1.75) | -0.88<br>(-1.08 to -0.69) |
| Marshall Islands | 2<br>(1 to 3) | 13.64<br>(10.12 to 18.68) | 5<br>(3 to 7) | 18.33<br>(13.12 to 25.41) | 1.56<br>(0.94 to 2.31) | 1.05<br>(0.8 to 1.3) |
| Mauritania | 167<br>(132 to 204) | 19.66<br>(15.58 to 23.56) | 230<br>(172 to 299) | 13.24<br>(9.93 to 16.91) | 0.38<br>(0.06 to 0.72) | -1.4<br>(-1.46 to -1.33) |
| Mauritius | 114<br>(96 to 134) | 17.36<br>(14.7 to 20.21) | 469<br>(363 to 590) | 28.1<br>(21.91 to 35.16) | 3.11<br>(2.27 to 4.14) | 1.94<br>(1.6 to 2.28) |
| Mexico | 2826<br>(2361 to 3314) | 8.31<br>(6.96 to 9.7) | 17888<br>(14225 to 22190) | 16.31<br>(12.98 to 20.17) | 5.33<br>(4.38 to 6.31) | 2.69<br>(2.31 to 3.07) |
| Micronesia (Federated States of) | 7<br>(5 to 9) | 16.74<br>(12.77 to 22.52) | 15<br>(10 to 20) | 26.31<br>(18.93 to 34.3) | 1.24<br>(0.55 to 2.14) | 1.5<br>(1.11 to 1.89) |

| | | | | | | |
|---|---|---|---|---|---|---|
| Monaco | 1<br>(1 to 2) | 1.35<br>(0.98 to 1.79) | 3<br>(2 to 4) | 2.14<br>(1.6 to 2.77) | 1.4<br>(0.91 to 2.07) | 2.02<br>(1.59 to 2.45) |
| Mongolia | 37<br>(27 to 50) | 4.43<br>(3.26 to 5.94) | 36<br>(25 to 50) | 2.35<br>(1.64 to 3.23) | -0.04<br>(-0.28 to 0.27) | -3.25<br>(-3.7 to -2.8) |
| Montenegro | 20<br>(15 to 25) | 3.53<br>(2.75 to 4.41) | 38<br>(28 to 49) | 4.11<br>(3.09 to 5.3) | 0.89<br>(0.45 to 1.37) | 1.69<br>(1.34 to 2.04) |
| Morocco | 1051<br>(800 to 1504) | 9.87<br>(7.37 to 14.65) | 2984<br>(2222 to 3958) | 12.27<br>(9.2 to 16.5) | 1.84<br>(1.11 to 2.64) | 0.76<br>(0.57 to 0.95) |
| Mozambique | 342<br>(258 to 457) | 8.13<br>(6.24 to 10.7) | 730<br>(549 to 957) | 9.18<br>(6.93 to 11.89) | 1.14<br>(0.49 to 1.97) | 0.48<br>(0.33 to 0.64) |
| Myanmar | 2389<br>(1743 to 3158) | 11.24<br>(8.35 to 14.61) | 4124<br>(3295 to 5180) | 10.25<br>(8.34 to 12.59) | 0.73<br>(0.27 to 1.38) | -0.37<br>(-0.47 to -0.26) |
| Namibia | 74<br>(53 to 105) | 12.38<br>(8.95 to 17.39) | 145<br>(102 to 206) | 12.15<br>(8.56 to 17.25) | 0.96<br>(0.42 to 1.56) | -0.34<br>(-0.66 to -0.02) |
| Nauru | 0<br>(0 to 1) | 16.63<br>(12.52 to 21.3) | 1<br>(0 to 1) | 22.78<br>(16.21 to 29.04) | 0.41<br>(0.07 to 0.84) | 0.95<br>(0.65 to 1.26) |
| Nepal | 363<br>(251 to 519) | 4.62<br>(3.18 to 6.67) | 1203<br>(820 to 1661) | 6.51<br>(4.44 to 8.95) | 2.31<br>(1.31 to 3.46) | 1.29<br>(1.02 to 1.56) |
| Netherlands | 642<br>(496 to 798) | 3.23<br>(2.52 to 4.01) | 1537<br>(1171 to 1916) | 3.9<br>(2.99 to 4.85) | 1.39<br>(1.04 to 1.77) | 1.56<br>(1.31 to 1.81) |
| New Zealand | 78<br>(61 to 98) | 2.14<br>(1.66 to 2.67) | 264<br>(201 to 331) | 2.96<br>(2.25 to 3.69) | 2.38<br>(1.9 to 2.98) | 1.2<br>(0.99 to 1.42) |
| Nicaragua | 131<br>(104 to 163) | 9.74<br>(7.76 to 12.04) | 847<br>(632 to 1092) | 22.32<br>(17.04 to 28.42) | 5.44<br>(4.14 to 6.9) | 3.24<br>(2.82 to 3.66) |
| Niger | 308<br>(230 to 409) | 14.49<br>(10.95 to 19.22) | 708<br>(527 to 947) | 12.08<br>(9.1 to 15.67) | 1.3<br>(0.78 to 2.01) | -0.6<br>(-0.65 to -0.55) |

| | | | | | | |
|---|---|---|---|---|---|---|
| Nigeria | 3961<br>(3090 to 5183) | 11.4<br>(8.99 to 14.63) | 6543<br>(5033 to 8318) | 9.83<br>(7.66 to 12.28) | 0.65<br>(0.24 to 1.13) | -0.36<br>(-0.48 to -0.23) |
| Niue | 0<br>(0 to 0) | 12.37<br>(9.23 to 15.76) | 0<br>(0 to 0) | 16.66<br>(11.88 to 21.73) | 0.23<br>(-0.06 to 0.56) | 0.93<br>(0.67 to 1.19) |
| North Macedonia | 56<br>(44 to 70) | 3.45<br>(2.75 to 4.28) | 109<br>(79 to 145) | 3.98<br>(2.97 to 5.2) | 0.94<br>(0.48 to 1.53) | 0.74<br>(0.35 to 1.12) |
| Northern Mariana Islands | 2<br>(2 to 3) | 16.53<br>(13.22 to 20.45) | 8<br>(6 to 10) | 20.96<br>(16.71 to 25.17) | 2.94<br>(2.07 to 3.88) | 1.02<br>(0.91 to 1.12) |
| Norway | 121<br>(99 to 145) | 1.58<br>(1.3 to 1.89) | 298<br>(236 to 361) | 2.48<br>(1.97 to 3) | 1.47<br>(1.18 to 1.77) | 2.23<br>(1.75 to 2.71) |
| Oman | 25<br>(18 to 35) | 5.93<br>(4.3 to 7.99) | 62<br>(48 to 78) | 7.17<br>(5.58 to 8.76) | 1.46<br>(0.77 to 2.38) | 1.11<br>(0.9 to 1.33) |
| Pakistan | 3469<br>(2389 to 4997) | 6.85<br>(4.76 to 9.97) | 9090<br>(6385 to 12273) | 9.91<br>(7.07 to 13.05) | 1.62<br>(0.91 to 2.48) | 1.32<br>(1.07 to 1.58) |
| Palau | 2<br>(1 to 2) | 18.83<br>(14.5 to 24.14) | 4<br>(3 to 5) | 24.04<br>(18.56 to 30.29) | 1.57<br>(0.9 to 2.41) | 0.78<br>(0.57 to 0.99) |
| Palestine | 121<br>(93 to 154) | 16.4<br>(12.7 to 20.78) | 203<br>(157 to 253) | 11.74<br>(9.16 to 14.47) | 0.68<br>(0.28 to 1.17) | -0.87<br>(-1.12 to -0.62) |
| Panama | 56<br>(45 to 69) | 4.02<br>(3.24 to 4.97) | 324<br>(239 to 432) | 7.6<br>(5.58 to 10.19) | 4.8<br>(3.53 to 6.45) | 2.33<br>(1.95 to 2.71) |
| Papua New Guinea | 62<br>(47 to 83) | 3.87<br>(2.97 to 5.05) | 190<br>(135 to 262) | 4.65<br>(3.43 to 6.12) | 2.04<br>(1.22 to 3.11) | 0.58<br>(0.44 to 0.72) |
| Paraguay | 85<br>(68 to 105) | 4.27<br>(3.37 to 5.23) | 491<br>(359 to 650) | 9.4<br>(6.87 to 12.39) | 4.78<br>(3.46 to 6.54) | 3.43<br>(3.2 to 3.67) |
| Peru | 718<br>(572 to 900) | 6.84<br>(5.42 to 8.6) | 2463<br>(1761 to 3306) | 7.53<br>(5.38 to 10.12) | 2.43<br>(1.5 to 3.56) | 0.46<br>(0.19 to 0.73) |

| | | | | | | |
|---|---|---|---|---|---|---|
| Philippines | 3620<br>(3007 to 4280) | 15.2<br>(12.81 to 17.79) | 12083<br>(9534 to 15312) | 17.28<br>(13.87 to 21.57) | 2.34<br>(1.67 to 3.17) | 0.79<br>(0.53 to 1.04) |
| Poland | 1125<br>(916 to 1339) | 2.72<br>(2.24 to 3.22) | 1437<br>(1103 to 1817) | 1.92<br>(1.47 to 2.43) | 0.28<br>(0.04 to 0.56) | -0.3<br>(-0.96 to 0.37) |
| Portugal | 465<br>(353 to 591) | 3.85<br>(2.96 to 4.82) | 1462<br>(1081 to 1875) | 4.62<br>(3.46 to 5.88) | 2.14<br>(1.55 to 2.76) | 0.86<br>(0.4 to 1.32) |
| Puerto Rico | 275<br>(223 to 327) | 8.05<br>(6.55 to 9.52) | 633<br>(462 to 820) | 7.53<br>(5.44 to 9.82) | 1.3<br>(0.82 to 1.91) | 0.42<br>(0.07 to 0.77) |
| Qatar | 8<br>(6 to 13) | 17.06<br>(12.34 to 30.05) | 40<br>(29 to 53) | 17.51<br>(13.24 to 22.45) | 3.79<br>(2.14 to 5.85) | 0.4<br>(0.15 to 0.66) |
| Republic of Korea | 745<br>(627 to 902) | 3.47<br>(2.88 to 4.15) | 2593<br>(2102 to 3160) | 3.16<br>(2.54 to 3.86) | 2.48<br>(1.86 to 3.23) | 0.15<br>(-0.02 to 0.32) |
| Republic of Moldova | 23<br>(18 to 29) | 0.77<br>(0.6 to 0.97) | 59<br>(46 to 75) | 1.04<br>(0.8 to 1.31) | 1.57<br>(1.08 to 2.1) | 1.12<br>(0.85 to 1.39) |
| Romania | 404<br>(341 to 468) | 1.7<br>(1.45 to 1.98) | 796<br>(567 to 1067) | 1.98<br>(1.41 to 2.64) | 0.97<br>(0.5 to 1.55) | -0.73<br>(-1.47 to 0.03) |
| Russian Federation | 1657<br>(1354 to 2007) | 1.05<br>(0.86 to 1.26) | 2860<br>(2210 to 3586) | 1.23<br>(0.95 to 1.53) | 0.73<br>(0.44 to 1.04) | -0.05<br>(-0.78 to 0.7) |
| Rwanda | 246<br>(189 to 315) | 10.93<br>(8.48 to 13.79) | 380<br>(289 to 488) | 8.92<br>(6.71 to 11.34) | 0.55<br>(0.17 to 1.03) | -1.32<br>(-1.56 to -1.08) |
| Saint Kitts and Nevis | 5<br>(4 to 6) | 12.75<br>(10.27 to 15.28) | 8<br>(6 to 10) | 14.39<br>(11.36 to 18.01) | 0.72<br>(0.38 to 1.14) | 1.08<br>(0.88 to 1.27) |
| Saint Lucia | 6<br>(5 to 8) | 8.32<br>(6.7 to 10.06) | 19<br>(14 to 24) | 9.31<br>(7.15 to 11.7) | 1.9<br>(1.33 to 2.53) | 0.41<br>(0.16 to 0.65) |
| Saint Vincent and the Grenadines | 4<br>(4 to 5) | 6.58<br>(5.33 to 7.91) | 12<br>(9 to 14) | 9.5<br>(7.58 to 11.55) | 1.65<br>(1.21 to 2.21) | 1.68<br>(1.41 to 1.95) |

| | | | | | | |
|---|---|---|---|---|---|---|
| Samoa | 10<br>(7 to 13) | 13.27<br>(10.07 to 17.5) | 20<br>(16 to 26) | 15.66<br>(12.14 to 20.17) | 1.02<br>(0.55 to 1.73) | 0.48<br>(0.27 to 0.7) |
| San Marino | 0<br>(0 to 1) | 1.36<br>(0.98 to 1.79) | 2<br>(1 to 2) | 1.75<br>(1.14 to 2.53) | 2.78<br>(1.63 to 4.19) | 1.61<br>(1.34 to 1.89) |
| Sao Tome and Principe | 10<br>(8 to 12) | 18.87<br>(15.26 to 22.36) | 19<br>(15 to 24) | 22.5<br>(17.45 to 27.36) | 0.92<br>(0.52 to 1.4) | 0.56<br>(0.46 to 0.65) |
| Saudi Arabia | 764<br>(566 to 1017) | 16.88<br>(12.69 to 21.86) | 2105<br>(1567 to 2701) | 19.29<br>(14.72 to 24.09) | 1.76<br>(0.89 to 2.87) | 0.73<br>(0.52 to 0.95) |
| Senegal | 459<br>(347 to 610) | 17.41<br>(13.44 to 23.2) | 901<br>(685 to 1188) | 14.56<br>(11.32 to 18.84) | 0.96<br>(0.53 to 1.53) | -0.57<br>(-0.67 to -0.46) |
| Serbia | 384<br>(295 to 497) | 3.95<br>(3.03 to 5.12) | 781<br>(541 to 1074) | 5.15<br>(3.74 to 6.94) | 1.04<br>(0.53 to 1.66) | 2.02<br>(1.64 to 2.41) |
| Seychelles | 7<br>(6 to 8) | 12.22<br>(10.25 to 14.44) | 17<br>(14 to 21) | 18.2<br>(15 to 21.7) | 1.56<br>(1.18 to 1.99) | 0.92<br>(0.53 to 1.31) |
| Sierra Leone | 230<br>(178 to 293) | 13.87<br>(10.86 to 17.28) | 355<br>(269 to 465) | 11.88<br>(9.24 to 15.22) | 0.54<br>(0.18 to 1.02) | -0.36<br>(-0.42 to -0.29) |
| Singapore | 80<br>(64 to 99) | 4.46<br>(3.55 to 5.49) | 243<br>(188 to 302) | 3.44<br>(2.64 to 4.28) | 2.05<br>(1.5 to 2.59) | 0.69<br>(-0.32 to 1.71) |
| Slovakia | 173<br>(132 to 221) | 3.01<br>(2.33 to 3.79) | 221<br>(158 to 298) | 2.43<br>(1.74 to 3.26) | 0.27<br>(-0.03 to 0.63) | -1.04<br>(-1.81 to -0.27) |
| Slovenia | 22<br>(16 to 29) | 0.96<br>(0.72 to 1.25) | 80<br>(56 to 110) | 1.46<br>(1.02 to 2) | 2.63<br>(1.43 to 4.18) | 2.31<br>(1.95 to 2.67) |
| Solomon Islands | 17<br>(12 to 24) | 12.83<br>(9.39 to 18.26) | 22<br>(17 to 29) | 7.69<br>(6.13 to 9.57) | 0.31<br>(-0.09 to 0.83) | -1.96<br>(-2.36 to -1.55) |
| Somalia | 194<br>(134 to 262) | 10.62<br>(7.49 to 14.04) | 476<br>(338 to 665) | 10.21<br>(7.42 to 14) | 1.46<br>(0.83 to 2.37) | -0.02<br>(-0.08 to 0.04) |

| | | | | | | |
|---|---|---|---|---|---|---|
| South Africa | 1465<br>(1236 to 1729) | 7.85<br>(6.63 to 9.22) | 4392<br>(3703 to 5195) | 11.65<br>(9.79 to 13.67) | 2<br>(1.63 to 2.38) | 0.68<br>(0.03 to 1.33) |
| South Sudan | 185<br>(126 to 257) | 9.76<br>(6.69 to 13.39) | 275<br>(191 to 389) | 9.86<br>(6.94 to 13.59) | 0.49<br>(0.09 to 1.03) | 0.02<br>(-0.02 to 0.05) |
| Spain | 2242<br>(1714 to 2860) | 4.43<br>(3.39 to 5.59) | 5215<br>(3737 to 6892) | 3.74<br>(2.72 to 4.92) | 1.33<br>(0.89 to 1.79) | -0.91<br>(-1.17 to -0.64) |
| Sri Lanka | 923<br>(755 to 1103) | 10.26<br>(8.47 to 12.18) | 2171<br>(1554 to 2920) | 9.59<br>(6.93 to 12.81) | 1.35<br>(0.71 to 2.2) | -0.23<br>(-0.63 to 0.18) |
| Sudan | 793<br>(573 to 1170) | 10.46<br>(7.58 to 15.93) | 1568<br>(1038 to 2517) | 10.6<br>(7.11 to 17.32) | 0.98<br>(0.31 to 1.97) | 0.36<br>(0.21 to 0.5) |
| Suriname | 17<br>(14 to 21) | 7.25<br>(5.88 to 8.81) | 65<br>(50 to 82) | 11.68<br>(8.94 to 14.77) | 2.71<br>(1.97 to 3.54) | 1.8<br>(1.49 to 2.1) |
| Sweden | 190<br>(150 to 234) | 1.12<br>(0.9 to 1.37) | 660<br>(504 to 817) | 2.37<br>(1.82 to 2.92) | 2.47<br>(1.96 to 3.08) | 2.6<br>(2.46 to 2.74) |
| Switzerland | 205<br>(153 to 259) | 1.8<br>(1.37 to 2.26) | 873<br>(631 to 1133) | 3.57<br>(2.61 to 4.6) | 3.26<br>(2.48 to 4.08) | 3.85<br>(3.41 to 4.29) |
| Syrian Arab Republic | 554<br>(422 to 733) | 12.83<br>(9.82 to 17.19) | 887<br>(648 to 1183) | 10.27<br>(7.61 to 13.26) | 0.6<br>(0.12 to 1.22) | -1.42<br>(-1.71 to -1.13) |
| Taiwan (Province of China) | 1154<br>(979 to 1352) | 9.66<br>(8.25 to 11.26) | 3251<br>(2492 to 4226) | 7.98<br>(6.11 to 10.43) | 1.82<br>(1.2 to 2.58) | -0.75<br>(-0.89 to -0.61) |
| Tajikistan | 10<br>(8 to 14) | 0.37<br>(0.27 to 0.49) | 37<br>(26 to 51) | 1.3<br>(0.91 to 1.81) | 2.61<br>(1.8 to 3.61) | 4.95<br>(4.59 to 5.32) |
| Thailand | 3411<br>(2789 to 4242) | 11.42<br>(9.36 to 13.9) | 11604<br>(8463 to 15407) | 11.78<br>(8.63 to 15.58) | 2.4<br>(1.51 to 3.67) | 0.11<br>(0 to 0.22) |
| Timor-Leste | 30<br>(22 to 43) | 12.53<br>(9.27 to 17.59) | 89<br>(65 to 118) | 12.72<br>(9.56 to 16.54) | 1.97<br>(1.15 to 3.01) | 0.06<br>(-0.11 to 0.23) |

| | | | | | | |
|---|---|---|---|---|---|---|
| Togo | 143 (111 to 186) | 14.52 (11.34 to 18.84) | 352 (268 to 455) | 12.68 (9.93 to 15.89) | 1.46 (0.86 to 2.21) | -0.47 (-0.5 to -0.44) |
| Tokelau | 0 (0 to 0) | 11.22 (8.37 to 14.95) | 0 (0 to 0) | 14.15 (10.71 to 18.92) | 0.21 (-0.09 to 0.59) | 0.73 (0.49 to 0.97) |
| Tonga | 5 (4 to 6) | 10 (7.79 to 13.23) | 11 (9 to 15) | 14.65 (11.18 to 19.66) | 1.36 (0.81 to 2.08) | 1.28 (0.89 to 1.67) |
| Trinidad and Tobago | 43 (35 to 52) | 5.93 (4.87 to 7.06) | 164 (117 to 220) | 9.14 (6.49 to 12.3) | 2.8 (1.86 to 4) | 2.04 (1.8 to 2.28) |
| Tunisia | 320 (245 to 413) | 8.5 (6.52 to 10.85) | 910 (632 to 1269) | 8.39 (5.85 to 11.58) | 1.84 (0.94 to 2.94) | -0.19 (-0.28 to -0.1) |
| Turkey | 3497 (2543 to 5199) | 11.72 (8.48 to 17.9) | 6650 (4938 to 8557) | 8.24 (6.1 to 10.63) | 0.9 (0.2 to 1.64) | -1.15 (-1.5 to -0.8) |
| Turkmenistan | 22 (17 to 29) | 1.28 (0.97 to 1.67) | 59 (42 to 82) | 1.71 (1.22 to 2.36) | 1.65 (1 to 2.47) | 0.46 (0.11 to 0.8) |
| Tuvalu | 1 (1 to 1) | 12.86 (9.53 to 16.96) | 1 (1 to 2) | 16.84 (11.93 to 23.33) | 0.97 (0.46 to 1.7) | 0.87 (0.65 to 1.1) |
| Uganda | 442 (318 to 588) | 8.92 (6.48 to 11.73) | 966 (720 to 1265) | 9.17 (6.9 to 12.11) | 1.18 (0.66 to 1.8) | -0.13 (-0.24 to -0.02) |
| Ukraine | 376 (300 to 460) | 0.64 (0.52 to 0.78) | 679 (528 to 846) | 0.93 (0.73 to 1.14) | 0.81 (0.54 to 1.14) | 1.36 (1.21 to 1.52) |
| United Arab Emirates | 50 (32 to 67) | 21.12 (12.12 to 27.41) | 348 (212 to 582) | 16.98 (9.74 to 26.63) | 5.92 (3.35 to 9.46) | -0.76 (-1.34 to -0.18) |
| United Kingdom | 1488 (1156 to 1847) | 1.61 (1.27 to 1.98) | 2643 (2026 to 3268) | 1.72 (1.32 to 2.12) | 0.78 (0.57 to 1.01) | -0.07 (-0.64 to 0.51) |
| United Republic of Tanzania | 683 (525 to 894) | 8.59 (6.62 to 11.01) | 1223 (1007 to 1482) | 6.57 (5.4 to 7.96) | 0.79 (0.38 to 1.35) | -1.28 (-1.53 to -1.03) |

| | | | | | | |
|---|---|---|---|---|---|---|
| United States of America | 13960<br>(11613 to 16121) | 4.11<br>(3.43 to 4.72) | 43329<br>(35082 to 50786) | 6.87<br>(5.59 to 8.03) | 2.1<br>(1.78 to 2.47) | 2.23<br>(2.04 to 2.41) |
| United States Virgin Islands | 4<br>(3 to 5) | 5.63<br>(4.33 to 7.13) | 14<br>(11 to 18) | 8.29<br>(6.55 to 10.24) | 2.63<br>(1.8 to 3.64) | 1.97<br>(1.71 to 2.23) |
| Uruguay | 186<br>(165 to 206) | 4.85<br>(4.31 to 5.35) | 408<br>(336 to 490) | 6.21<br>(5.17 to 7.41) | 1.2<br>(0.86 to 1.57) | 1.32<br>(1.15 to 1.49) |
| Uzbekistan | 155<br>(109 to 240) | 1.45<br>(1.01 to 2.32) | 294<br>(215 to 402) | 2.24<br>(1.62 to 3.03) | 0.9<br>(0.22 to 1.61) | 1.26<br>(0.39 to 2.14) |
| Vanuatu | 5<br>(3 to 7) | 9.37<br>(6.52 to 13.19) | 22<br>(16 to 31) | 15.23<br>(11.05 to 21.37) | 3.38<br>(2.12 to 5.37) | 1.87<br>(1.74 to 1.99) |
| Venezuela (Bolivarian Republic of) | 359<br>(292 to 442) | 4.29<br>(3.49 to 5.23) | 2668<br>(1926 to 3637) | 9.69<br>(7.01 to 13.2) | 6.44<br>(4.61 to 8.79) | 2.53<br>(2.02 to 3.04) |
| Viet Nam | 4324<br>(3332 to 5525) | 12.1<br>(9.42 to 15.47) | 8651<br>(6455 to 10818) | 11.16<br>(8.48 to 13.75) | 1<br>(0.33 to 1.68) | -0.37<br>(-0.83 to 0.08) |
| Yemen | 325<br>(224 to 457) | 8.69<br>(6.11 to 12.41) | 841<br>(616 to 1172) | 8.39<br>(6.1 to 11.69) | 1.59<br>(0.93 to 2.49) | -0.15<br>(-0.22 to -0.07) |
| Zambia | 242<br>(189 to 308) | 11.35<br>(8.8 to 14.31) | 532<br>(398 to 692) | 10.77<br>(8.22 to 13.83) | 1.2<br>(0.55 to 1.96) | -0.44<br>(-0.6 to -0.28) |
| Zimbabwe | 380<br>(274 to 568) | 12.04<br>(8.85 to 18.18) | 879<br>(614 to 1313) | 16.01<br>(11.37 to 23.88) | 1.31<br>(0.76 to 1.97) | 1.3<br>(1 to 1.61) |

Table S9: Changes in incidence number according to population-level determinants and causes from 1990 to 2019 globally and by SDI quintile

| location | Overall difference | Aging | Population growth | Epidemiologic changes | Aging_percent (%) | Population growth _percent (%) | Epidemiologic changes_percent (%) |
|---|---|---|---|---|---|---|---|
| Global | 976152.38 | 291291.345 | 476849.254 | 208011.778 | 29.84 | 48.85 | 21.31 |
| High SDI | 248693.23 | 112717.506 | 93341.622 | 42634.105 | 45.32 | 37.53 | 17.14 |
| High-middle SDI | 215455.92 | 79217.225 | 77764.545 | 58474.145 | 36.77 | 36.09 | 27.14 |
| Middle SDI | 327695.69 | 112325.163 | 127882.311 | 87488.214 | 34.28 | 39.02 | 26.7 |
| Low-middle SDI | 141732.96 | 30362.192 | 75556.52 | 35814.253 | 21.42 | 53.31 | 25.27 |
| Low SDI | 41980.62 | -1237.938 | 32378.248 | 10840.31 | -2.95 | 77.13 | 25.82 |

Table S10: Changes in deaths number according to population-level determinants and causes from 1990 to 2019 globally and by SDI quintile

| location | Overall difference | Aging | Population growth | Epidemiologic changes | Aging_percent (%) | Population growth _percent (%) | Epidemiologic changes_percent (%) |
|---|---|---|---|---|---|---|---|
| Global | 278269.87 | 106424.69 | 135647.993 | 36197.183 | 38.25 | 48.75 | 13.01 |
| High SDI | 68039.55 | 32626.226 | 16994.418 | 18418.902 | 47.95 | 24.98 | 27.07 |
| High-middle SDI | 43070.9 | 24312.959 | 17286.439 | 1471.497 | 56.45 | 40.13 | 3.42 |
| Middle SDI | 103806.52 | 49234.052 | 47820.085 | 6752.386 | 47.43 | 46.07 | 6.5 |
| Low-middle SDI | 46191.59 | 15968.711 | 30583.776 | -360.9 | 34.57 | 66.21 | -0.78 |
| Low SDI | 16973.68 | 473.052 | 20275.926 | -3775.298 | 2.79 | 119.46 | -22.24 |

Table S11: Incidence of CKD due to hypertension in global 1990-2019 APC Model Analysis

| Factors | CKD incidence | | | |
|---|---|---|---|---|
| | Coefficient | RR | 95% CI | |
| | | | Lower | Upper |
| Age | | | | |
| 15-19 | -2.051332 | 0.128563543 | 0.128381754 | 0.128745718 |
| 20-24 | -1.778653 | 0.168865456 | 0.168676095 | 0.169055198 |
| 25-29 | -1.432682 | 0.238667956 | 0.238442998 | 0.238893365 |
| 30-34 | -1.175384 | 0.308700416 | 0.308442142 | 0.308959215 |
| 35-39 | -0.9808222 | 0.375002645 | 0.374715503 | 0.375290007 |
| 40-44 | -0.7912175 | 0.453292575 | 0.452971622 | 0.453613802 |
| 45-49 | -0.605703 | 0.545690671 | 0.545327417 | 0.546054113 |
| 50-54 | -0.3707891 | 0.690189487 | 0.689762047 | 0.690617192 |
| 55-59 | -0.0979568 | 0.906688072 | 0.906171769 | 0.907204668 |
| 60-64 | 0.1819941 | 1.199607116 | 1.198981564 | 1.200232874 |
| 65-69 | 0.4980219 | 1.645463159 | 1.644666125 | 1.646260579 |
| 70-74 | 0.83842 | 2.312710007 | 2.311648948 | 2.313771553 |
| 75-79 | 1.153157 | 3.16817908 | 3.166734719 | 3.169620929 |
| 80-84 | 1.417616 | 4.127269292 | 4.125263927 | 4.129275633 |
| 85-89 | 1.614676 | 5.026259152 | 5.023415094 | 5.02909979 |
| 90-94 | 1.746692 | 5.735597902 | 5.731349398 | 5.739855295 |
| 95 plus | 1.833962 | 6.25863431 | 6.251278482 | 6.265998794 |
| Period | | | | |
| 1994 | -0.3658435 | 0.693611343 | 0.693355725 | 0.693867055 |
| 1999 | -0.2219881 | 0.800924895 | 0.800659273 | 0.801190606 |
| 2004 | -0.0685145 | 0.93377992 | 0.933499828 | 0.934060003 |
| 2009 | 0.0830665 | 1.086614066 | 1.086308879 | 1.08691923 |
| 2014 | 0.223028 | 1.249855569 | 1.249511781 | 1.250199327 |
| 2019 | 0.3502517 | 1.419424773 | 1.419015186 | 1.419834478 |
| Cohort | | | | |
| 1895-1899 | 1.200442 | 3.321584739 | 3.308430805 | 3.334790971 |
| 1900-1904 | 1.077875 | 2.938428751 | 2.932774784 | 2.944093618 |
| 1905-1909 | 0.9527831 | 2.592915971 | 2.58976283 | 2.596072692 |
| 1910-1914 | 0.8400613 | 2.316508974 | 2.314370203 | 2.31864949 |
| 1915-1919 | 0.7233319 | 2.061289793 | 2.059673963 | 2.06290689 |
| 1920-1924 | 0.6118161 | 1.843776843 | 1.842507287 | 1.845047274 |

| | | | | |
|---|---|---|---|---|
| 1925-1929 | 0.4929761 | 1.637181392 | 1.636144075 | 1.638219203 |
| 1930-1934 | 0.3764323 | 1.457076892 | 1.456197083 | 1.457957232 |
| 1935-1939 | 0.2650765 | 1.303530692 | 1.302753368 | 1.304308611 |
| 1940-1944 | 0.1579135 | 1.171064893 | 1.170351815 | 1.171778406 |
| 1945-1949 | 0.0518265 | 1.053192998 | 1.052520854 | 1.053865571 |
| 1950-1954 | -0.0599529 | 0.941808892 | 0.94118675 | 0.94243135 |
| 1955-1959 | -0.1681263 | 0.845247073 | 0.844663969 | 0.845830664 |
| 1960-1964 | -0.2721522 | 0.761738316 | 0.761182906 | 0.762294207 |
| 1965-1969 | -0.389974 | 0.677074478 | 0.676561721 | 0.677587624 |
| 1970-1974 | -0.5030363 | 0.604691844 | 0.60420913 | 0.605174883 |
| 1975-1979 | -0.6160093 | 0.540095502 | 0.539633702 | 0.540557697 |
| 1980-1984 | -0.7285918 | 0.482588092 | 0.482120402 | 0.483056285 |
| 1985-1989 | -0.8365169 | 0.433216836 | 0.432735064 | 0.433699145 |
| 1990-1994 | -0.9472236 | 0.387816263 | 0.387289037 | 0.388344246 |
| 1995-1999 | -1.058154 | 0.347095958 | 0.346449574 | 0.347743549 |
| 2000-2004 | -1.170797 | 0.310119677 | 0.309130117 | 0.311112405 |
| Intercept | -5.112672 | | | |
| Log likelihood | -280465.7287 | | | |
| AIC | 1833.384 | | | |
| BIC | 554823.8 | | | |

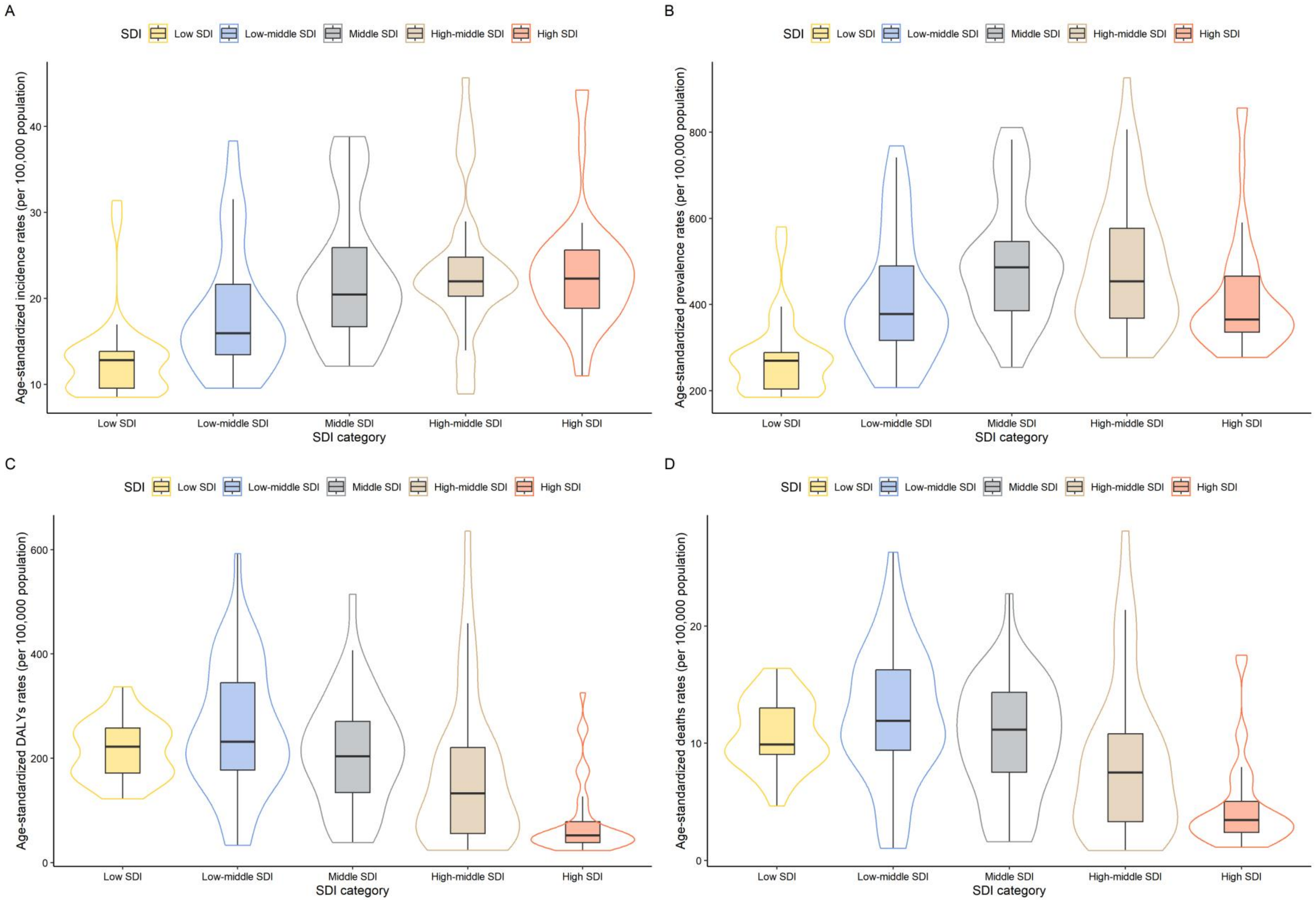


Figure S1: SDI-specific burden in terms of age-standardized incidence rates (ASIR) (A), age-standardized prevalence rates (ASPR) (B), age-standardized disability-adjusted life years rates (ASDR) (C), and age-standardized mortality rates (ASMR) (D) in 204 countries.

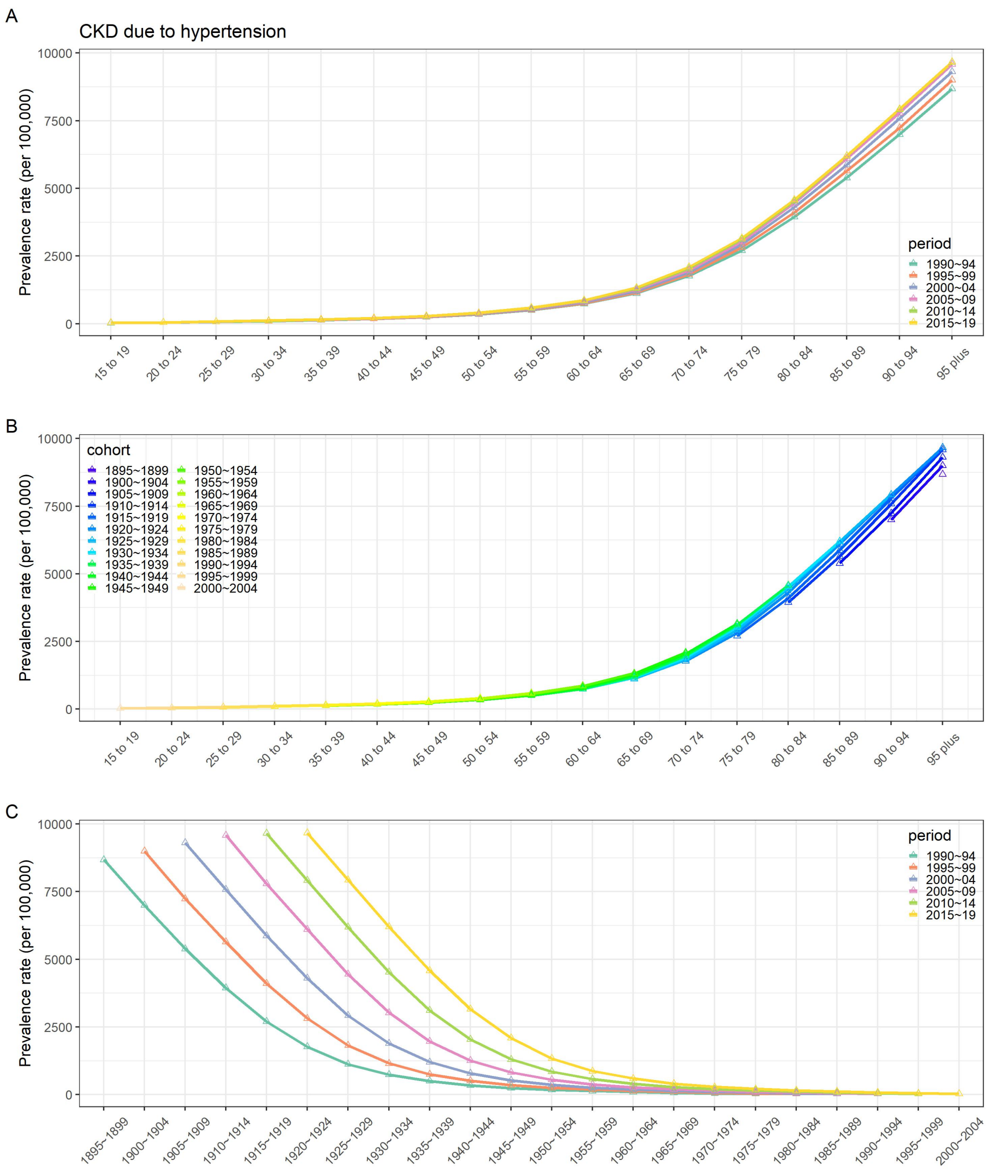
A
CKD due to hypertension
Prevalence rate (per 100,000)
period
1990~94
1995~99
2000~04
2005~09
2010~14
2015~19
B
cohort
C

Figure S2: Age base variation of CKD due to hypertension prevalence, by period (A); age base variation of CKD due to hypertension prevalence, by cohort (B); cohort base variation of CKD due to hypertension prevalence, by period (C).

| | Global | Low SDI | Low-middle SDI | Middle SDI | High-middle SDI | High SDI | Central Asia | Central Europe | Eastern Europe | Australasia | High-income Asia Pacific | High-income North America | Western Europe | Andean Latin America | Caribbean | Central Latin America | Southern Latin America | Tropical Latin America | North Africa and Middle East | South Asia | East Asia | Southest Asia | Central Sub-Saharan Africa | Eastern Sub-Saharan Africa | Southern Sub-Saharan Africa | Western Sub-Saharan Africa |
|---|---|---|---|---|---|---|---|---|---|---|---|---|---|---|---|---|---|---|---|---|---|---|---|---|---|---|
| Ischemic heart disease | 1284.62 | 655.08 | 1206.98 | 1368.90 | 1865.82 | 1101.34 | 2653.80 | 2794.66 | 4883.50 | 814.00 | 622.98 | 1225.71 | 1130.85 | 433.16 | 1403.40 | 886.22 | 821.01 | 957.03 | 1653.97 | 1352.48 | 1292.96 | 1192.25 | 516.52 | 384.77 | 770.26 | 491.34 |
| Intracerebral hemorrhage | 525.81 | 384.81 | 582.18 | 676.19 | 545.16 | 201.61 | 823.47 | 558.85 | 661.56 | 109.70 | 258.65 | 172.40 | 189.85 | 136.77 | 529.63 | 187.58 | 276.24 | 322.71 | 248.68 | 446.63 | 910.31 | 1020.77 | 430.06 | 398.99 | 399.53 | 380.77 |
| Ischemic stroke | 420.23 | 147.44 | 297.08 | 492.13 | 731.33 | 329.61 | 542.09 | 1138.45 | 1529.99 | 206.09 | 411.43 | 256.23 | 340.65 | 106.11 | 342.78 | 147.01 | 247.00 | 292.21 | 408.90 | 216.03 | 757.30 | 536.56 | 144.44 | 133.12 | 317.72 | 142.17 |
| Hypertensive heart disease | 277.97 | 232.95 | 240.35 | 347.19 | 280.43 | 226.07 | 312.73 | 639.89 | 212.96 | 70.12 | 123.51 | 302.27 | 257.40 | 142.96 | 407.96 | 156.97 | 262.94 | 255.99 | 358.20 | 166.32 | 392.66 | 370.88 | 364.60 | 26.46 | 393.86 | 166.90 |
| Chronic kidney disease due to hypertension | 128.76 | 79.04 | 114.83 | 161.92 | 109.75 | 156.66 | 39.97 | 95.48 | 50.29 | 79.44 | 146.58 | 209.21 | 117.91 | 162.90 | 162.25 | 264.34 | 194.79 | 117.54 | 137.15 | 98.18 | 122.57 | 226.40 | 73.63 | 64.49 | 182.20 | 97.31 |
| Subarachnoid hemorrhage | 82.16 | 42.62 | 85.68 | 86.07 | 96.35 | 90.75 | 99.45 | 152.61 | 193.33 | 62.42 | 154.91 | 84.42 | 74.87 | 67.67 | 107.89 | 75.31 | 93.79 | 118.61 | 40.58 | 82.93 | 95.66 | 104.28 | 29.28 | 30.20 | 29.83 | 16.79 |
| Chronic kidney disease due to diabetes mellitus type 2 | 57.16 | 29.14 | 53.28 | 74.24 | 49.17 | 66.00 | 45.88 | 46.00 | 18.39 | 20.26 | 82.26 | 78.09 | 44.47 | 62.72 | 80.39 | 137.34 | 82.01 | 53.49 | 66.80 | 49.91 | 53.54 | 94.98 | 24.76 | 22.84 | 59.15 | 30.71 |
| Chronic kidney disease due to other and unspecified causes | 50.22 | 28.71 | 52.77 | 56.13 | 48.92 | 57.55 | 70.86 | 68.09 | 39.13 | 59.83 | 55.68 | 56.13 | 65.30 | 56.55 | 80.12 | 124.11 | 72.96 | 61.15 | 61.57 | 58.13 | 36.97 | 38.98 | 19.99 | 18.88 | 57.36 | 21.74 |
| Atrial fibrillation | 43.14 | 13.25 | 27.13 | 36.67 | 63.89 | 90.34 | 40.35 | 115.56 | 103.82 | 112.87 | 50.38 | 96.34 | 119.53 | 16.64 | 22.05 | 28.38 | 49.58 | 42.01 | 19.90 | 28.14 | 49.02 | 36.17 | 13.88 | 9.43 | 22.37 | 11.98 |
| Other cardiovascular and circulatory diseases | 36.96 | 37.82 | 37.03 | 29.82 | 41.30 | 46.59 | 35.67 | 99.86 | 68.07 | 31.45 | 25.54 | 38.23 | 63.63 | 17.91 | 57.58 | 26.76 | 71.32 | 46.88 | 54.48 | 33.85 | 16.94 | 24.13 | 37.18 | 34.46 | 45.27 | 55.28 |
| Rheumatic heart disease | 31.62 | 38.72 | 54.71 | 27.91 | 19.07 | 9.99 | 44.56 | 16.83 | 22.41 | 9.66 | 7.80 | 8.44 | 13.98 | 9.94 | 34.26 | 8.10 | 15.75 | 19.12 | 13.95 | 72.54 | 31.55 | 11.49 | 22.28 | 15.73 | 28.98 | 15.32 |
| Chronic kidney disease due to glomerulonephritis | 27.47 | 23.95 | 31.37 | 30.84 | 22.57 | 23.57 | 25.41 | 28.44 | 31.94 | 39.46 | 14.56 | 28.85 | 22.44 | 42.72 | 63.03 | 107.72 | 23.87 | 46.66 | 16.65 | 30.27 | 17.17 | 18.18 | 22.11 | 24.41 | 51.59 | 26.34 |
| Other cardiomyopathy | 25.47 | 16.79 | 11.29 | 16.08 | 57.19 | 37.26 | 120.87 | 127.12 | 156.87 | 25.64 | 27.34 | 47.30 | 41.71 | 7.44 | 33.09 | 12.62 | 66.49 | 77.61 | 13.13 | 1.48 | 6.97 | 25.27 | 40.59 | 21.11 | 72.76 | 26.59 |
| Aortic aneurysm | 16.04 | 6.73 | 9.97 | 11.03 | 26.56 | 34.03 | 16.35 | 48.70 | 57.71 | 31.10 | 53.15 | 23.10 | 40.11 | 6.89 | 22.15 | 11.09 | 29.48 | 41.48 | 7.94 | 9.38 | 10.32 | 11.30 | 9.49 | 7.42 | 13.48 | 5.65 |
| Chronic kidney disease due to diabetes mellitus type 1 | 14.38 | 9.82 | 16.11 | 19.96 | 11.04 | 7.94 | 8.36 | 6.47 | 9.49 | 2.85 | 8.95 | 7.93 | 4.67 | 12.73 | 21.78 | 34.74 | 12.71 | 12.65 | 11.36 | 15.18 | 12.60 | 34.09 | 8.82 | 7.26 | 18.68 | 11.27 |
| Non-rheumatic calcific aortic valve disease | 8.12 | 2.86 | 3.91 | 3.94 | 10.06 | 28.45 | 2.54 | 28.84 | 9.59 | 25.61 | 22.15 | 24.56 | 42.08 | 3.06 | 8.45 | 6.64 | 21.21 | 12.05 | 6.21 | 3.76 | 2.24 | 2.35 | 3.12 | 2.67 | 5.27 | 2.90 |
| Endocarditis | 7.04 | 4.43 | 5.44 | 6.11 | 8.80 | 12.40 | 3.50 | 8.64 | 17.84 | 8.59 | 9.07 | 13.06 | 15.66 | 3.50 | 9.61 | 4.91 | 16.05 | 11.91 | 4.57 | 5.00 | 2.92 | 12.60 | 5.77 | 5.29 | 9.29 | 4.02 |
| Peripheral artery | 5.48 | 1.83 | 2.06 | 3.06 | 10.67 | 13.94 | 1.74 | 19.41 | 33.58 | 15.24 | 5.14 | 16.02 | 20.15 | 1.09 | 8.86 | 3.09 | 5.53 | 6.79 | 1.90 | 1.64 | 3.33 | 2.82 | 4.26 | 2.57 | 9.27 | 1.18 |

1000+ 900 800 700 600 500 400 300 200 100 0

Figure S3: All age DALY rates (per 100,000 population) for 18 diseases attributable to hypertension by regions, for both sexes, 2019.

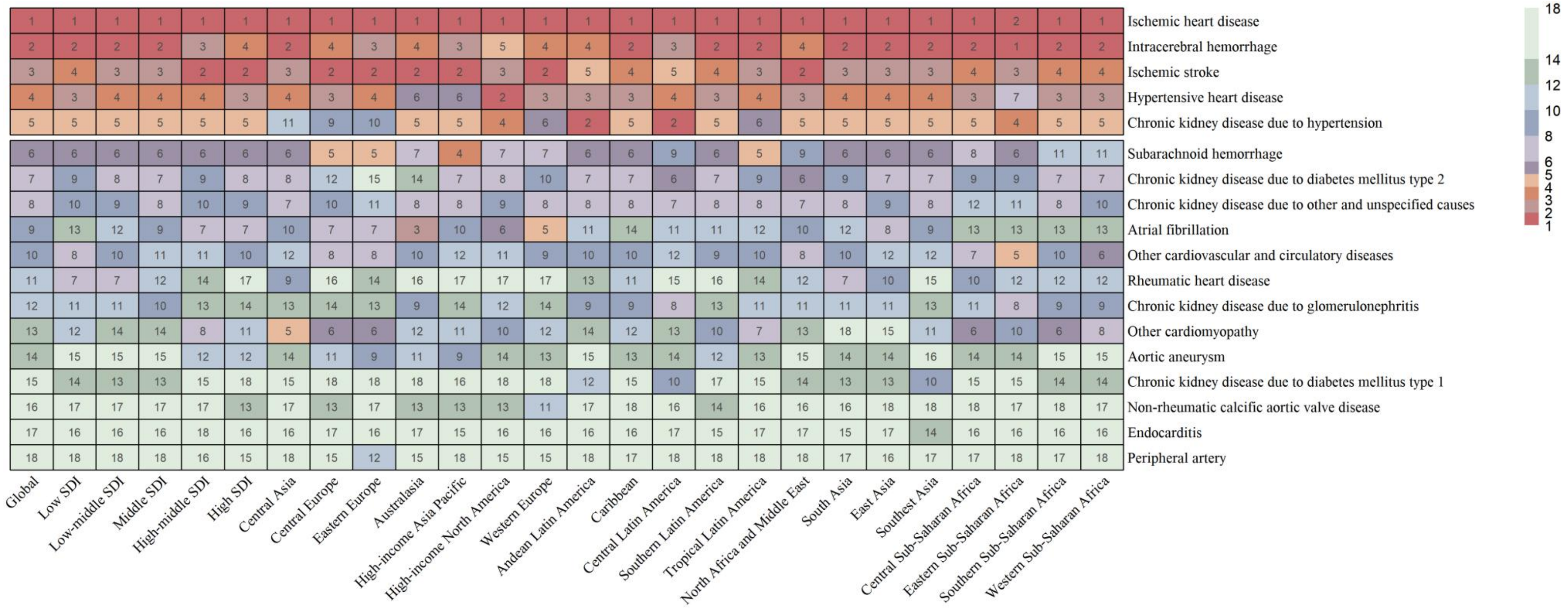


Figure S4: Ranking of all age DALY rates for 18 diseases attributable to hypertension by regions, for both sexes, 2019.
